# Early-life Urban Environment, Nutrition, and Pubertal Timing in Southern Europe: An Exposome Analysis

**DOI:** 10.64898/2026.06.09.26355261

**Authors:** Marta Pinto da Costa, María Arteaga Jover, Adrià Setó Llorens, Aurélie Portefaix, Ana Isabel Ribeiro, Susana Santos, Maria-Jose Lopez-Espinosa, Carmen Iñiguez, Mikel Subiza-Pérez, Ane Arregi, Rosaura Leis, Gloria Bueno, Xavier Basagaña, Marta Cirach, Mark Nieuwenhuijsen, Mònica Guxens, Martine Vrijheid, Carmen Freire, Joana Araújo, Sofia Vilela, Augusto Anguita-Ruiz

## Abstract

**Background:** Urban environmental and lifestyle factors during early life may influence pubertal timing, but the combined effects of multiple environmental exposures within an exposome analytical framework remain poorly understood.

**Objective:** To examine the association between early-life urban environmental exposures and pubertal timing, and to explore whether these exposures interact with early-life nutritional factors, namely breastfeeding duration and childhood diet quality.

**Methods:** Data from two European population-based birth cohorts were analysed: Generation XXI (G21, Portugal; n=5263; 51.5% girls) and INfancia y Medio Ambiente (INMA, Spain; n=1019; 50.1% girls). Urban environmental exposures including indicators of air pollution, traffic, built environment, and natural spaces were estimated at 4 early-life stages at both cohorts: pregnancy (INMA only), birth, 1 year, and 4–5 years of age. Pubertal development timing was assessed using Tanner staging and/or the Pubertal Development Scale (PDS), and age at menarche was self-reported. Exposome-Wide Association Study (ExWAS) models and unsupervised clustering followed by ordinal logistic regression models were used to examine single- and multi-exposure associations, respectively. Regression models were fitted adjusting for relevant child characteristics, maternal factors, and household socioeconomic conditions, and corrected for multiple testing.

**Results:** Individuals living in more unfavourable urban environments characterised by higher building density, air pollution, and lower access to natural spaces showed earlier pubertal timing according to multiple outcomes, across multiple early-life exposure periods, and in both cohorts. In the G21 cohort, these environmental profiles were associated with earlier age at menarche, particularly for exposures at 1-1.5 and 4-5 years (e.g., 1-1.5y: β=-0.172, FDR-adjusted p-value=0.041), while in the INMA cohort, boys exposed to more unfavourable environmental profiles showed more advanced pubertal development, also particularly for exposures at 1-1.5 and 4-5 years of age (e.g., 1-1.5y; β=0.572, FDR-adjusted p-value=0.008). Among environmental domains, air pollution and traffic were the factors most consistently associated with pubertal timing. Regarding early-life nutritional factors, longer duration of exclusive breastfeeding was associated with a lower Tanner stage among girls in G21. No significant interactions between breastfeeding duration and environmental exposure clusters were observed.

**Conclusion:** Early-life urban environmental exposures, particularly air pollution and traffic, may influence pubertal timing. Exclusive breastfeeding may have a protective role against earlier pubertal development. These findings highlight the importance of improving urban environmental conditions and promoting breastfeeding to support healthy developmental trajectories.

**Graphical abstract:** 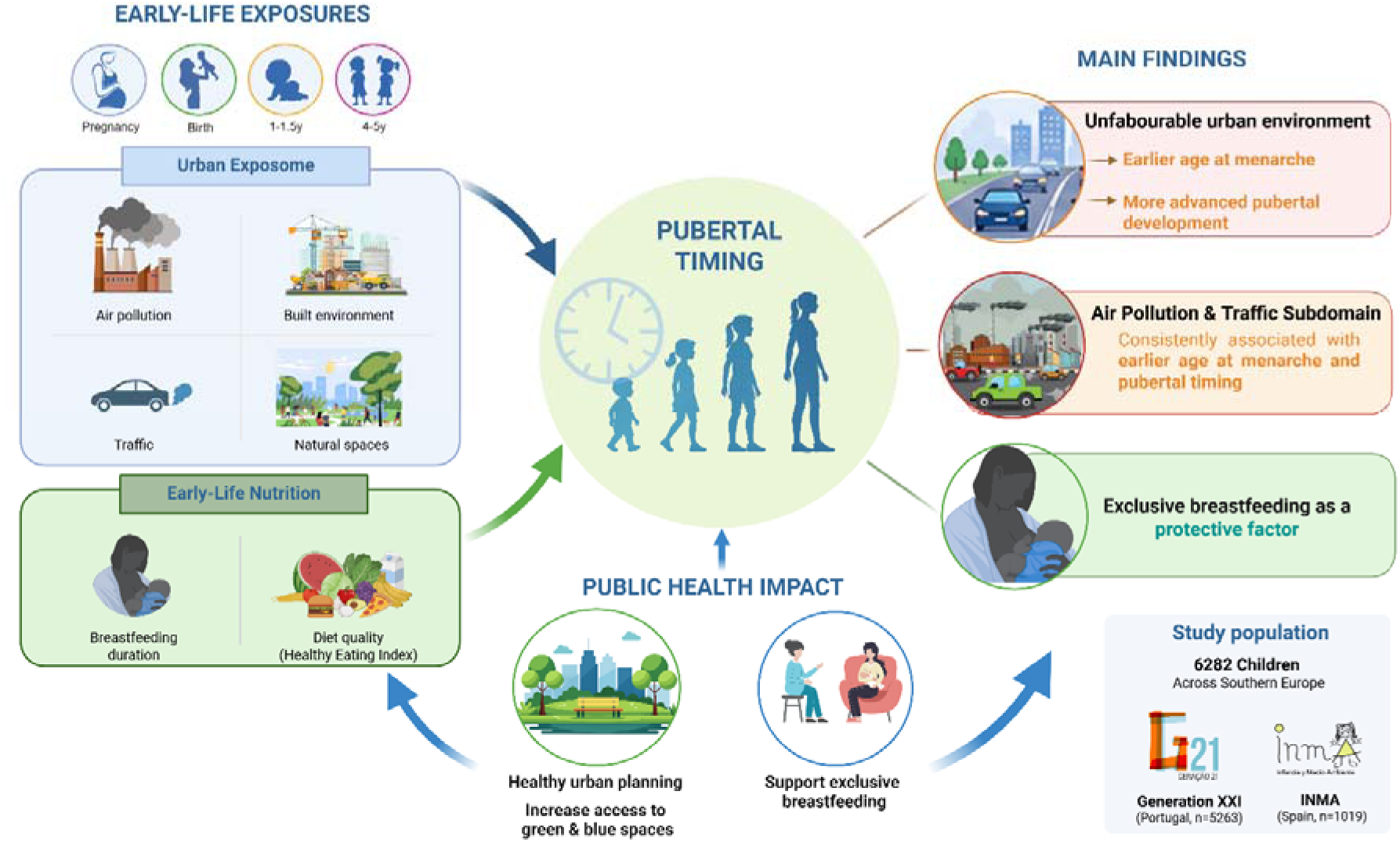

## 1. Introduction

A growing body of evidence indicates a worldwide declining trend in the age at menarche and pubertal onset (1–6). This trend raises public health concerns, as earlier pubertal timing carries significant health implications, including an increased risk of metabolic disorders (7), cardiovascular disease (8, 9), psychological and behavioural problems (10–12), substance use (13), fertility impairment (14), and a higher risk of breast and other reproductive cancers (15–17).

Pubertal timing is a complex and multifactorial trait shaped by both genetic and non-genetic influences (18). Although genetic predisposition plays a major role in determining pubertal timing, existing scientific evidence points to specific external and personal environmental factors as additional relevant contributors to this relationship: breastfeeding, nutrition, socioeconomic status, and environmental exposures (18–21). The study of urban environmental factors as determinants of health and disease has gained a lot of attention with the advent of exposome research. The exposome framework provides a comprehensive strategy to explore environmental risk factors by capturing the totality of exposures across the life course (22, 23). Within the Developmental Origins of Health and Disease (DOHaD) paradigm, the prenatal and early-life period is increasingly recognized as a sensitive window in which environmental exposures can program long-term health. (22, 24).

Concerning urban environmental exposures, several studies have evaluated their association with pubertal timing. Most studies examining the impact of the urban environment on pubertal development have focused on air pollution, with some reporting associations with earlier pubertal onset, while others have not confirmed these findings (25–28). For instance, living within 150 meters of a major road has been associated with earlier pubertal onset in girls (25), and residence in urban area has been linked with earlier age at menarche (29,30), potentially reflecting differences in health, nutrition, socioeconomic conditions (31, 32), and exposure to environmental pollutants during childhood compared with rural settings (32). Conversely, other studies have found that in utero and childhood exposure to Particulate Matter (PMLL) was associated with a decreased age at menarche (27), while exposure to sulfur dioxide and nitrogen dioxide was associated with delayed pubertal onset in both sexes (26). In contrast, a study conducted in two European birth cohorts did not find any significant associations between ambient air pollutants and pubertal development (28).

Some authors have further hypothesized that proximity to green spaces may influence the age at menarche (33), by reducing stress, lowering adiposity risk, and promoting opportunities for physical activity - factors known to be associated with the timing of reproductive development (32, 34, 35). Nevertheless, the only available study, conducted among adolescents in Germany and Australia, did not support the hypothesis (36).

As described, current epidemiological evidence on the association between urban environmental exposures and pubertal timing is limited. Moreover, previous studies have mainly assessed single environmental exposures, without accounting for the complex correlations and interactions between them, highlighting the need for exposome-based approaches in big multi-cohort studies. To date, the impact of the totality of urban environmental exposures (external exposome landscape) that an individual experiences across early life on pubertal timing remains largely unexplored.

Regarding personal exposome factors, breastfeeding and childhood nutrition are also modifiable factors that appear to play a significant role in pubertal timing. (37–39) While some studies have found no association between breastfeeding and puberty onset (40, 41), an increasing body of evidence suggests that breastfeeding, and longer breastfeeding duration, may act as protective factors against early pubertal onset, particularly in girls (42–44). Regarding childhood nutrition, although most studies have examined specific nutrients (21, 37, 45–49) or food groups (37, 46, 47) rather than overall dietary quality, higher diet quality has been associated with a later age at puberty onset (50, 51).

To address these gaps, the present study aimed to explore the influence of early-life urban environmental exposures, measured prenatally (pregnancy) and postnatally (at birth, 1 year, and 4-5 years), on pubertal timing within an exposome framework in two well-known birth exposome cohorts representative of South Europe (Spain and Portugal). In addition, this study explored how these environmental exposures interact with early-life nutritional factors, such as breastfeeding duration and diet quality, in order to determine which of these modifiable risk factors plays a more decisive role in pubertal timing (Figure 1).

**Figure 1.**
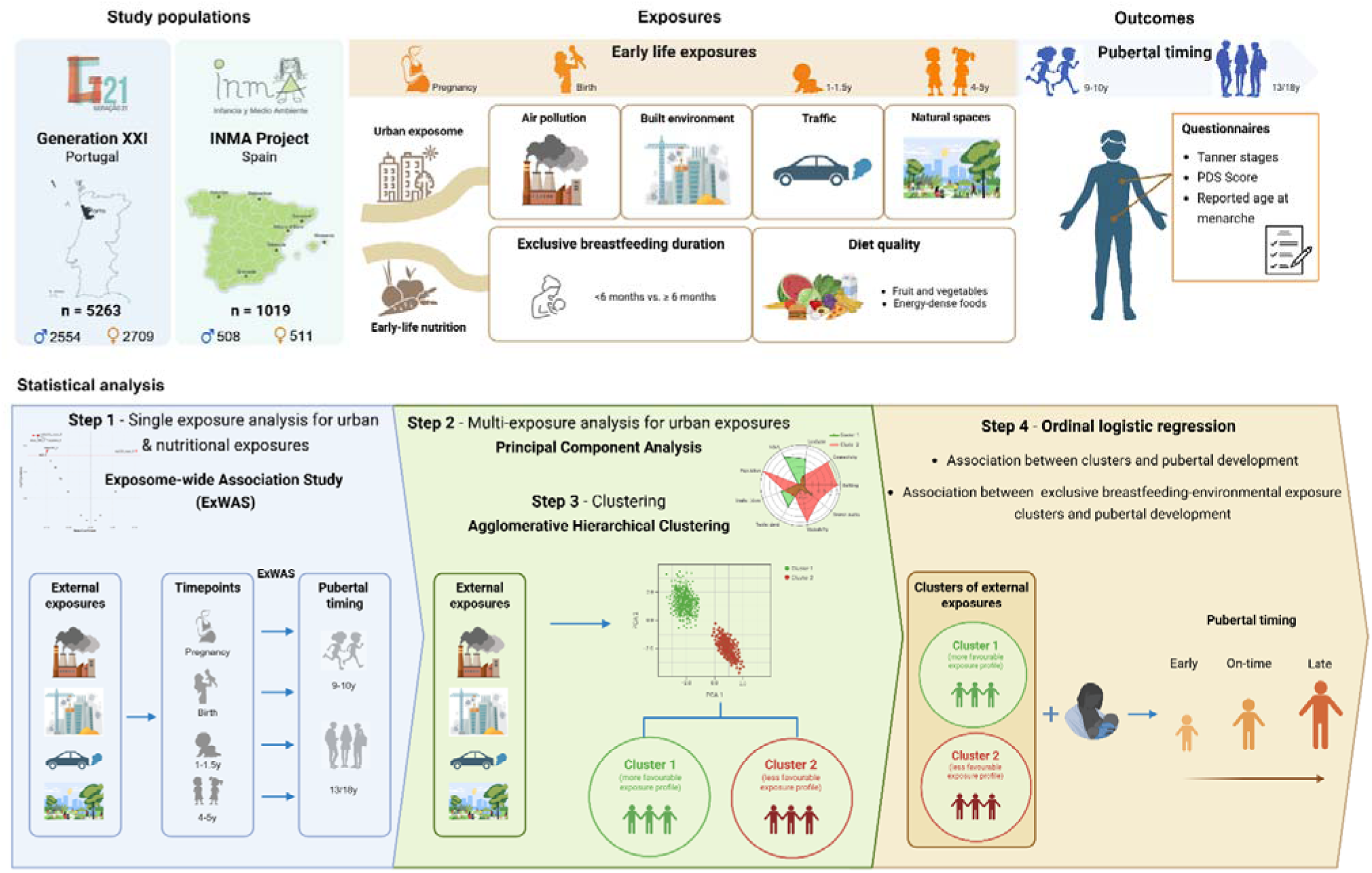
Overview of the study design, exposure and outcome assessment, and analytical framework. Figure 1 presents an overview of the study design, including the participating cohorts, the early-life urban environmental and nutritional exposures, pubertal outcomes, and the analytical strategy applied in the INMA and G21 cohorts. Early-life urban environmental exposures, including built environment, natural spaces, air pollution and traffic, were assessed across different life stages. Statistical analysis included Exposome-wide Association Study (ExWAS), Principal Component Analysis, Agglomerative Hierarchical Clustering, and Ordinal Logistic Regression models to evaluate associations with pubertal timing.

## 2. Methods

### 2.1. Study population

The present study included data from two independent European birth cohorts: Generation XXI (G21) in Portugal and INfancia y Medio Ambiente (INMA) in Spain (Supplementary Figure 1). G21 is a population-based cohort previously described (52, 53). For this cohort, 8647 newborns delivered in the five maternity hospitals in the metropolitan area of Porto were invited and enrolled at baseline. All families were invited to participate in follow-up assessments when children were aged 4 (86% of participation), 7 (80% of participation), 10 (76% of participation), 13 (54% of participation – follow-up was interrupted due to the COVID-19 pandemic), and 18 years (41% of participation). At each evaluation wave, data were collected by trained interviewers through structured questionnaires, including information on sociodemographic conditions, health status, lifestyle, biological samples, as well as objective anthropometric measures. For the present study, we included individuals with complete data on pubertal development (Tanner Stage and/or age at menarche) assessed at 10, 13, and/or 18 years, as well as breastfeeding history and dietary intake collected at 4 years of age., and data on urban environmental exposures available at least at birth, 1 year and 4-5 years of age. After excluding twins (n=144) and children with congenital anomalies or diseases that might influence the analysis of foo5263d intake or pubertal development (n=50), a final sample of 5263 children from the G21 cohort was considered.

The INMA Project is a Spanish network of population-based birth cohorts that recruited 3768 mother-child pairs during pregnancy between 1997 and 2008 in the Spanish regions of Ribera d’Ebre, Menorca, Granada, Valencia, Sabadell, Asturias and Gipuzkoa (54). The recruitment and general characteristics of the INMA cohorts are described elsewhere (54). For the present study, data from Valencia, Sabadell and Gipuzkoa cohorts were included. Eligibility criteria comprised: residence in one of the study areas, maternal age ≥16 years of age, singleton pregnancy, no assisted conception, intention to deliver at the reference hospital, and ability to communicate in Spanish or the regional language. Although follow-up time points varied slightly across cohorts, mother-child pairs were generally assessed during the third trimester of pregnancy, at birth, at 1-1.5 years, 2-2.5 years, 4-5 years and at approximately 9.5 years of age. At each evaluation wave, data were collected through face-to-face interviews by trained INMA personnel and included questionnaire-based information, clinical records, physical examinations, ultrasound scans, biological samples, biomarkers, dietary assessments, and environmental measurements. For this study, we included all children with complete data on pubertal development (assessed using the Pubertal Development Scale and/or Tanner Stage) and age at menarche at 9.5 years, as well as breastfeeding history and dietary intake assessed at 1-1.5, 4-5 and 9.5 years, and environmental exposure data available during at least one of the following periods: pregnancy, birth, 1-1.5 years, and/or 4-5 years. A final sample of 1019 children from the INMA cohort was considered.

### 2.2. Pubertal timing assessment

In the INMA cohorts, pubertal development was assessed using clinical Tanner staging (55, 56) and/or parental assessment of the child’s pubertal status using the Pubertal Development Scale (PDS) at the 9-year follow-up (57). The PDS is a reliable and valid instrument for assessing pubertal development in children (58, 59), and is based on five physical characteristics, including pubic hair development and skin changes in both sexes, voice deepening and facial hair growth in boys, and breast development in girls. Response options range from 1 (‘not yet started’) to 4 (‘seems complete’). Following the criteria proposed by Carskadon and Acebo (58) and Shirtcliff et al. (60), PDS scores were converted into a 5-point ordinal scale (1 - Prepubertal, 2 - Early pubertal, 3 - Midpubertal, 4 - Late pubertal, 5 - Postpubertal). The Tanner staging of genital development and pubic hair growth was performed by a pediatrician or trained health care professional, according to the five Tanner stages (ranging from 1-prepubertal to 5 – postpubertal). The PDS was used to assess pubertal development in children from the Gipuzkoa and Sabadell cohorts, while both Tanner staging and the PDS were used in children from the Valencia cohort. Information on age at menarche was collected only in the Sabadell cohort.

In the G21 cohort, pubertal development was assessed following the Tanner stage criteria, performed by trained nurses at the 10-year follow-up. In females, pubic hair and breast development were evaluated, while in males, pubic hair development was assessed according to Tanner staging, and testicular volume was additionally measured. Each parameter was classified from Tanner stage 1 (prepubertal) to Tanner stage 5 (postpubertal). Tanner 1 indicated that puberty had not started yet, whereas Tanner stage 2 was defined as the mark of the beginning of puberty in both cohorts. Females were classified as Tanner stage 2 or higher if they presented slightly pigmented and downy pubic hair, and breast bud stage with an elevation of breast and papilla enlargement of the aureola. Males were classified as Tanner stage 2 if they presented a testicular volume higher than 4ml, and also a slightly pigmented and downy pubic hair. At the 13- and 18-year follow-ups, girls self-reported their age at menarche. The final age at menarche variable was derived as follows: if age at menarche was reported at only one follow-up, that value was considered as the final one; if reported at both follow-ups, and the difference between reports did not exceed two years, the mean of the two values was calculated and considered as the final age at menarche.

### 2.3. Assessment of the urban environment during early-life

Urban environmental exposures considered in this study were obtained using Geographic Information Systems based on the mothers’ residential addresses during pregnancy, at birth, and annually throughout the child’s life in both cohorts. In the present study, information collected during pregnancy (available only in the INMA cohorts), at birth, at 1-1.5 years, and at 4-5 years of age was included. Urban environment variables were selected based on literature and data availability in the current and ongoing G21 and INMA studies, and included indicators of air pollution (e.g., NO_2_, PM_2.5_), built environment (e.g., building density, connectivity density, walkability index, Land use Shannon’s Evenness Index), natural spaces (e.g., NDVI, distance to nearest major blue/green space), and traffic (e.g., inverse distance to nearest road) (Supplementary Table 1). The built environment and natural spaces were measured within different buffer sizes (100m, 300m, and 500m), but in this study, only variables corresponding to the 300m buffer were included. Further details on exposure assessment methods in the INMA and Generation XXI cohorts are available in Supplementary Section I, and have been described previously (61–64).

### 2.4. Breastfeeding and Healthy Eating Index

Information on the child’s diet and breastfeeding was obtained using interviewer-administered questionnaires with the mothers when the child was 6 months old (G21 and INMA), 14–15 months (G21 and INMA), 24 months (G21 only), and 4–5 years of age (G21 and INMA).

In both cohorts, mothers were asked about the duration of any and exclusive breastfeeding. Any breastfeeding was defined as the child receiving any type of breast milk feeding, regardless of whether it was combined with other foods. Exclusive breastfeeding was defined according to the World Health Organization (WHO) definition as the period during which the child received only breast milk and no other foods or liquids (including water or infant formula), while allowing the intake of oral rehydration salt solutions, vitamins, minerals, and medicines (65). However, in the G21 cohort, breastfeeding was classified as exclusive even if the child had already been given water. Information on exclusive breastfeeding was not collected in the Valencia cohort. The duration of non-exclusive breastfeeding was considered as a continuous variable, and the duration of exclusive breastfeeding was considered as both continuous and categorical variables. Children were grouped into two categories: (a) exclusive breastfeeding for < 24 weeks (6 months); and (b) exclusive breastfeeding for ≥ 24 weeks.

Dietary information was collected using a Food Frequency Questionnaire administered at 4 years of age, validated in both cohorts (66, 67). Two food groups were considered for the analysis, ‘Fruit and Vegetables (FV) (fresh fruit and vegetables) and ‘Energy-dense Foods’ (EDF) (cakes, candies, chocolate, sweet pastries, biscuits, added sugar, ice cream, salty snacks, pizza, and chips), as these can be considered indicators of dietary quality. Based on these two food groups, a Healthy Eating Index (HEI) was also developed, adapted from a previously established index in the G21 cohort (68, 69). For each food group, quartiles of consumption were calculated, and the mean values of each quartile were used to define cut-off points. Each quartile was assigned a score ranging from 1 to 4. For the FV group, considered a healthier food group, the lowest quartile of consumption was assigned 1 point, the intermediate quartiles were assigned 2–3 points, and the highest quartile of consumption was assigned 4 points. For the EDF group, which includes less healthy foods, the score was assigned reversely. The scores were summed to obtain a total HEI score, ranging from 2 to 8, with higher scores indicating better overall diet quality.

### 2.5. Other maternal and child characteristics

A comprehensive set of covariates was selected based on previous studies examining associations between single urban environmental exposure, breastfeeding, diet, and pubertal timing. The following variables were considered: child exact age at pubertal assessment (years), child birthweight (grams), maternal pre-pregnancy body mass index (BMI) (kg/m2), maternal age at delivery (years), and household income.

Child BMI z-score was not included as a covariate because it could potentially act as a mediator rather than a confounder in the pathway between early-life exposures and pubertal timing. Therefore, instead of including BMI as a covariate, a sensitivity analysis restricted to children with normal weight was performed to explore the robustness of the associations, acknowledging the potential reduction in statistical power due to the smaller sample size.

### 2.6. Statistical analyses

The whole statistical pipeline is illustrated in Supplementary Figure 2.

#### Data pre-processing

The correlations between exposure variables within each cohort and at each time point were explored by computing Pearson correlation matrices. At this stage, variables with correlations > 0.9 were excluded. This procedure led to the exclusion of NOx due to its high correlation with NO_2_. To minimize the influence of extreme values, winsorization was applied to all numeric variables. Values below the 2.5th percentile and above the 97.5th percentile were replaced with their respective cut-off values.

To handle missing data, an imputation procedure was performed using the MissForest algorithm (70). Variables with more than 20% missing data were excluded from the imputation process, as was the case for child BMI in the INMA cohort (∼38% missingness).

#### Association analysis between pre- and postnatal environmental exposures and pubertal timing

To explore the influence of prenatal and postnatal environmental factors on pubertal timing, we included 23 exposures during pregnancy (INMA only); at birth, 17 exposures in G21 and 9 in INMA; at 1-1.5 years, 19 exposures in G21 and 20 in INMA; and at 4-5 years of age, 20 exposures in G21 and 21 in INMA (Supplementary Table 1). Associations were evaluated considering both single-exposure and multiple-exposure effects.

#### Single-exposure analysis

In a first step, single-exposure associations were examined through an Exposome-Wide Association Study (ExWAS) stratified by sex, using ordinal logistic regression models to assess associations between individual environmental exposures and pubertal timing. All ExWAS models were stratified by sex and adjusted for key covariates (child exact age, birthweight, maternal pre-pregnancy BMI, maternal age, and household income). Statistical significance was assessed using p-values adjusted for multiple comparisons using the Benjamini–Hochberg procedure to control the false discovery rate (FDR). The correction was applied separately within each set of analyses, defined by cohort, outcome, sex stratum, and exposure time point. Thus, for each outcome, all exposure-specific associations tested within the same exposure period and sex stratum were jointly included in a single FDR adjustment, while associations from different exposure periods, sex strata, outcomes, or cohorts were adjusted independently. Although FDR correction was applied in all analyses, FDR-corrected p-values were identical to nominal p-values in some analyses, as indicated in the tables.

#### Multi-exposure analysis

Given that individuals are simultaneously exposed to different urban environmental factors that may be correlated, and that these exposures may interact and co-occur within the same context, a multi-exposure approach was considered in the present study, focusing on the urban environmental exposome. This approach aimed to assess the combined effect of urban environmental exposures, rather than analysing each exposure separately.

In a second step, to account for the effect of simultaneous exposures. an unsupervised clustering analysis was performed to identify groups of children with similar patterns of environmental exposure using Agglomerative Hierarchical Clustering (AHC), followed by association analysis between identified clusters and puberty outcomes. To reduce dimensionality, Principal Component Analysis (PCA) was applied prior to clustering, retaining components explaining 80-90% of the total variance. Clustering was conducted separately by sex, for each time point at which exposure were available and for each cohort. In INMA, exposures were standardized within each subcohort prior to PCA and clustering to reduce the influence of between-city differences on cluster assignment. Subcohort and degree of urbanization were not included as clustering variables, as the aim was to identify clusters based on environmental exposure profiles rather than predefined geographic or urbanization categories. Their distribution was subsequently examined descriptively across clusters, and sensitivity analyses excluding rural areas were performed. For consistency and interpretability, a two-cluster solution was predefined and applied across all analyses, while the Calinski–Harabasz (CH) index was used as complementary metric. As the main analysis, ordinal logistic regression models adjusting for key covariates, as described above, were fitted to evaluate the association between identified urban environmental exposure clusters and pubertal development. Following characterization of the clusters, the group with the lowest burden of environmental hazards was designated as Cluster 1 and was used as the reference category for this regression.

In a third step, the clustering procedure was repeated using AHC, focusing on urban environmental exposures grouped into subdomains (built environment, natural spaces, and traffic and air pollution) (Supplementary Table 1), to explore which subdomains might be driving the associations observed in the exposome analysis. For each subdomain, a two-cluster solution was predefined. Associations between these subdomain-specific clusters and pubertal timing were then assessed using ordinal logistic regression models, adjusted for the same covariates.

In a fourth and final step, the association between combined exclusive breastfeeding duration-environmental exposure groups and pubertal development was examined using ordinal logistic regression models, adjusted for the same set of covariates. In this analysis, exclusive breastfeeding duration was considered as a categorical variable, and environmental exposure was represented by the AHC-derived clusters. The reference group (Group 1) included participants classified in the more favourable environmental exposure cluster who had been exclusively breastfed for 24 weeks or more.

All multi-exposure analyses were adjusted for multiple testing using Benjamini-Hochberg FDR approach. As a global FDR correction strategy, for k=2 clustering analyses, all exposure-outcome associations tested across outcomes and exposure periods within the same sex stratum were jointly included in a single FDR adjustment, while associations from different sex strata were adjusted independently. Additionally, alternative block-specific FDR corrections were examined depending on the analytical approach and were defined by outcome, sex stratum, exposure period, and clustering solution. However, because only one exposure variable was tested (cluster 2, using cluster 1 as reference), FDR-corrected p-values were identical to the nominal p-values. In subdomain analyses, associations across outcomes, exposure periods, and subdomains within the same sex stratum were jointly included in a single FDR adjustment. For interaction analyses, FDR correction was applied separately within each interaction type (e.g., exposome x diet, exposome x breastfeeding) and sex stratum, jointly considering all interaction tests across exposure periods and outcomes. All correction strategies are transparently described in the corresponding tables and supplementary material.

Statistical analyses were performed using R software (version 4.4.3).

#### Sensitivity analysis

As sensitivity analyses, ordinal logistic regression models based on the AHC-derived clusters were performed excluding participants living in rural areas in both cohorts. In addition, analyses restricted to participants with normal weight were conducted in the G21 cohort only. Ordinal logistic regression models were also fitted including only individuals who remained in the same environmental exposure cluster across all available time points. Subdomain-specific analyses (built environment, natural spaces, and traffic and air pollution) were also repeated after excluding participants living in rural areas in both cohorts, as well as in analyses restricted to participants with normal weight in the G21 cohort.

### 2.7. Ethical considerations

All phases of the study complied with the Ethical Principles for Medical Research Involving Human Subjects expressed in the Declaration of Helsinki. The baseline and follow-up evaluations were approved by the University of Porto Medical School/ S. João Hospital Centre Ethics Committee, except the 13-year follow-up that was approved by the ISPUP Ethics Committee. At baseline and follow-up evaluations, all procedures were explained to participants, and an informed consent was signed by one of the parents or legal guardians (at 13 years, it was also signed by the participants). The baseline evaluation was approved by the Data Protection National Commission, and the study follows the EU General Data Protection Regulation under close supervision of the Data Protection Office of ISPUP. In the INMA cohort, written informed consent was obtained from all mothers during pregnancy and from the child’s legal guardians at the clinical examination, and the research protocol was approved by the Ethics Committees of each region.

## 3. Results

### 3.1. Study population

Figures 1 and Supplementary Figures 1 and 2 summarise the study design and analytical approach. The majority of children in both cohorts were girls (51.5% in G21 and 50.1% in INMA), with a mean age at outcome assessment of 8.7 years in INMA and 10.2 years in G21 (p<0.001) (Table 1). The prevalence of exclusive breastfeeding for ≥24 weeks was similar between sexes and did not differ significantly between cohorts.

**Figure 2.**
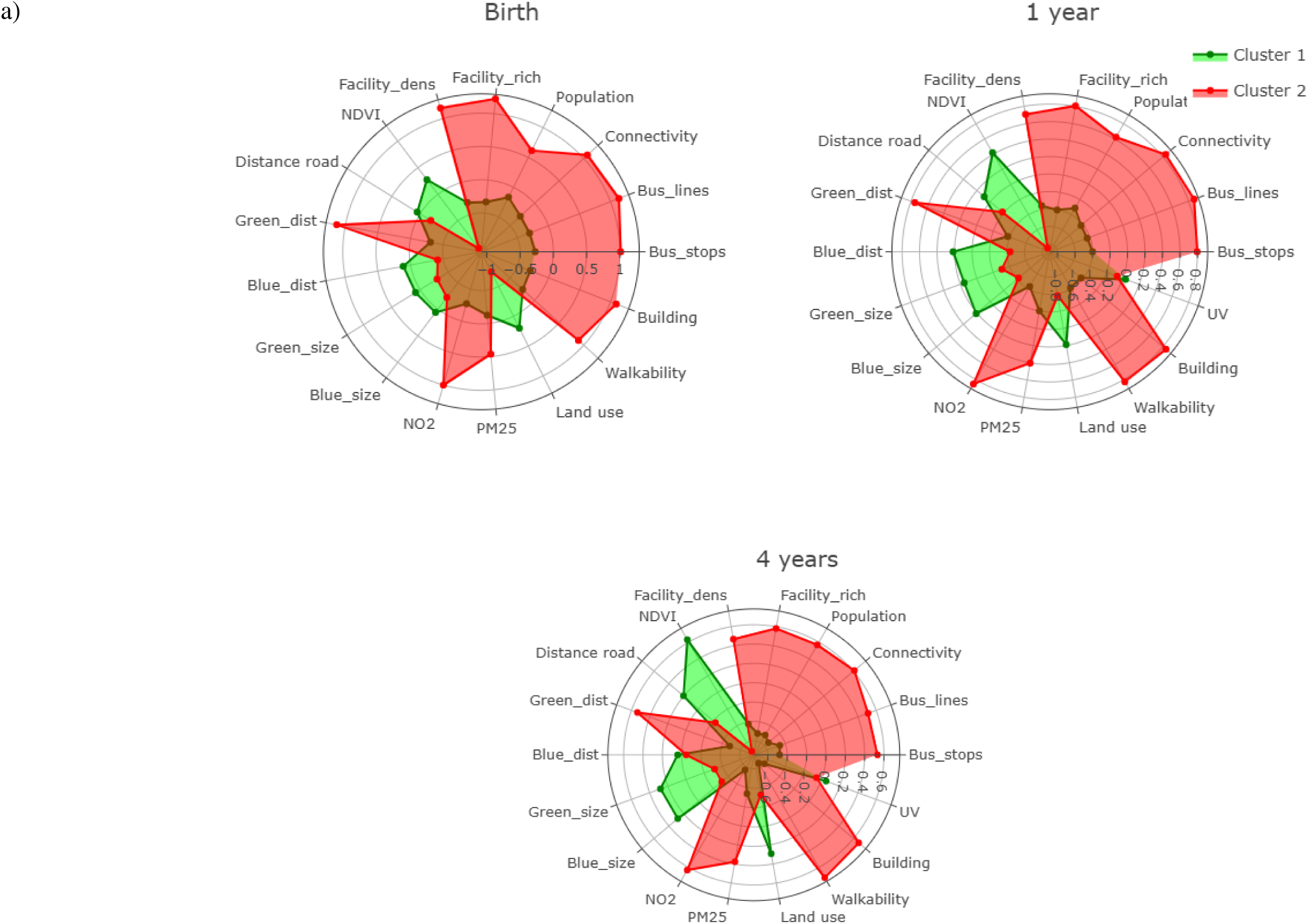
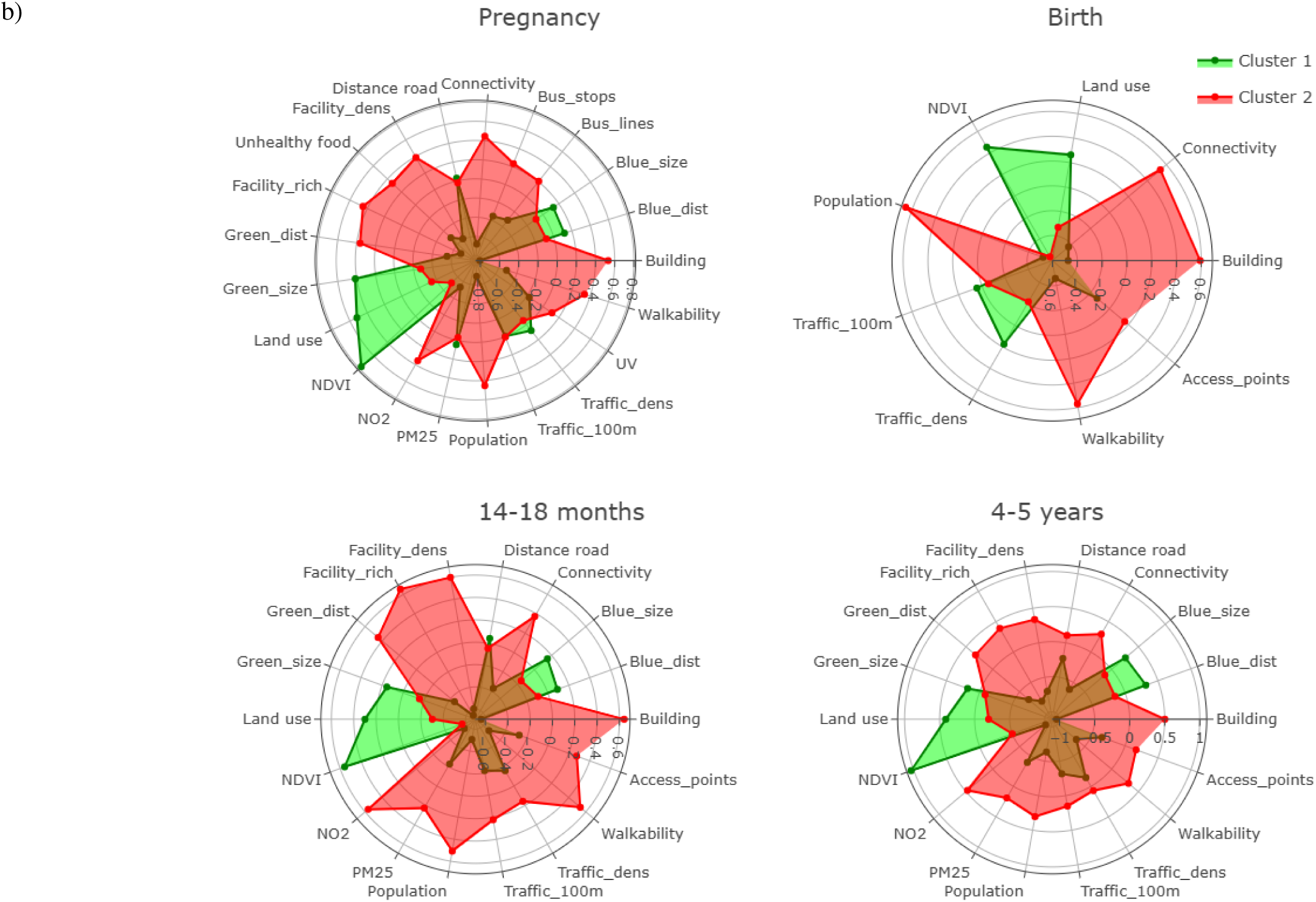
Radar plots of environmental exposure clusters across early-life time windows in the G21 and INMA cohorts. a) G21 cohort; b) INMA cohort. Green areas represent Cluster 1 and red areas represent Cluster 2. Abbreviations: Access_points,; Building, building density; Blue_dist, distance to nearest major blue space; Blue_size, size of the nearest major blue space; Green_dist, distance to nearest major green space; Green_size, size of the nearest major green space; Bus_lines, length of public transport lines; Bus_stops, density of public bus stops; Connectivity, connectivity density; Distance road, inverse distance to nearest road; Facility_dens, facility density; Facility_rich, facility richness; Unhealthy food, unhealthy food facility density; Land use, land use Shannon’s Evenness Index; NDVI, Normalized Difference Vegetation Index; NO2, nitrogen dioxide; PM25, particulate matter with an aerodynamic diameter of less than 2.5 μm; Population, population density; Traffic_100, traffic load on all roads in 100m buffer; Traffic_dens, traffic density in nearest road; UV, average of vitamin D UV dose; Walkability, walkability index; F&V, fruit and vegetables; EDF, energy-dense foods; HEI, healthy eating index; BF_any, non-exclusive breastfeeding duration; BF_exc, exclusive breastfeeding duration.

**Table 1.**
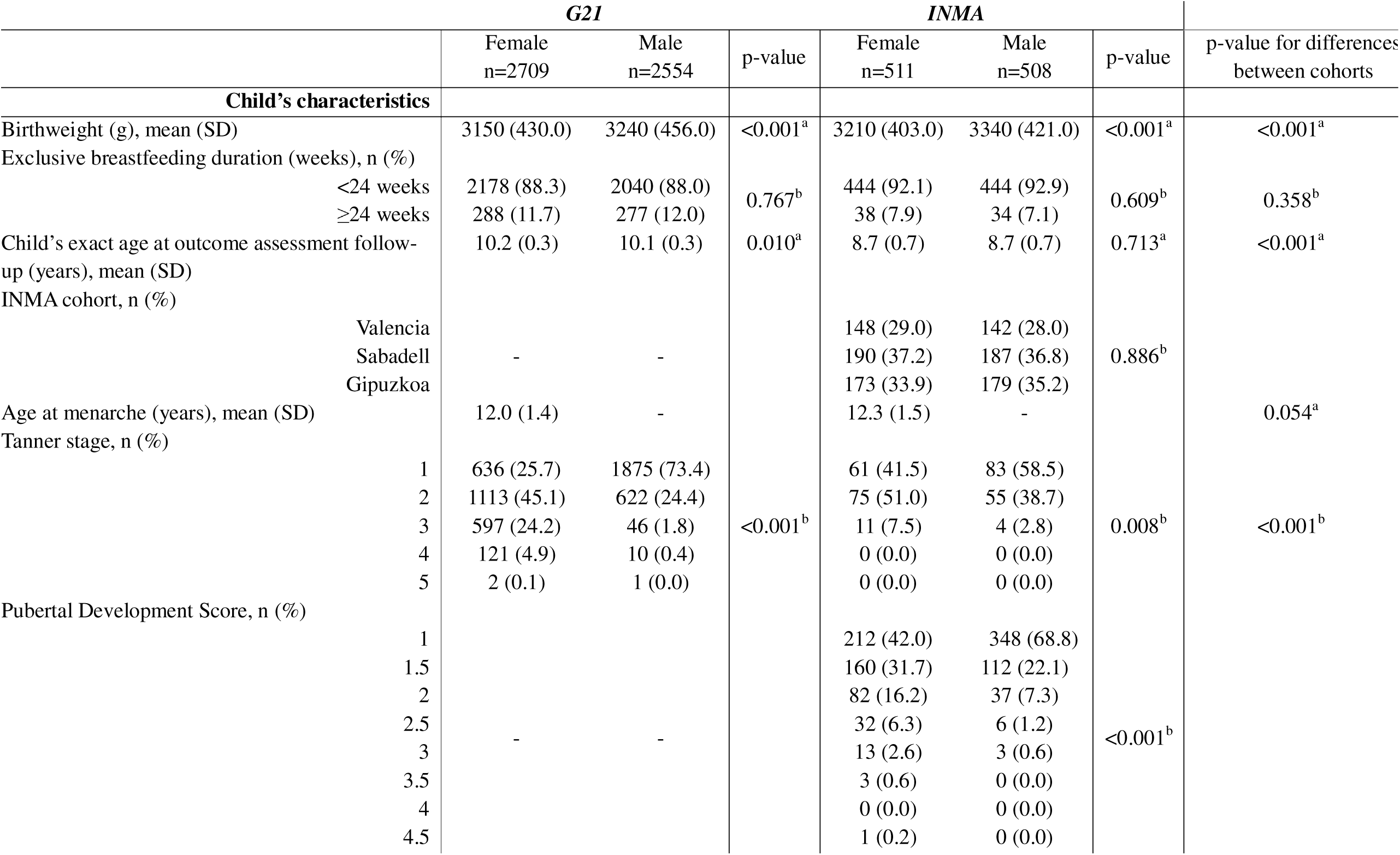
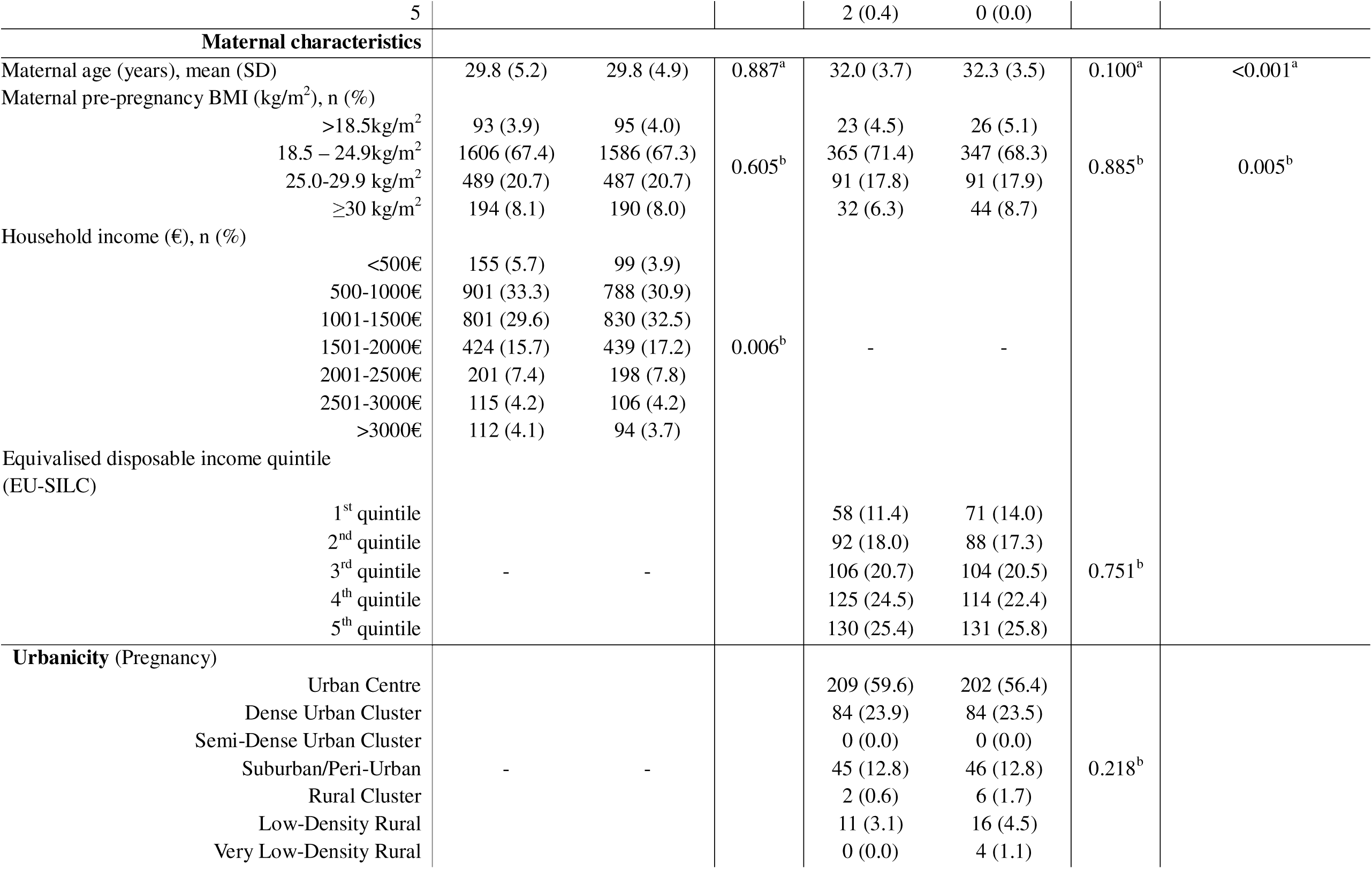
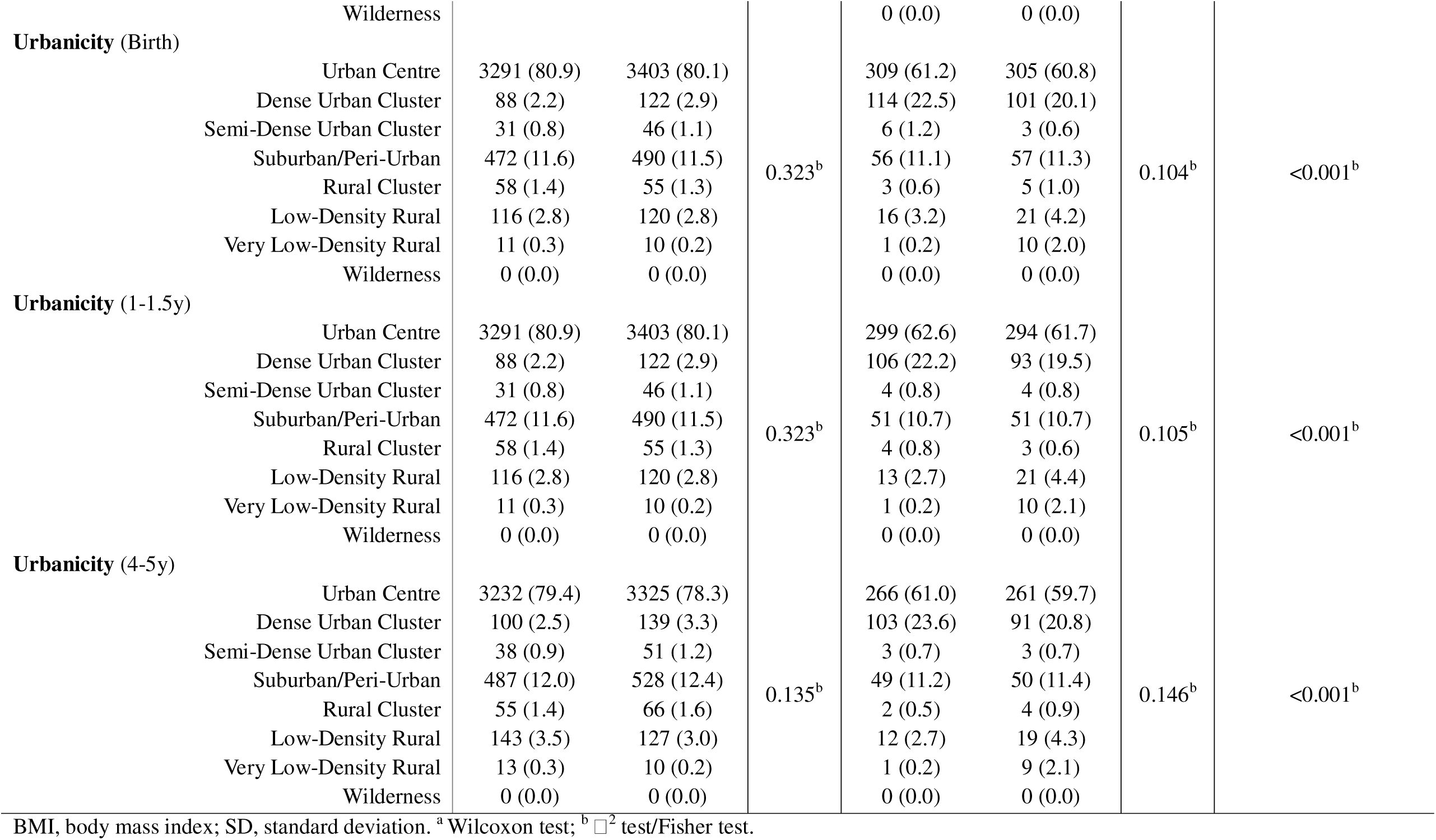
Maternal and child characteristics by sex and cohort.

In both cohorts, at the time of outcome assessment, most girls had already reached Tanner stage 2 or higher (74.3% in G21 and 58.5% in INMA), whereas the majority of boys had not yet initiated puberty, being classified as Tanner stage 1 (73.4% in G21 and 58.5% in INMA). Similarly, PDS distributions in INMA indicated more advanced development among girls compared to boys (p<0.001). The mean age at menarche was slightly lower in G21 compared to INMA, although this difference was not statistically significant. Mothers in INMA were older than those in G21 (p<0.001), and there were small differences in pre-pregnancy BMI distributions (p=0.005).

Similar clustering patterns were observed across both cohorts (Supplementary Figure 4). In general, Cluster 1 was characterised by greater availability of blue and green spaces and lower levels of urbanicity and air pollution, whereas Cluster 2 reflected more urbanised environments with higher building density, connectivity, walkability, population density, and air pollution levels. Compared with INMA, the G21 cohort generally presented higher levels of urbanicity and air pollution indicators, but also greater availability of green and blue spaces.

### 3.2. Single-exposure models for pubertal timing

ExWAS analysis has been proposed as an initial approach to study the urban environmental and lifestyle determinants associated with pubertal timing. FDR-adjusted associations between urban environmental and lifestyle exposures at birth and at 1 and 4 years of age and pubertal timing are presented in Supplementary Figure 3. Overall, significant results were only observed in the G21 cohort. In G21, a longer duration of exclusive breastfeeding was associated with a lower Tanner stage among females, indicating less advanced pubertal development, after FDR correction. Regarding age at menarche, environmental exposures at birth and at 1 year of age related to built environment factors, including higher walkability index, greater density of public bus stops, higher building and population density, and higher concentrations of air pollutants such as NOL and PML.L (at 1 year only), were negatively associated with earlier age at menarche, suggesting accelerated pubertal timing after FDR correction. Conversely, higher levels of NDVI, an indicator of access to natural spaces, were positively associated with a later age at menarche after FDR correction, indicating a protective effect against accelerated pubertal timing. At 4 years of age, exposure to built environment factors followed a similar pattern to that observed at birth and 1 year, with higher population density and walkability index being negatively associated with age at menarche after FDR correction, suggesting that greater exposure at 4 years was related to an earlier onset of menarche (Supplementary Figure 3). In both cohorts, no statistically significant associations after FDR correction were observed for diet quality variables.

### 3.3. Multi-exposure models for pubertal timing (urban environmental exposures)

To account for the combined effects of multiple correlated urban environmental exposures, we applied a clustering approach to derive environmental profiles, allowing the identification of real-life exposure patterns rather than isolated factors (Figure 2). For each exposure data point, our clustering approach revealed two urban environmental profiles. A cluster representing a more favourable or healthy environment, characterised by greater availability of green spaces, vegetation, and land-use diversity, together with lower building density, traffic intensity, and air pollution levels. On the other hand, the second cluster reflected a less favourable or unhealthy profile, involving higher building density, traffic intensity, and pollutant concentrations, as well as lower access to natural spaces. Those profiles were identified ubiquitously for each exposure follow-up and cohort (Figure 2 and Supplementary Figure 4).

Subsequently, the association between clusters at each exposure time point and pubertal outcomes, including Tanner stage, PDS score, and age at menarche was assessed. In the G21 cohort, no statistically significant associations were observed between urban environmental clusters and Tanner stage after FDR correction. When age at menarche was considered as the outcome, females in the unhealthier cluster exhibited an earlier age at menarche compared with those in the healthier cluster at exposure ages 1 (β=0-0.172, nominal p-value=0.041; FDR-adjusted p-value=0.041; global FDR-adjusted p-value=0.084) and 4 years (β=-0.181, nominal p-value=0.024; FDR-adjusted p-value=0.024; global FDR-adjusted p-value=0.084) (Supplementary Table 2 and Figure 3).

**Figure 3.**
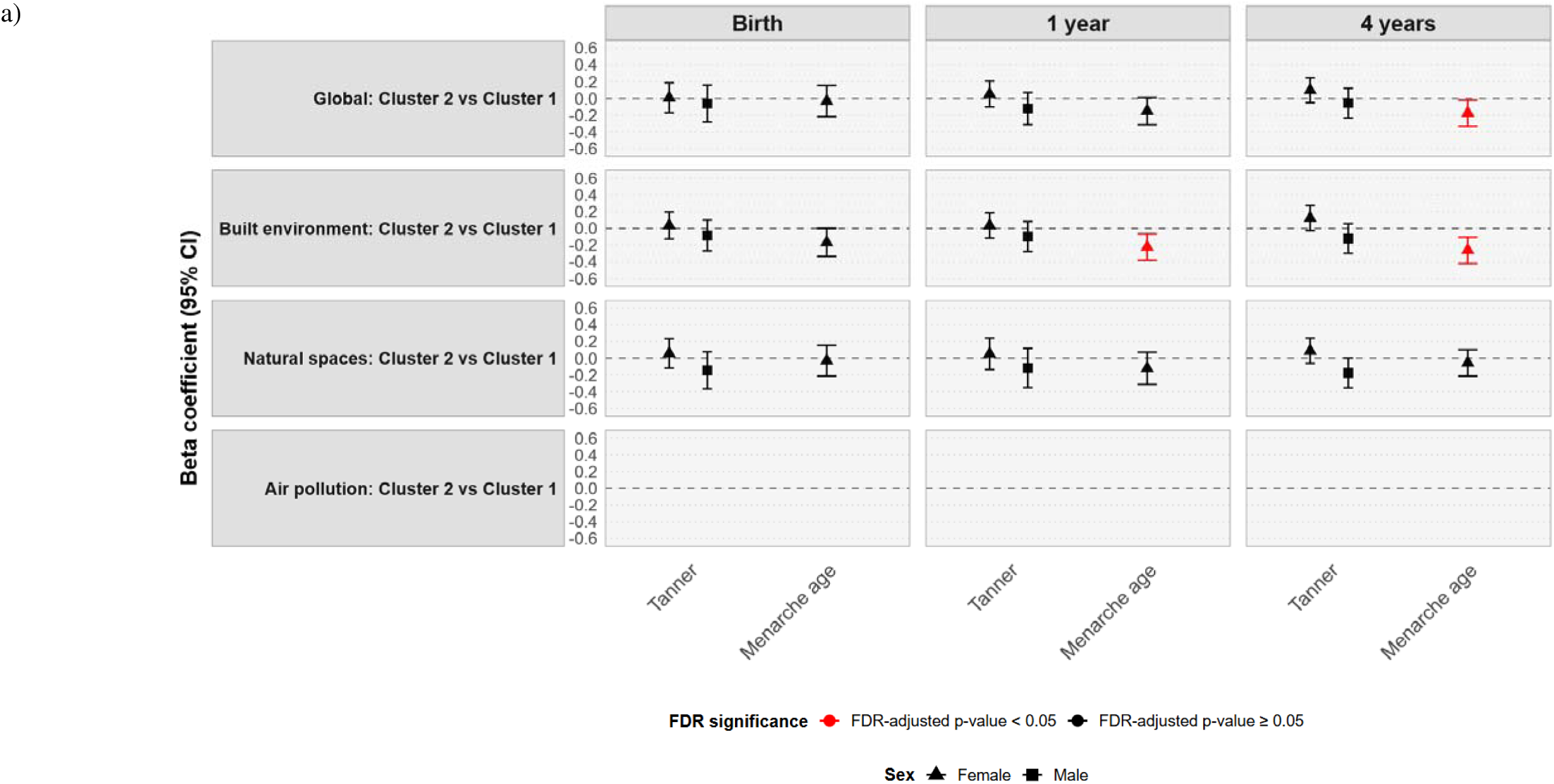
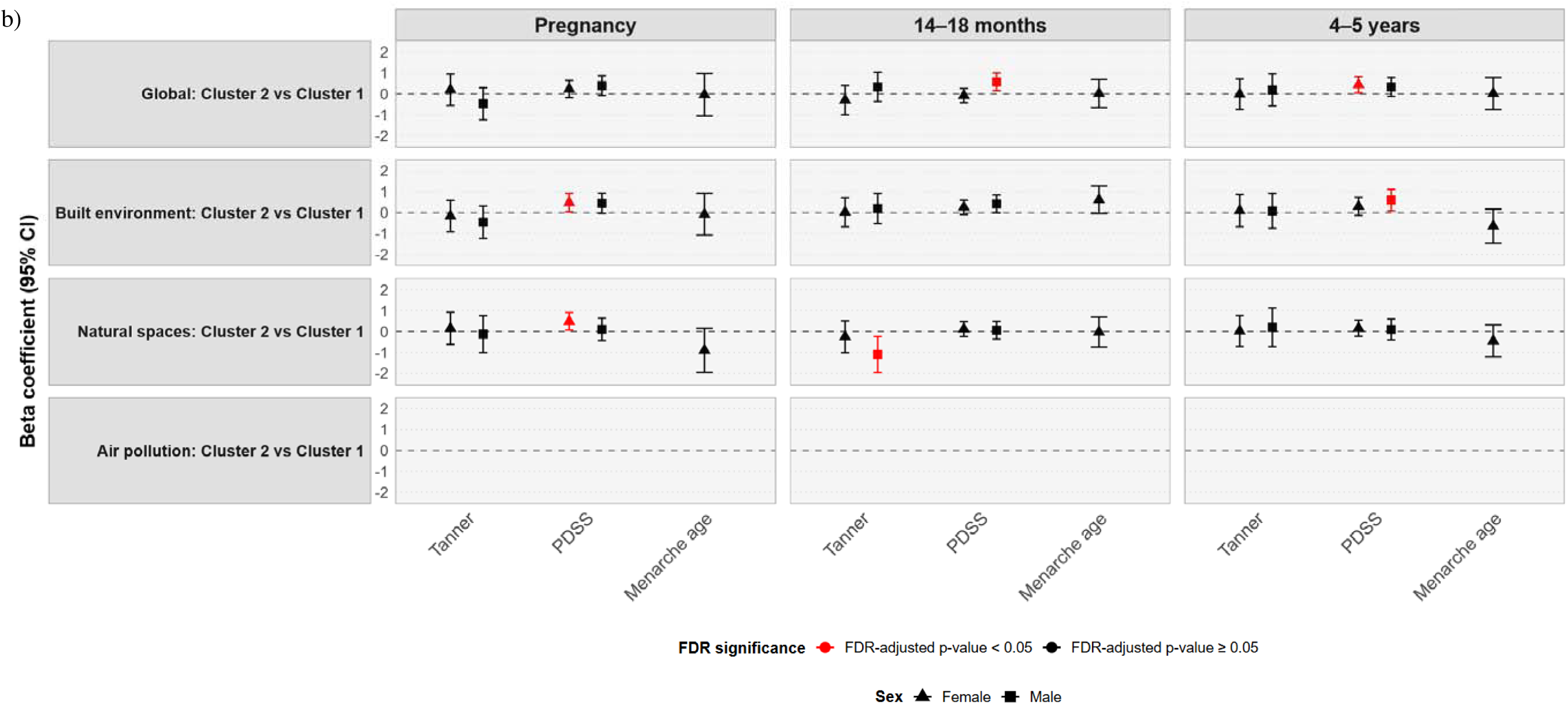
Standardized beta estimates from cluster-based ordinal regression models for pubertal outcomes across different exposure time points in the G21 and INMA cohorts. **a)** G21 cohort; b) INMA cohort. Estimates represent standardized beta coefficients comparing Cluster 2 (less favourable environmental profile) versus Cluster 1 (favourable environmental profile). Results with FDR-adjusted p-values <0.05 are highlighted in red. Results are shown for global environmental exposure and each environmental subdomain (built environment, natural spaces, and air pollution & traffic). Models are stratified by sex. In the G21 cohort (a), statistically significant associations after FDR correction were observed for the global analysis at 4 years and for the built environment subdomain at 1 and 4 years with age at menarche as the outcome. At 4 years, belonging to Cluster 2 (less favourable urban environment) was associated with a lower age at menarche. Similarly, at 1 and 4 years, belonging to Cluster 2 of the built environment subdomain was also associated with a lower age at menarche, compared with girls belonging to Cluster 1. Associations with Tanner stage as the outcome were not statistically significant after FDR correction. In the INMA cohort (b), statistically significant associations after FDR correction were observed for the global analysis and for the built environment and natural spaces subdomains. During pregnancy, belonging to Cluster 2 in the built environment and natural spaces subdomains was positively associated with PDSS scores in females. At 14–18 months, significant associations were observed for the global analysis and the natural spaces subdomain, where Cluster 2 was associated with higher PDSS scores in males and lower Tanner stages, also in males, respectively. At 4–5 years, significant positive associations were identified between Cluster 2 from the global analysis and PDSS scores in females, and between Cluster 2 from the built environment subdomain and PDSS scores in males. Associations with age at menarche were not statistically significant after FDR correction.

In the INMA cohort, no statistically significant associations were identified between urban environmental clusters and either Tanner stage or age at menarche. Regarding PDS score, males in the unhealthier cluster showed higher PDS scores than those in the healthier cluster at exposure ages 14–18 months (β=0.572, nominal p-value=0.008; FDR-adjusted p-value=0.008; global FDR-adjusted p-value=0.065) and 4–5 years (β=0.437, nominal p-value=0.027; FDR-adjusted p-value=0.027; global FDR-adjusted p-value=0.327) (Supplementary Table 2 and Figure 3).

### 3.4. Multi-exposure models by exposome subdomain for pubertal timing (urban environmental exposures)

Clustering analyses were also conducted within exposome subdomains to identify which external exposure domains are the most relevant for the pubertal timing outcome. The exposome was organized into three main subdomains according to the nature of the variables assessed: (i) Air pollution & Traffic, (ii) Natural spaces, and (iii) Built environment.

In G21, when Tanner stage was considered as the outcome, only one statistically significant association was observed at 4 years of age in the ‘Air Pollution & Traffic’ subdomain (β=0.187, nominal p-value=0.045; FDR-adjusted p-value=0.045; global FDR-adjusted p-value=0.153), with females in the unhealthier cluster presenting a higher Tanner stage compared with those in the healthier cluster, suggesting a more advanced pubertal development in the unhealthier cluster. No statistically significant associations were observed for the remaining exposure time-points or subdomains. When age at menarche was considered as the outcome, statistically significant associations were observed in the ‘Air Pollution & Traffic’ subdomain at all exposure time-points (Birth: β=-0.253, nominal p-value=0.028; FDR-adjusted p-value=0.028; global FDR-adjusted p-value=0.111; 1 year: β=-0.330, nominal p-value=0.004; FDR-adjusted p-value=0.004; global FDR-adjusted p-value=0.028; 4 years: β=-0.219, nominal p-value=0.024; FDR-adjusted p-value=0.024; global FDR-adjusted p-value=0.111), and in the ‘Built environment’ subdomain at 1 year (β=-0.224, nominal p-value=0.005; FDR-adjusted p-value=0.005; global FDR-adjusted p-value=0.032) and 4 years of age (β=-0.263, nominal p-value=0.001; FDR-adjusted p-value=0.001; global FDR-adjusted p-value=0.026). In all statistically significant associations, females in the unhealthier cluster presented a lower age at menarche compared with those in the healthier cluster, consistent with accelerated pubertal timing (Supplementary Table 3).

In INMA, considering Tanner stage as the outcome, a statistically significant association was observed only at the 1-1.5-year time-point. In the ‘Natural spaces’ subdomain, males in the unhealthier cluster, characterised by lower access to natural spaces compared with the healthier cluster, showed a higher Tanner stage (β=1.100, nominal p-value=0.012; FDR-adjusted p-value=0.012; global FDR-adjusted p-value=0.213), indicating more advanced pubertal development in this group. No statistically significant associations were observed for age at menarche at any exposure time-point or subdomain. When PDS was considered as the outcome, for exposure during pregnancy, females in the unhealthier cluster of the ‘Built environment’ (β=0.479, nominal p-value=0.031; FDR-adjusted p-value=0.031; global FDR-adjusted p-value=0.294) and ‘Natural spaces’ (β=0.486, nominal p-value=0.024; FDR-adjusted p-value=0.024; global FDR-adjusted p-value=0.294) subdomains presented higher PDS scores compared with females in the healthier cluster, also suggesting more advanced pubertal development. At 1-1.5 years and at 4-5 years of age, females in the unhealthier cluster of the ‘Air Pollution & Traffic’ subdomain also presented higher PDS scores than females in the healthier cluster (1-1.5 years: β=0.380, nominal p-value=0.033; FDR-adjusted p-value=0.033; global FDR-adjusted p-value=0.294; 4-5 years: β=0.381, nominal p-value=0.044; FDR-adjusted p-value=0.044; global FDR-adjusted p-value=0.294). In addition, at 4-5 years of age, males in unhealthier cluster of the ‘Built environment’ subdomain also presented higher PDS scores compared with males in healthier cluster (β=0.599, nominal p-value=0.024; FDR-adjusted p-value=0.024; global FDR-adjusted p-value=0.213), consistent with more advanced pubertal development in males in the unhealthier cluster (Supplementary Table 3).

### 3.5. Multi-exposure models for pubertal timing and interaction with exclusive breastfeeding duration

To explore the potential combined effects of early-life nutrition and urban environmental exposures, a joint analysis was conducted integrating exclusive breastfeeding duration (<24 weeks vs. ≥24 weeks) with the previously defined urban environmental exposure clusters (healthier vs. unhealthier clusters). This approach aimed to assess how environmental exposures interact with early-life nutritional factors and to determine which one of these factors plays a more important role in pubertal timing.

In the combined analysis of exclusive breastfeeding duration and urban environmental exposure clusters, no statistically significant associations were observed on pubertal development after FDR correction for any outcome, exposure time point, or cohort (Supplementary Table 4).

### 3.6. Sensitivity analysis

In the sensitivity analysis restricted to the urban population (Supplementary Table 5), no FDR-statistically significant associations were observed in the G21 cohort. In the INMA cohort, a significant association was observed only among boys for exposures measured at 1-1.5 years, with boys in the unhealthier cluster presenting higher PDS scores than those in the healthier cluster (β=0.550, nominal p-value=0.014; FDR-adjusted p-value=0.014; global FDR-adjusted p-value=0.111), indicating more advanced pubertal development in the unhealthier cluster. In the sensitivity analysis restricted to participants with normal weight in G21 (Supplementary Table 6), no significant associations were observed for any outcome or exposure time point. In the sensitivity analysis restricted to individuals who remained in the same cluster over time (Supplementary Table 7), no significant associations were observed in G21, while in INMA, boys showed higher PDS scores in the unhealthier cluster, at all exposure time points (Pregnancy: β=1.013, nominal p-value=0.024; FDR-adjusted p-value=0.024; global FDR-adjusted p-value=0.048; Birth: β=1.014, nominal p-value=0.024; FDR-adjusted p-value=0.024; global FDR-adjusted p-value=0.048; 1-1.5 years: β=1.015, nominal p-value=0.024; FDR-adjusted p-value=0.024; global FDR-adjusted p-value=0.048; 4-5 years: β=1.016, nominal p-value=0.024; FDR-adjusted p-value=0.024; global FDR-adjusted p-value=0.048).

In the subdomain-specific sensitivity analysis restricted to the urban population (Supplementary Table 8), G21 females in unhealthier cluster of the ‘Built environment’ subdomain had a lower age at menarche at exposure time-points 1 (β=-0.190, nominal p-value=0.022; FDR-adjusted p-value=0.046; global FDR-adjusted p-value=0.142) and 4 years of age (β=-0.239, nominal p-value=0.004; FDR-adjusted p-value=0.012; global FDR-adjusted p-value=0.071), indicating earlier menarche compared with females in the healthier cluster. In both time points, females in unhealthier cluster of the ‘Built environment’ subdomain had a lower age at menarche compared with females in the healthier cluster. At the 1-year exposure time point, the unhealthier cluster of the ‘Air Pollution & Traffic’ subdomain was also associated with a lower age at menarche (β= −0.282, nominal p-value=0.031; FDR-adjusted p-value=0.046; global FDR-adjusted p-value=0.142), consistent with earlier pubertal timing.

In the sensitivity analysis restricted to participants with normal weight in G21 (Supplementary Table 9), subdomain analyses showed statistically significant associations at 4 years of age in the ‘Natural spaces’ (β=0.222, nominal p-value=0.032; FDR-adjusted p-value=0.047; global FDR-adjusted p-value=0.189) and ‘Air Pollution & Traffic’ (β=0.278, nominal p-value=0.028; FDR-adjusted p-value=0.047; global FDR-adjusted p-value=0.189) subdomains, with girls in unhealthier cluster presenting a higher Tanner stage compared with girls in the healthier cluster, indicating more advanced pubertal development. At the 1-year exposure time point, girls in the unhealthier of the Built environment subdomain had a lower age at menarche (β=-0.266, nominal p-value=0.016; FDR-adjusted p-value=0.047; global FDR-adjusted p-value=0.189), suggesting earlier pubertal timing.

## 4. Discussion

This study provides evidence that living in an unfavourable urban environment, characterized by higher building and population density, traffic and air pollution, and lower access to natural spaces, is associated with an earlier age at menarche in Portuguese girls and with a more advanced pubertal stage among Spanish boys. To our knowledge, this is the first study to examine the effect of multiple early-life urban environmental exposures measured at repeated time points across early-life and under an exposome approach on pubertal timing. All urban environmental exposome subdomains evaluated in our study, namely ‘Built environment’, ‘Natural spaces’, and ‘Air Pollution & Traffic’, were independently associated with pubertal development or age at menarche in the two cohorts. In particular, the ‘Air Pollution & Traffic’ subdomain appeared to be the most consistently associated with pubertal development across almost all exposure time points, especially among girls. Additionally, the ExWAS analysis, reported how a longer duration of exclusive breastfeeding was also associated with a lower Tanner stage at 10 years of age among females in the G21 Portuguese cohort. However, interaction analyses did not provide evidence of synergistic or combined effect between exclusive breastfeeding duration and environmental exposures on pubertal timing. Similarly, we found no evidence that exclusive breastfeeding duration attenuated the potential adverse effects of unhealthier urban environmental exposures on pubertal timing. Importantly, under the more conservative global FDR correction approach, statistically significant associations were substantially reduced and remained only in the subdomain analyses within the G21 cohort, specifically for age at menarche in relation to the Air pollution subdomain at 1–1.5 years and the Built environment subdomain at 1–1.5 and 4–5 years of age.

The present study corroborates previous evidence indicating that a longer duration of breastfeeding, in this case exclusive breastfeeding, is associated with a later age at menarche (42, 44, 71, 72). It is possible that childhood body weight status mediates the relationship between breastfeeding and pubertal timing, as longer breastfeeding duration appears to be protective against childhood obesity (73), and obesity, in turn, has been associated with earlier pubertal timing (74). However, some studies have also reported no association between weight status and earlier pubertal onset (75, 76). An alternative pathway may be that breastfeeding is associated with other maternal and family behaviours considered healthier, including higher diet quality during childhood (77) and better sleep habits (78), factors that have also been linked to pubertal development (50, 79). This study therefore supports the potential value of investing in breastfeeding policies as a public health intervention with potential long-term benefits for child pubertal development.

Regarding urban environmental exposures, air pollution and traffic showed the most consistently associations with pubertal timing across cohorts, with associations observed for age at menarche in the G21 cohort and PDS score in girls in the INMA cohort, suggesting a potential role of these exposures in pubertal development. Previous studies have also reported associations between exposure to air pollution and traffic-related factors and pubertal development (25, 80–84). For example, a prospective cohort study conducted in the United States reported that girls with higher in utero and childhood exposure to air pollution were more likely to experience an earlier age at menarche (85). Several biological mechanisms may underlie these associations. Air pollution derived from industrial and vehicle emissions contains multiple compounds, including PM and NO, as well as persistent organic pollutants and polycyclic aromatic hydrocarbons (PAHs), which may act as endocrine-disrupting chemicals or have endocrine-disrupting properties (86). These pollutants can interfere with hormonal pathways that are key regulators of reproductive development, and may activate the aryl hydrocarbon, androgen, and estrogen receptors (86, 87). PM has been suggested to influence estrogen signalling, which may stimulate the release of kisspeptin and subsequently trigger gonadotropin-releasing hormone secretion, a central regulator of pubertal onset (88). In addition, exposure to PM may exert obesogenic effects. Previous studies have shown that higher prenatal and postnatal exposure to PM2.5 is associated with higher BMI z-scores and an increased risk of childhood obesity (89, 90). Increased adiposity, in turn, has long been recognised as an important trigger of pubertal onset in girls (74). In general, all these findings highlight that air pollution and traffic exposure may have broader implications beyond respiratory health, influencing key stages of child development. Notably, in our sensitivity analysis restricted to participants with normal weight, the previously observed subdomain associations were no longer statistically significant in the majority of cases, suggesting that child BMI may play an important role in the relationship between urban environmental exposures and pubertal timing, potentially acting as a mediator. However, the reduced sample size in these restricted analyses may also have limited statistical power.

Access to natural spaces also appears to play an important role in pubertal development in both boys and girls in the INMA cohort. Only few studies have investigated the influence of access to natural spaces on pubertal timing (91, 92). One study found no association between living in areas with more green spaces and age at menarche (92), whereas another study showed that African American girls who lived in neighbourhoods with greater availability of parks, walking or hiking trails, playing fields, and basketball or tennis courts experienced lower rates of onset of breast and pubic hair development over four years of follow-up (91). To our knowledge, our study is the first to suggest a potential protective association between natural spaces and pubertal timing in both sexes. Natural spaces, including green and blue spaces, may promote higher levels of physical activity and reduce sedentary behaviours (33, 93). Moreover, increased exposure to nature also seems to be associated with reduced stress levels, as nature induces a response that reduce cortisol levels (94). In turn, early-life stress appears to act as a potential trigger for menarche (32).

Finally, we also observed an association between the ‘Built Environment’ subdomain and pubertal timing, including age at menarche. This subdomain may influence pubertal timing more indirectly, as neighbourhoods with higher population and built density are often associated with fewer opportunities for physical activity (95). In addition, less favourable built environments may also be linked to higher exposure to air pollution (96).

In the sensitivity analysis restricted to the urban population, most associations observed in the main analysis were no longer statistically significant, particularly in the G21 cohort. This suggests that the observed associations may be driven by differences between rural and urban environments, rather than contrasts within urban settings. Nonetheless, the association that remained in the INMA cohort among boys at 1-1.5 years, with higher PDS scores in the unhealthier cluster, suggests that this effect may be less dependent on the urban-rural context for evaluated Spanish cities.

Based on data from these two Iberian cohorts, pubertal timing in Southern European populations appears to follow similar patterns to those reported in other European settings and worldwide (97). In our study, exposure profiles characterised by higher air pollution and lower access to green spaces were consistently associated with more advanced pubertal development and a younger age at menarche, suggesting that the quality of the urban environment may play a relevant role in shaping pubertal development. Compared with other European regions, Portugal and Spain are generally characterised by intermediate levels of air pollution, higher than those observed in Northern Europe but lower than those reported in parts of Eastern Europe (98). However, access to green spaces shows marked intra-urban heterogeneity, particularly in densely built-up areas. In major cities such as Barcelona, large urban parks exist but are unevenly distributed across neighbourhoods (99). These findings highlight the importance of incorporating child health considerations into urban environmental policies, with a particular focus on reducing traffic-related emissions and promoting equitable access to green infrastructure.

One of the main strengths of this study is the use of data from two longitudinal cohorts, based on relatively large population-based samples with high participation rates, in which some of the findings were replicated across cohorts. However, several limitations should be considered. First, some measurement error may arise from the assessment of Tanner stages, particularly from the measurement of testicular volume in males, even when following standardized guidelines and using an orchidometer, as this is a sensitive and highly invasive procedure. Another limitation is that most children in the INMA cohort were assessed using the PDS rather than Tanner stages, which is considered the gold standard. Although the PDS does not provide a complete estimation of Tanner stages, it has been previously validated, including in the INMA-Valencia subcohort, showing moderate to good agreement with physician-assessed Tanner stages (100, 101). In addition, a substantial proportion of missing data on child BMI in the INMA cohort (∼37% missingness) could not be imputed and, therefore, BMI could not be considered as an adjustment variable in the present study. Nevertheless, a sensitivity analysis restricted to normal-weight children was conducted to address this limitation. Furthermore, recall bias may affect breastfeeding information, as it was collected only at specific time points. Finally, maternal and child dietary information was based on self-reported data, which may be subject to misreporting and social desirability bias.

## 5. Conclusion

This study highlights that exposures to the urban environment, from pregnancy through childhood, may influence pubertal timing in both boys and girls. Air pollution and traffic appear to be the determinants most consistently associated with pubertal timing. Duration of exclusive breastfeeding also seems to act as a protective factor against early onset of menarche. No statistically significant associations were observed in the interaction analysis between exclusive breastfeeding duration and urban environment clusters, thereby limiting the determination of which factor plays a more prominent role in pubertal timing, and suggesting that these factors may operate through independent pathways rather than synergistic effects. These results reinforce the importance of the first years of life as a sensitive window for environmental influences on development. Improving the urban environment through policies that explicitly consider child health, and implementing strategies that support longer breastfeeding duration, may represent complementary public health strategies to reduce the risk of earlier puberal development in both sexes.

## Data Availability

All data produced in the present study are available upon reasonable request to the authors

