## Supplementary Tables for "Early-life Urban Environment, Nutrition, and Pubertal Timing in Southern Europe: An Exposome Analysis": Supplementary material.docx

**Section I. Exposure assessment methods**

In this section, we describe the methods used to estimate all exposures included in the exposome for Generation XXI and INMA cohorts. A detailed list of all exposure variables included in the analyses is provided in **Table 1**.

1.1. Air pollution and traffic

Exposure to outdoor air pollution, including nitrogen dioxide (NO₂) and particulate matter (PM), was estimated using environmental monitoring data combined with land-use regression models. These models incorporated geographical information on land use, traffic indicators, and altitude (61,62), and were used to predict individual-level residential exposure.

1.2. Natural spaces

Residential greenness was assessed using the Normalized Difference Vegetation Index (NDVI) within a 300m buffer around each participant’s residence. NDVI is a satellite-based indicator of vegetation cover, with values ranging from -1 to 1, where higher values indicate greater photosynthetically active vegetation. Satellite images were selected for each study area during peak greenness periods under cloud-free conditions, and NDVI was estimated at the time of delivery (63). Access to green and blue spaces was measured as the straight-line distance from the participant’s residence to the nearest green space and blue space, respectively, using data from an urban atlas database (63).

1.3. Built environment

Built environment indicators were derived from multiple spatial data sources. Traffic data and public transport accessibility were obtained from municipal records, while population density was derived from the Spanish Statistical Office (INE). Street connectivity and facility locations were obtained from digital street maps provided by Navteq, and building density data were retrieved from regional cartographic institutes (63). Street connectivity was defined as the number of intersections per square kilometre within 300m buffers. Facilities were characterized using two complementary indicators: (i) a facility richness index, defined as the number of different facility types divided by the maximum possible number of facility types within the 300-m buffer; and (ii) a facility density index, calculated as the number of facilities per unit area within the 300m buffer.

Land-use characteristics were summarized using a land-use mix indicator based on Shannon’s Evenness Index, reflecting the diversity of land-use categories within the area. Traffic exposure was characterised using three indicators: total traffic load within a 100m buffer, traffic density on the nearest road, and the inverse distance to the nearest road. Accessibility to public transport was assessed using two measures within 300m buffers using two measures: the total length of public bus lines and the number of bus stops. The food environment was defined as the density of unhealthy food outlets, calculated as the number of such facilities divided by the area of the 300m buffer, and assessed for the first trimester of pregnancy. Finally, a walkability index was constructed by combining population density, street connectivity, facility richness, and land-use evenness. The index was calculated using both the mean and the sum of the deciles of these components within 300m buffers, resulting in a walkability score ranging from 0 to 1 (63).

**Section II. Supplementary tables**

**Supplementary Table 1.** Environmental exposures considered at each time point, by cohort.

|  | Pregnancy | Birth | 1-1.5y | 4-5y | 9-10y | 13/18y |
| --- | --- | --- | --- | --- | --- | --- |
| INMA |  |  |  |  |  |  |
| Air pollution | NO_2_ | - | NO_2_ | NO_2_ | - | - |
|  | PM_2.5_ |  | PM_2.5_ | PM_2.5_ |  |  |
| Built environment | Building density | Building density | Building density | Building density | - | - |
|  | Population density | Population density | Population density | Population density |  |  |
|  | Lenght of public transport lines | Number of accesspoints | Number of accesspoints | Number of accesspoints |  |  |
|  | Density of public bus stops | Connectivity density | Connectivity density | Connectivity density |  |  |
|  | Connectivity density | Land use Shannon’s Evenness Index | Land use Shannon’s Evenness Index | Land use Shannon’s Evenness Index |  |  |
|  | Facility density | Walkability index | Facility density | Facility density |  |  |
|  | Facility richness |  | Facility richness | Facility richness |  |  |
|  | Unhealthy food facility density |  | Walkability index | Walkability index |  |  |
|  | Land use Shannon’s Evenness Index |  |  |  |  |  |
|  | Walkability index |  |  |  |  |  |
| Natural spaces | Distance to nearest major blue space | NDVI | Distance to nearest major blue space | Distance to nearest major blue space | - | - |
|  | Size of nearest major blue space |  | Size of nearest major blue space | Size of nearest major blue space |  |  |
|  | Distance to nearest major green space |  | Distance to nearest major green space | Distance to nearest major green space |  |  |
|  | Size of nearest major green space |  | Size of nearest major green space | Size of nearest major green space |  |  |
|  | Normalized Difference Vegetation Index (NDVI) |  | NDVI | NDVI |  |  |
| Traffic | Inverse distance to nearest road | Traffic load on all roads in 100m buffer | Inverse distance to nearest road | Inverse distance to nearest road | - | - |
|  | Traffic load on all roads in 100m buffer | Traffic density in nearest road | Traffic load on all roads in 100m buffer | Traffic load on all roads in 100m buffer |  |  |
|  | Traffic density in nearest road |  | Traffic density in nearest road | Traffic density in nearest road |  |  |
| Lifestyle | Energy-dense foods (EDF) | - | Total breastfeeding duration | EDF | - | - |
|  | Fruit and vegetables (FV) |  | Exclusive breastfeeding duration | FV |  |  |
|  | Healthy Eating Index (HEI) |  |  | HEI |  |  |
| Outcome assessment | - | - | - | - | Pubertal Development Scale | - |
|  |  |  |  |  | Tanner stages (Valencia) |  |
|  |  |  |  |  | Age at menarche (Sabadell) |  |
| G21 |  |  |  |  |  |  |
| Air pollution | - | NO_2_ | NO_2_ | NO_2_ | - | - |
|  |  | PM_2.5_ | PM_2.5_ | PM_2.5_ |  |  |
| Built environment | - | Building density | Building density | Building density | - | - |
|  |  | Population density | Population density | Population density |  |  |
|  |  | Lenght of public transport lines | Lenght of public transport lines | Lenght of public transport lines |  |  |
|  |  | Density of public bus stops | Density of public bus stops | Density of public bus stops |  |  |
|  |  | Connectivity density | Connectivity density | Connectivity density |  |  |
|  |  | Facility density | Facility density | Facility density |  |  |
|  |  | Facility richness | Facility richness | Facility richness |  |  |
|  |  | Land use Shannon’s Evenness Index | Land use Shannon’s Evenness Index | Land use Shannon’s Evenness Index |  |  |
|  |  | Walkability index | Walkability index | Walkability index |  |  |
| Natural spaces | - | Distance to nearest major blue space | Distance to nearest major blue space | Distance to nearest major blue space | - | - |
|  |  | Size of nearest major blue space | Size of nearest major blue space | Size of nearest major blue space |  |  |
|  |  | Distance to nearest major green space | Distance to nearest major green space | Distance to nearest major green space |  |  |
|  |  | Size of nearest major green space | Size of nearest major green space | Size of nearest major green space |  |  |
|  |  | NDVI | NDVI | NDVI |  |  |
| Traffic | - | Inverse distance to nearest road | Inverse distance to nearest road | Inverse distance to nearest road | - | - |
| Lifestyle | - | - | Total breastfeeding duration | EDF | - | - |
|  |  |  | Exclusive breastfeeding duration | FV |  |  |
|  |  |  |  | HEI |  |  |
| Outcome assessment |  |  |  |  | Tanner stages | Age at menarche |

NO_2,_ nitrogen dioxide; PM_2.5_, particulate matter with an aerodynamic diameter of less than 2.5 μm; NDVI, Normalized Difference Vegetation Index; EDF, Energy-dense foods; FV, Fruit and vegetables; HEI, Healthy Eating Index.

**Supplementary Table 2**. Associations between early-life environmental exposure clusters and pubertal timing in the G21 and INMA cohorts.

|  | G21 | | | | INMA | | | | | |
| --- | --- | --- | --- | --- | --- | --- | --- | --- | --- | --- |
|  | **Tanner stage** | | **Menarche age** | | **Tanner stage** | | **Menarche age** | | **PDS** | |
|  | **β** | **p-value** | **β** | **p-value** | **β** | **p-value** | **β** | **p-value** | **β** | **p-value** |
| Pregnancy |  | |  | |  |  |  |  |  |  |
| Female |  | |  | |  |  |  |  |  |  |
| Cluster 1 | - | | - | | Ref. | | Ref. | | Ref. | |
| Cluster 2 |  |  |  |  | 0.192 | 0.616 | -0.041 | 0.937 | 0.231 | 0.279 |
| Male |  | |  | |  |  |  |  |  |  |
| Cluster 1 | - | | **-** | | Ref. | | - | | Ref. | |
| Cluster 2 |  |  |  |  | -0.473 | 0.228 |  |  | 0.391 | 0.102 |
| Birth |  |  |  |  |  |  |  |  |  |  |
| Female |  |  |  |  |  |  |  |  |  |  |
| Cluster 1 | Ref. | | Ref. | | Ref. | | Ref. | | Ref. | |
| Cluster 2 | 0.019 | 0.835 | -0.053 | 0.579 | -0.137 | 0.699 | 0.082 | 0.806 | 0.129 | 0.449 |
| Male |  |  |  |  |  |  |  |  |  |  |
| Cluster 1 | Ref. | | **-** | | Ref. | | **-** | | Ref. | |
| Cluster 2 | -0.071 | 0.535 |  |  | 0.112 | 0.752 |  |  | 0.282 | 0.168 |
| 1-1.5 years |  | |  | |  |  |  |  |  |  |
| Female |  | |  | |  |  |  |  |  |  |
| Cluster 1 | Ref. | | Ref. | | Ref. | | Ref. | | Ref. | |
| Cluster 2 | 0.060 | 0.454 | **-0.172** | **0.041** | -0.299 | 0.402 | 0.019 | 0.957 | -0.086 | 0.623 |
| Male |  | |  | |  |  |  |  |  |  |
| Cluster 1 | Ref. | | - | | Ref. | | - | | Ref. | |
| Cluster 2 | -0.122 | 0.216 |  |  | 0.332 | 0.354 |  |  | **0.572** | **0.008** |
| 4-5 years |  |  |  |  |  |  |  |  |  |  |
| Female |  |  |  |  |  |  |  |  |  |  |
| Cluster 1 | Ref. | | Ref. | | Ref. | | Ref. | | Ref. | |
| Cluster 2 | 0.099 | 0.191 | **-0.181** | **0.024** | -0.014 | 0.970 | 0.016 | 0.968 | 0.333 | 0.150 |
| Male |  |  |  |  |  |  |  |  |  |  |
| Cluster 1 | Ref. | | - | | Ref. | | - | | Ref. | |
| Cluster 2 | -0.059 | 0.517 |  |  | 0.188 | 0.630 |  |  | **0.437** | **0.027** |

Cluster 1 represents a more favourable or healthier environmental profile, characterised by greater availability of green spaces, vegetation and land-use diversity, and lower building density, traffic intensity, and air pollution levels. Cluster 2 represents a less favourable or unhealthier environmental profile, characterised by higher building density, traffic intensity, and pollutant concentrations, and lower access to natural spaces. Cluster 1 was used as the reference category in the present analysis. P-values are FDR adjusted using the Benjamini-Hochberg procedure. FDR correction was applied separately by cohort, sex, outcome, and exposure time point. In all analysis, FDR-corrected p-values were identical to the nominal p-values.

**Supplementary Table 3.** Associations between clusters of environmental subdomains (natural spaces, built environment, and air pollution and traffic) and pubertal timing indicators.

|  |  | G21 | | | | | INMA | | | | | | | |
| --- | --- | --- | --- | --- | --- | --- | --- | --- | --- | --- | --- | --- | --- | --- |
|  |  | **Tanner stage** | | **Menarche age** | | | **Tanner stage** | | **Menarche age** | | | | **PDS** | |
|  |  | **β** | **p-value** | **β** | | **p-value** | **β** | **p-value** | **β** | | **p-value** | | **β** | **p-value** |
| Pregnancy |  |  | |  | | |  |  |  | |  | |  |  |
| Built environment | Female |  | |  | | |  |  |  | |  | |  |  |
|  | Cluster 1 | - | | - | | | Ref. | | Ref. | | | | Ref. | |
|  | Cluster 2 |  |  |  |  |  | -0.164 | 0.671 | -0.079 | | 0.877 | | **0.479** | **0.031** |
|  | Male |  | |  | | |  |  | - | | | |  |  |
|  | Cluster 1 | - | | **-** | | | Ref. | |  |  |  |  | Ref. | |
|  | Cluster 2 |  |  |  |  |  | -0.461 | 0.248 |  |  |  |  | 0.446 | 0.070 |
| Natural spaces | Female |  | |  | | |  |  |  | | | |  |  |
|  | Cluster 1 | - | | **-** | | | Ref. | | Ref. | | | | Ref. | |
|  | Cluster 2 |  |  |  |  |  | 0.150 | 0.703 | -0.900 | | | 0.093 | **0.486** | **0.024** |
|  | Male |  | |  | | |  |  | **-** | | | |  |  |
|  | Cluster 1 | - | | **-** | | | Ref. | |  |  |  |  | Ref. | |
|  | Cluster 2 |  |  |  |  |  | -0.126 | 0.779 |  |  |  |  | 0.104 | 0.704 |
| Air pollution & traffic | Female |  | |  | | |  |  |  | | | |  |  |
|  | Cluster 1 | - | | **-** | | | Ref. | | Ref. | | | | Ref. | |
|  | Cluster 2 |  |  |  |  |  | -0.923 | 0.133 | -0.451 | | | 0.435 | 0.161 | 0.525 |
|  | Male |  | |  | | |  |  | **-** | | | |  |  |
|  | Cluster 1 | - | | **-** | | | Ref. | |  |  |  |  | Ref. | |
|  | Cluster 2 |  |  |  |  |  | 0.305 | 0.596 |  |  |  |  | 0.086 | 0.773 |
| Birth |  |  |  |  | |  |  |  |  | |  | |  |  |
| Built environment | Female |  |  |  | |  |  |  |  | |  | |  |  |
|  | Cluster 1 | Ref. | | Ref. | | | - | | - | | | | - | |
|  | Cluster 2 | 0.035 | 0.671 | -0.168 | | 0.051 |  |  |  |  |  |  |  |  |
|  | Male |  |  | **-** | | |  |  |  | |  | |  |  |
|  | Cluster 1 | Ref. | |  |  |  | **-** | | **-** | | | | - | |
|  | Cluster 2 | -0.085 | 0.369 |  |  |  |  |  |  |  |  |  |  |  |
| Natural spaces | Female |  |  |  | | |  |  |  | | | |  |  |
|  | Cluster 1 | Ref. | | Ref. | | | - | | **-** | | | | **-** | |
|  | Cluster 2 | 0.057 | 0.524 | -0.029 | 0.755 | |  |  |  |  |  |  |  |  |
|  | Male |  |  | - | | |  |  |  | | | |  |  |
|  | Cluster 1 | Ref. | |  |  |  | - | | **-** | | | | **-** | |
|  | Cluster 2 | -0.145 | 0.200 |  |  |  |  |  |  |  |  |  |  |  |
| Air pollution & traffic | Female |  |  |  | | |  |  |  | | | |  |  |
|  | Cluster 1 | Ref. | | Ref. | | | - | | **-** | | | | **-** | |
|  | Cluster 2 | 0.014 | 0.895 | **-0.253** | **0.028** | |  |  |  |  |  |  |  |  |
|  | Male |  |  | - | | |  |  |  | | | |  |  |
|  | Cluster 1 | Ref. | |  |  |  | - | | **-** | | | | **-** | |
|  | Cluster 2 | -0.031 | 0.813 |  |  |  |  |  |  |  |  |  |  |  |
| 1-1.5 years |  |  | |  | | |  |  |  | |  | |  |  |
| Built environment | Female |  | |  | | |  |  |  | |  | |  |  |
|  | Cluster 1 | Ref. | | Ref. | | | Ref. | | Ref. | | | | Ref. | |
|  | Cluster 2 | 0.034 | 0.657 | **-0.224** | | **0.005** | 0.023 | 0.949 | 0.621 | | 0.065 | | 0.262 | 0.133 |
|  | Male |  | | - | | |  |  | - | | | |  |  |
|  | Cluster 1 | Ref. | |  |  |  | Ref. | |  |  |  |  | Ref. | |
|  | Cluster 2 | -0.098 | 0.284 |  |  |  | 0.198 | 0.585 |  |  |  |  | 0.418 | 0.051 |
| Natural spaces | Female |  |  |  | | |  |  |  | | | |  |  |
|  | Cluster 1 | Ref. | | Ref. | | | Ref. | | Ref. | | | | Ref. | |
|  | Cluster 2 | 0.052 | 0.585 | -0.121 | 0.219 | | 0.251 | 0.516 | 0.028 | | | 0.939 | -0.115 | 0.528 |
|  | Male |  |  | - | | |  |  | - | | | |  |  |
|  | Cluster 1 | Ref. | |  |  |  | Ref. | |  |  |  |  | Ref. | |
|  | Cluster 2 | -0.117 | 0.330 |  |  |  | **1.100** | **0.012** |  |  |  |  | -0.059 | 0.784 |
| Air pollution & traffic | Female |  |  |  | | |  |  |  | | | |  |  |
|  | Cluster 1 | Ref. | | Ref. | | | Ref. | | Ref. | | | | Ref. | |
|  | Cluster 2 | 0.041 | 0.702 | **-0.330** | **0.004** | | -0.549 | 0.148 | -0.303 | | | 0.393 | **0.380** | **0.033** |
|  | Male |  |  | - | | |  |  | - | | | |  |  |
|  | Cluster 1 | Ref. | |  |  |  | Ref. | |  |  |  |  | Ref. | |
|  | Cluster 2 | -0.040 | 0.750 |  |  |  | 0.101 | 0.792 |  |  |  |  | 0.197 | 0.354 |
| 4-5 years |  |  |  |  | |  |  |  |  | |  | |  |  |
| Built environment | Female |  |  |  | |  |  |  |  | |  | |  |  |
|  | Cluster 1 | Ref. | | Ref. | | | Ref. | | Ref. | | | | Ref. | |
|  | Cluster 2 | 0.123 | 0.106 | **-0.263** | | **0.001** | 0.096 | 0.807 | -0.646 | | 0.121 | | 0.303 | 0.170 |
|  | Male |  |  | - | | |  |  | - | | | |  |  |
|  | Cluster 1 | Ref. | |  |  |  | Ref. | |  |  |  |  | Ref. | |
|  | Cluster 2 | -0.122 | 0.183 |  |  |  | 0.083 | 0.846 |  |  |  |  | **0.599** | **0.024** |
| Natural spaces | Female |  |  |  | | |  |  |  | | | |  |  |
|  | Cluster 1 | Ref. | | Ref. | | | Ref. | | Ref. | | | | Ref. | |
|  | Cluster 2 | 0.088 | 0.246 | -0.055 | 0.490 | | 0.023 | 0.951 | -0.452 | 0.248 | | | 0.158 | 0.426 |
|  | Male |  |  | - | | |  |  | - | | | |  |  |
|  | Cluster 1 | Ref. | |  |  |  | Ref. | |  |  |  |  | Ref. | |
|  | Cluster 2 | -0.176 | 0.052 |  |  |  | 0.201 | 0.670 |  |  |  |  | 0.098 | 0.699 |
| Air pollution & traffic | Female |  |  |  | | |  |  |  | | | |  |  |
|  | Cluster 1 | Ref. | | Ref. | | | Ref. | | Ref. | | | | Ref. | |
|  | Cluster 2 | **0.187** | **0.045** | **-0.219** | **0.024** | | -0.186 | 0.601 | -0.351 | 0.368 | | | **0.381** | **0.044** |
|  | Male |  |  | - | | |  |  | - | | | |  |  |
|  | Cluster 1 | Ref. | |  |  |  | Ref. | |  |  |  |  | Ref. | |
|  | Cluster 2 | -0.058 | 0.597 |  |  |  | 0.479 | 0.203 |  |  |  |  | 0.334 | 0.131 |

Cluster 1 represents a more favourable or healthier environmental profile, characterised by greater availability of green spaces, vegetation and land-use diversity, and lower building density, traffic intensity, and air pollution levels. Cluster 2 represents a less favourable or unhealthier environmental profile, characterised by higher building density, traffic intensity, and pollutant concentrations, and lower access to natural spaces. Cluster 1 was used as the reference category in the present analysis. P-values are FDR adjusted using the Benjamini-Hochberg procedure. FDR correction was applied separately by cohort, sex, outcome, exposure time point, and urban environmental subdomain. In all analysis, FDR-corrected p-values were identical to the nominal p-values.

**Supplementary Table 4**. Ordinal logistic regression models of pubertal development by combined exclusive breastfeeding duration and environmental exposure clusters.

|  | G21 | | | | | INMA | | | |
| --- | --- | --- | --- | --- | --- | --- | --- | --- | --- |
|  | **Tanner stage** | | | **Menarche age** | | **Menarche age** | | **PDS** | |
|  | **β** | **p-value** | | **β** | **p-value** | **β** | **p-value** | **β** | **p-value** |
| Pregnancy |  | | |  | |  |  |  |  |
| Female |  | | |  | |  |  |  |  |
| Group 1 | Ref. | | | Ref. | | Ref. | | Ref. | |
| Group 2 |  | | |  | | 0.970 | 0.611 | 0.114 | 0.923 |
| Group 3 |  | | |  | | -1.528 | 0.611 | 0.536 | 0.923 |
| Group 4 |  | | |  | | 0.919 | 0.611 | 0.364 | 0.923 |
| Male |  | | |  | |  |  |  |  |
| Group 1 | Ref. | | | **-** | | - | | Ref. | |
| Group 2 |  | | |  |  |  |  | -0.935 | 0.379 |
| Group 3 |  | | |  |  |  |  | -0.750 | 0.630 |
| Group 4 |  | | |  |  |  |  | -0.486 | 0.630 |
| Birth |  |  | |  |  |  |  |  |  |
| Female |  |  | |  |  |  |  |  |  |
| Group 1 | Ref. | | | Ref. | | Ref. | | Ref. | |
| Group 2 | 0.173 | 0.506 | | -0.184 | 0.447 | 0.253 | 0.742 | -0.174 | 0.923 |
| Group 3 | -0.042 | 0.884^#^ | | -0.494 | 0.447 | -1.462 | 0.611 | -0.442 | 0.923 |
| Group 4 | 0.217 | 0.506 | | -0.163 | 0.573 | 0.510 | 0.611 | -0.076 | 0.923 |
| Male |  |  | |  |  |  |  |  |  |
| Group 1 | Ref. | | | **-** | | **-** | | Ref. | |
| Group 2 | -0.326 | | 0.056 |  |  |  |  | -0.809 | 0.379 |
| Group 3 | -0.720 | | 0.056 |  |  |  |  | -1.384 | 0.379 |
| Group 4 | -0.395 | | 0.056 |  |  |  |  | -0.824 | 0.379 |
| 1-1.5 years |  | | |  | |  |  |  |  |
| Female |  | | |  | |  |  |  |  |
| Group 1 | Ref. | | | Ref. | | Ref. | | Ref. | |
| Group 2 | 0.106 | 0.762 | | -0.044 | 0.856 | -0.584 | 0.611 | 0.224 | 0.923 |
| Group 3 | -0.154 | 0.762 | | -0.063 | 0.856 | -2.638 | 0.127 | 0.237 | 0.923 |
| Group 4 | 0.193 | 0.506 | | -0.226 | 0.447 | -0.194 | 0.781^#^ | 0.042 | 0.923^#^ |
| Male |  | | |  | |  |  |  |  |
| Group 1 | Ref. | | | - | | - | | Ref. | |
| Group 2 | -0.269 | | 0.134 |  |  |  |  | -0.445 | 0.630 |
| Group 3 | -0.369 | | 0.204^#^ |  |  |  |  | -0.352 | 0.751 |
| Group 4 | -0.413 | | 0.056 |  |  |  |  | -0.149 | 0.817 |
| 4-5 years |  |  | |  |  |  |  |  |  |
| Female |  |  | |  |  |  |  |  |  |
| Group 1 | Ref. | | | Ref. | | Ref. | | Ref. | |
| Group 2 | -0.071 | | 0.762 | -0.033 | 0.856^#^ | -0.565 | 0.646 | -0.188 | 0.923 |
| Group 3 | -0.376 | | 0.506 | -0.097 | 0.856 | -1.376 | 0.611 | 0.122 | 0.923 |
| Group 4 | 0.085 | | 0.762 | -0.246 | 0.447 | -0.338 | 0.742 | 0.248 | 0.923 |
| Male |  |  | |  |  |  |  |  |  |
| Group 1 | Ref. | | | - | | - | | Ref. | |
| Group 2 | -0.409 | | 0.056 |  |  |  |  | -0.574 | 0.630 |
| Group 3 | -0.590 | | 0.056 |  |  |  |  | -0.186 | 0.817^#^ |
| Group 4 | -0.460 | | 0.056 |  |  |  |  | -0.338 | 0.751 |

Four groups were defined according to combinations of urban environmental exposure clusters and exclusive breastfeeding duration (<24 weeks vs. ≥24 weeks): Group 1 (favourable environmental profile and ≥24 weeks of exclusive breastfeeding; reference category), Group 2 (favourable environmental profile and <24 weeks), Group 3 (unfavourable environmental profile and ≥24 weeks), and Group 4 (unfavourable environmental profile and <24 weeks). P-values are FDR adjusted using the Benjamini-Hochberg procedure. FDR correction was applied separately within each interaction type (e.g., exposome x diet, exposome x breastfeeding) and sex stratum. ^#^FDR-corrected p-values were identical to the nominal p-values.

**Supplementary Table 5.** Associations between early-life environmental exposure clusters and pubertal timing in the G21 and INMA cohorts, restricted to the urban population.

|  | G21 | | | | INMA | | | | | |
| --- | --- | --- | --- | --- | --- | --- | --- | --- | --- | --- |
|  | **Tanner stage** | | **Menarche age** | | **Tanner stage** | | **Menarche age** | | **PDS** | |
|  | **β** | **p-value** | **β** | **p-value** | **β** | **p-value** | **β** | **p-value** | **β** | **p-value** |
| Pregnancy |  | |  | |  |  |  |  |  |  |
| Female |  | |  | |  |  |  |  |  |  |
| Cluster 1 | - | | - | | Ref. | | Ref. | | Ref. | |
| Cluster 2 |  |  |  |  | 0.192 | 0.616 | -0.041 | 0.937 | 0.142 | 0.521 |
| Male |  | |  | |  |  |  |  |  |  |
| Cluster 1 | - | | **-** | | Ref. | | - | | Ref. | |
| Cluster 2 |  |  |  |  | -0.492 | 0.215 |  |  | 0.299 | 0.234 |
| Birth |  |  |  |  |  |  |  |  |  |  |
| Female |  |  |  |  |  |  |  |  |  |  |
| Cluster 1 | Ref. | | Ref. | | Ref. | | Ref. | | Ref. | |
| Cluster | 0.013 | 0.891 | -0.018 | 0.856 | -0.185 | 0.604 | 0.082 | 0.806 | 0.062 | 0.719 |
| Male |  |  |  |  |  |  |  |  |  |  |
| Cluster 1 | Ref. | | **-** | | Ref. | | **-** | | Ref. | |
| Cluster 2 | -0.054 | 0.641 |  |  | 0.055 | 0.878 |  |  | 0.213 | 0.314 |
| 1-1.5 years |  | |  | |  |  |  |  |  |  |
| Female |  | |  | |  |  |  |  |  |  |
| Cluster 1 | Ref. | | Ref. | | Ref. | | Ref. | | Ref. | |
| Cluster 2 | 0.055 | 0.494 | -0.126 | 0.139 | -0.316 | 0.378 | 0.072 | 0.835 | -0.176 | 0.325 |
| Male |  | |  | |  |  |  |  |  |  |
| Cluster 1 | Ref. | | - | | Ref. | | - | | Ref. | |
| Cluster 2 | -0.099 | 0.323 |  |  | 0.260 | 0.471 |  |  | **0.550** | **0.014** |
| 4-5 years |  |  |  |  |  |  |  |  |  |  |
| Female |  |  |  |  |  |  |  |  |  |  |
| Cluster 1 | Ref. | | Ref. | | Ref. | | Ref. | | Ref. | |
| Cluster 2 | 0.107 | 0.170 | -0.143 | 0.083 | -0.087 | 0.820 | 0.016 | 0.968 | 0.335 | 0.111 |
| Male |  |  |  |  |  |  |  |  |  |  |
| Cluster 1 | Ref. | | - | | Ref. | | - | | Ref. | |
| Cluster 2 | -0.031 | 0.738 |  |  | 0.121 | 0.764 |  |  | 0.308 | 0.243 |

Cluster 1 represents a more favourable or healthier environmental profile, characterised by greater availability of green spaces, vegetation and land-use diversity, and lower building density, traffic intensity, and air pollution levels. Cluster 2 represents a less favourable or unhealthier environmental profile, characterised by higher building density, traffic intensity, and pollutant concentrations, and lower access to natural spaces. Cluster 1 was used as the reference category in the present analysis. P-values are FDR adjusted using the Benjamini-Hochberg procedure. FDR correction was applied separately by cohort, sex, outcome, and exposure time point. In all analysis, FDR-corrected p-values were identical to the nominal p-values.

**Supplementary Table 6.** Associations between early-life environmental exposure clusters and pubertal timing in the G21 cohorts, restricted to normal weight.

|  | G21 | | | |
| --- | --- | --- | --- | --- |
|  | **Tanner stage** | | **Menarche age** | |
|  | **β** | **p-value** | **β** | **p-value** |
| Birth |  |  |  |  |
| Female |  |  |  |  |
| Cluster 1 | Ref. | | Ref. | |
| Cluster 2 | 0.078 | 0.527 | -0.090 | 0.487 |
| Male |  |  |  |  |
| Cluster 1 | Ref. | | **-** | |
| Cluster 2 | 0.077 | 0.618 |  |  |
| 1-1.5 years |  | |  | |
| Female |  | |  | |
| Cluster 1 | Ref. | | Ref. | |
| Cluster 2 | 0.166 | 0.122 | -0.174 | 0.128 |
| Male |  | |  | |
| Cluster 1 | Ref. | | - | |
| Cluster 2 | -0.094 | 0.491 |  |  |
| 4-5 years |  |  |  |  |
| Female |  |  |  |  |
| Cluster 1 | Ref. | | Ref. | |
| Cluster 2 | 0.103 | 0.315 | -0.117 | 0.285 |
| Male |  |  |  |  |
| Cluster 1 | Ref. | | - | |
| Cluster 2 | 0.090 | 0.478 |  |  |

Cluster 1 represents a more favourable or healthier environmental profile, characterised by greater availability of green spaces, vegetation and land-use diversity, and lower building density, traffic intensity, and air pollution levels. Cluster 2 represents a less favourable or unhealthier environmental profile, characterised by higher building density, traffic intensity, and pollutant concentrations, and lower access to natural spaces. Cluster 1 was used as the reference category in the present analysis. P-values are FDR adjusted using the Benjamini-Hochberg procedure. FDR correction was applied separately by cohort, sex, outcome, and exposure time point. In all analysis, FDR-corrected p-values were identical to the nominal p-values.

**Supplementary Table 7.** Associations between early-life environmental exposure clusters and pubertal timing in the G21 and INMA cohorts of individuals always in the same cluster.

|  | G21 | | | | INMA | | | | | |
| --- | --- | --- | --- | --- | --- | --- | --- | --- | --- | --- |
|  | **Tanner stage** | | **Menarche age** | | **Tanner stage** | | **Menarche age** | | **PDS** | |
|  | **β** | **95% CI** | **β** | **95% CI** | **β** | **95% CI** | **β** | **95% CI** | **β** | **95% CI** |
| Pregnancy |  | |  | |  |  |  |  |  |  |
| Female |  | |  | |  |  |  |  |  |  |
| Cluster 1 | - | | - | | Ref. | | Ref. | | Ref. | |
| Cluster 2 |  |  |  |  | -0.470 | 0.394 | 0.607 | 0.476 | 0.580 | 0.100 |
| Male |  | |  | |  |  |  |  |  |  |
| Cluster 1 | - | | **-** | | Ref. | | - | | Ref. | |
| Cluster 2 |  |  |  |  | 0.232 | 0.662 |  |  | **1.013** | **0.024** |
| Birth |  |  |  |  |  |  |  |  |  |  |
| Female |  |  |  |  |  |  |  |  |  |  |
| Cluster 1 | Ref. | | Ref. | | Ref. | | Ref. | | Ref. | |
| Cluster | 0.104 | 0.454 | -0.159 | 0.286 | -0.470 | 0.394 | 0.608 | 0.476 | 0.578 | 0.101 |
| Male |  |  |  |  |  |  |  |  |  |  |
| Cluster 1 | Ref. | | **-** | | Ref. | | **-** | | Ref. | |
| Cluster 2 | 0.137 | 0.442 |  |  | 0.232 | 0.662 |  |  | **1.014** | **0.024** |
| 1-1.5 years |  | |  | |  |  |  |  |  |  |
| Female |  | |  | |  |  |  |  |  |  |
| Cluster 1 | Ref. | | Ref. | | Ref. | | Ref. | | Ref. | |
| Cluster 2 | 0.104 | 0.454 | -0.159 | 0.286 | -0.470 | 0.394 | 0.607 | 0.476 | 0.579 | 0.101 |
| Male |  | |  | |  |  |  |  |  |  |
| Cluster 1 | Ref. | | - | | Ref. | | - | | Ref. | |
| Cluster 2 | 0.137 | 0.442 |  |  | 0.232 | 0.662 |  |  | **1.015** | **0.024** |
| 4-5 years |  |  |  |  |  |  |  |  |  |  |
| Female |  |  |  |  |  |  |  |  |  |  |
| Cluster 1 | Ref. | | Ref. | | Ref. | | Ref. | | Ref. | |
| Cluster 2 | 0.104 | 0.454 | -0.159 | 0.286 | -0.470 | 0.394 | 0.608 | 0.476 | 0.577 | 0.102 |
| Male |  |  |  |  |  |  |  |  |  |  |
| Cluster 1 | Ref. | | - | | Ref. | | - | | Ref. | |
| Cluster 2 | 0.137 | 0.442 |  |  | 0.232 | 0.662 |  |  | **1.016** | **0.024** |

Cluster 1 represents a more favourable or healthier environmental profile, characterised by greater availability of green spaces, vegetation and land-use diversity, and lower building density, traffic intensity, and air pollution levels. Cluster 2 represents a less favourable or unhealthier environmental profile, characterised by higher building density, traffic intensity, and pollutant concentrations, and lower access to natural spaces. Cluster 1 was used as the reference category in the present analysis. P-values are FDR adjusted using the Benjamini-Hochberg procedure. FDR correction was applied separately by cohort, sex, outcome, and exposure time point. In all analysis, FDR-corrected p-values were identical to the nominal p-values.

**Supplementary Table 8.** Associations between clusters of environmental subdomains (natural spaces, built environment, and air pollution and traffic) and pubertal timing indicators, restricted to the urban population.

|  |  | G21 | | | | | INMA | | | | | | | |
| --- | --- | --- | --- | --- | --- | --- | --- | --- | --- | --- | --- | --- | --- | --- |
|  |  | **Tanner stage** | | **Menarche age** | | | **Tanner stage** | | **Menarche age** | | | | **PDS** | |
|  |  | **β** | **95% CI** | **β** | | **95% CI** | **β** | **95% CI** | **β** | | **95% CI** | | **β** | **95% CI** |
| Pregnancy |  |  | |  | | |  |  |  | |  | |  |  |
| Built environment | Female |  | |  | | |  |  |  | |  | |  |  |
|  | Cluster 1 | - | | - | | | Ref. | | Ref. | | | | Ref. | |
|  | Cluster 2 |  |  |  |  |  | -0.164 | 0.703 | -0.079 | | 0.877^#^ | | 0.407 | 0.118 |
|  | Male |  | |  | | |  |  | - | | | |  |  |
|  | Cluster 1 | - | | **-** | | | Ref. | |  |  |  |  | Ref. | |
|  | Cluster 2 |  |  |  |  |  | -0.477 | 0.707 |  |  |  |  | 0.354 | 0.536 |
| Natural spaces | Female |  | |  | | |  |  |  | | | |  |  |
|  | Cluster 1 | - | | **-** | | | Ref. | | Ref. | | | | Ref. | |
|  | Cluster 2 |  |  |  |  |  | 0.150 | 0.703^#^ | -0.900 | | | 0.280 | 0.444 | 0.118 |
|  | Male |  | |  | | |  |  | **-** | | | |  |  |
|  | Cluster 1 | - | | **-** | | | Ref. | |  |  |  |  | Ref. | |
|  | Cluster 2 |  |  |  |  |  | -0.144 | 0.748^#^ |  |  |  |  | 0.053 | 0.953 |
| Air pollution & Traffic | Female |  | |  | | |  |  |  | | | |  |  |
|  | Cluster 1 | - | | **-** | | | Ref. | | Ref. | | | | Ref. | |
|  | Cluster 2 |  |  |  |  |  | -0.923 | 0.398 | -0.451 | | | 0.652 | 0.081 | 0.753^#^ |
|  | Male |  | |  | | |  |  | **-** | | | |  |  |
|  | Cluster 1 | - | | **-** | | | Ref. | |  |  |  |  | Ref. | |
|  | Cluster 2 |  |  |  |  |  | 0.299 | 0.748 |  |  |  |  | 0.018 | 0.953^#^ |
| Birth |  |  |  |  | |  |  |  |  | |  | |  |  |
| Built environment | Female |  |  |  | |  |  |  |  | |  | |  |  |
|  | Cluster 1 | Ref. | | Ref. | | | - | | - | | | | - | |
|  | Cluster 2 | 0.028 | 0.741^#^ | -0.108 | | 0.353 |  |  |  |  |  |  |  |  |
|  | Male |  |  | **-** | | |  |  |  | |  | |  |  |
|  | Cluster 1 | Ref. | |  |  |  | **-** | | **-** | | | | - | |
|  | Cluster 2 | -0.040 | 0.915 |  |  |  |  |  |  |  |  |  |  |  |
| Natural spaces | Female |  |  |  | | |  |  |  | | | |  |  |
|  | Cluster 1 | Ref. | | Ref. | | | - | | **-** | | | | **-** | |
|  | Cluster 2 | 0.051 | 0.741 | 0.005 | 0.960^#^ | |  |  |  |  |  |  |  |  |
|  | Male |  |  | - | | |  |  |  | | | |  |  |
|  | Cluster 1 | Ref. | |  |  |  | - | | **-** | | | | **-** | |
|  | Cluster 2 | -0.129 | 0.779 |  |  |  |  |  |  |  |  |  |  |  |
| Air pollution & Traffic | Female |  |  |  | | |  |  |  | | | |  |  |
|  | Cluster 1 | Ref. | | Ref. | | | - | | **-** | | | | **-** | |
|  | Cluster 2 | 0.080 | 0.741 | -0.207 | 0.346 | |  |  |  |  |  |  |  |  |
|  | Male |  |  | - | | |  |  |  | | | |  |  |
|  | Cluster 1 | Ref. | |  |  |  | - | | **-** | | | | **-** | |
|  | Cluster 2 | 0.016 | 0.915^#^ |  |  |  |  |  |  |  |  |  |  |  |
| 1-1.5 years |  |  | |  | | |  |  |  | |  | |  |  |
| Built environment | Female |  | |  | | |  |  |  | |  | |  |  |
|  | Cluster 1 | Ref. | | Ref. | | | Ref. | | Ref. | | | | Ref. | |
|  | Cluster 2 | 0.029 | 0.711^#^ | **-0.190** | | **0.046** | 0.013 | 0.971^#^ | 0.700 | | 0.118 | | 0.179 | 0.319^#^ |
|  | Male |  | | - | | |  |  | - | | | |  |  |
|  | Cluster 1 | Ref. | |  |  |  | Ref. | |  |  |  |  | Ref. | |
|  | Cluster 2 | -0.059 | 0.799 |  |  |  | 0.108 | 0.849 |  |  |  |  | 0.392 | 0.262 |
| Natural spaces | Female |  |  |  | | |  |  |  | | | |  |  |
|  | Cluster 1 | Ref. | | Ref. | | | Ref. | | Ref. | | | | Ref. | |
|  | Cluster 2 | 0.047 | 0.711 | -0.088 | 0.372^#^ | | 0.304 | 0.659 | 0.070 | | | 0.849^#^ | -0.258 | 0.310 |
|  | Male |  |  | - | | |  |  | - | | | |  |  |
|  | Cluster 1 | Ref. | |  |  |  | Ref. | |  |  |  |  | Ref. | |
|  | Cluster 2 | -0.101 | 0.799 |  |  |  | 1.047 | 0.052 |  |  |  |  | -0.035 | 0.881^#^ |
| Air pollution & Traffic | Female |  |  |  | | |  |  |  | | | |  |  |
|  | Cluster 1 | Ref. | | Ref. | | | Ref. | | Ref. | | | | Ref. | |
|  | Cluster 2 | 0.057 | 0.711 | **-0.282** | **0.046** | | -0.563 | 0.424 | -0.370 | | | 0.450 | 0.233 | 0.310 |
|  | Male |  |  | - | | |  |  | - | | | |  |  |
|  | Cluster 1 | Ref. | |  |  |  | Ref. | |  |  |  |  | Ref. | |
|  | Cluster 2 | 0.011 | 0.937^#^ |  |  |  | 0.073 | 0.849^#^ |  |  |  |  | 0.161 | 0.707 |
| 4-5 years |  |  |  |  | |  |  |  |  | |  | |  |  |
| Built environment | Female |  |  |  | |  |  |  |  | |  | |  |  |
|  | Cluster 1 | Ref. | | Ref. | | | Ref. | | Ref. | | | | Ref. | |
|  | Cluster 2 | 0.141 | 0.109 | **-0.239** | | **0.012** | 0.098 | 0.979 | -0.646 | | 0.362 | | 0.217 | 0.536 |
|  | Male |  |  | - | | |  |  | - | | | |  |  |
|  | Cluster 1 | Ref. | |  |  |  | Ref. | |  |  |  |  | Ref. | |
|  | Cluster 2 | -0.093 | 0.486 |  |  |  | 0.000 | 1.000^#^ |  |  |  |  | 0.521 | 0.278 |
| Natural spaces | Female |  |  |  | | |  |  |  | | | |  |  |
|  | Cluster 1 | Ref. | | Ref. | | | Ref. | | Ref. | | | | Ref. | |
|  | Cluster 2 | 0.115 | 0.143^#^ | -0.040 | 0.627^#^ | | -0.010 | 0.979^#^ | -0.452 | 0.368 | | | 0.100 | 0.618^#^ |
|  | Male |  |  | - | | |  |  | - | | | |  |  |
|  | Cluster 1 | Ref. | |  |  |  | Ref. | |  |  |  |  | Ref. | |
|  | Cluster 2 | -0.160 | 0.262 |  |  |  | 0.124 | 1.000 |  |  |  |  | 0.117 | 0.649^#^ |
| Air pollution & Traffic | Female |  |  |  | | |  |  |  | | | |  |  |
|  | Cluster 1 | Ref. | | Ref. | | | Ref. | | Ref. | | | | Ref. | |
|  | Cluster 2 | 0.219 | 0.095 | -0.182 | 0.135 | | -0.295 | 0.979 | -0.351 | 0.368^#^ | | | 0.280 | 0.466 |
|  | Male |  |  | - | | |  |  | - | | | |  |  |
|  | Cluster 1 | Ref. | |  |  |  | Ref. | |  |  |  |  | Ref. | |
|  | Cluster 2 | -0.012 | 0.919^#^ |  |  |  | 0.384 | 0.946 |  |  |  |  | 0.295 | 0.331 |

Cluster 1 represents a more favourable or healthier environmental profile, characterised by greater availability of green spaces, vegetation and land-use diversity, and lower building density, traffic intensity, and air pollution levels. Cluster 2 represents a less favourable or unhealthier environmental profile, characterised by higher building density, traffic intensity, and pollutant concentrations, and lower access to natural spaces. Cluster 1 was used as the reference category in the present analysis. P-values are FDR adjusted using the Benjamini-Hochberg procedure. FDR correction was applied separately by cohort, sex, outcome, exposure time point, and urban environmental subdomain. ^#^FDR-corrected p-values were identical to the nominal p-values.

**Supplementary Table 9.** Associations between clusters of environmental subdomains (natural spaces, built environment, and air pollution and traffic) and pubertal timing indicators, restricted to normal weight (G21 only).

|  |  | G21 | | | | |
| --- | --- | --- | --- | --- | --- | --- |
|  |  | **Tanner stage** | | **Menarche age** | | |
|  |  | **β** | **95% CI** | **β** | | **95% CI** |
| Birth |  |  |  |  | |  |
| Built environment | Female |  |  |  | |  |
|  | Cluster 1 | Ref. | | Ref. | | |
|  | Cluster 2 | 0.118 | 0.280^#^ | -0.200 | | 0.260 |
|  | Male |  |  | **-** | | |
|  | Cluster 1 | Ref. | |  |  |  |
|  | Cluster 2 | -0.054 | 0.887 |  |  |  |
| Natural spaces | Female |  |  |  | | |
|  | Cluster 1 | Ref. | | Ref. | | |
|  | Cluster 2 | 0.208 | 0.243 | -0.095 | 0.455^#^ | |
|  | Male |  |  | - | | |
|  | Cluster 1 | Ref. | |  |  |  |
|  | Cluster 2 | 0.022 | 0.887^#^ |  |  |  |
| Air pollution & Traffic | Female |  |  |  | | |
|  | Cluster 1 | Ref. | | Ref. | | |
|  | Cluster 2 | 0.166 | 0.280 | -0.195 | 0.344 | |
|  | Male |  |  | - | | |
|  | Cluster 1 | Ref. | |  |  |  |
|  | Cluster 2 | 0.079 | 0.887 |  |  |  |
| 1-1.5 years |  |  | |  | | |
| Built environment | Female |  | |  | | |
|  | Cluster 1 | Ref. | | Ref. | | |
|  | Cluster 2 | 0.089 | 0.387^#^ | **-0.266** | | **0.047** |
|  | Male |  | | - | | |
|  | Cluster 1 | Ref. | |  |  |  |
|  | Cluster 2 | -0.039 | 0.906 |  |  |  |
| Natural spaces | Female |  |  |  | | |
|  | Cluster 1 | Ref. | | Ref. | | |
|  | Cluster 2 | 0.147 | 0.361 | -0.151 | 0.295 | |
|  | Male |  |  | - | | |
|  | Cluster 1 | Ref. | |  |  |  |
|  | Cluster 2 | -0.019 | 0.906^#^ |  |  |  |
| Air pollution & Traffic | Female |  |  |  | | |
|  | Cluster 1 | Ref. | | Ref. | | |
|  | Cluster 2 | 0.188 | 0.361 | -0.165 | 0.295 | |
|  | Male |  |  | - | | |
|  | Cluster 1 | Ref. | |  |  |  |
|  | Cluster 2 | 0.084 | 0.906 |  |  |  |
| 4-5 years |  |  |  |  | |  |
| Built environment | Female |  |  |  | |  |
|  | Cluster 1 | Ref. | | Ref. | | |
|  | Cluster 2 | 0.199 | 0.053^#^ | -0.209 | | 0.170 |
|  | Male |  |  | - | | |
|  | Cluster 1 | Ref. | |  |  |  |
|  | Cluster 2 | -0.013 | 0.920 |  |  |  |
| Natural spaces | Female |  |  |  | | |
|  | Cluster 1 | Ref. | | Ref. | | |
|  | Cluster 2 | **0.222** | **0.047** | -0.137 | 0.320 | |
|  | Male |  |  | - | | |
|  | Cluster 1 | Ref. | |  |  |  |
|  | Cluster 2 | 0.041 | 0.920 |  |  |  |
| Air pollution & Traffic | Female |  |  |  | | |
|  | Cluster 1 | Ref. | | Ref. | | |
|  | Cluster 2 | **0.278** | **0.047** | -0.081 | 0.537^#^ | |
|  | Male |  |  | - | | |
|  | Cluster 1 | Ref. | |  |  |  |
|  | Cluster 2 | 0.160 | 0.917 |  |  |  |

Cluster 1 represents a more favourable or healthier environmental profile, characterised by greater availability of green spaces, vegetation and land-use diversity, and lower building density, traffic intensity, and air pollution levels. Cluster 2 represents a less favourable or unhealthier environmental profile, characterised by higher building density, traffic intensity, and pollutant concentrations, and lower access to natural spaces. Cluster 1 was used as the reference category in the present analysis. P-values are FDR adjusted using the Benjamini-Hochberg procedure. FDR correction was applied separately by cohort, sex, outcome, exposure time point, and urban environmental subdomain. ^#^FDR-corrected p-values were identical to the nominal p-values.

**Section III. Supplementary figures**

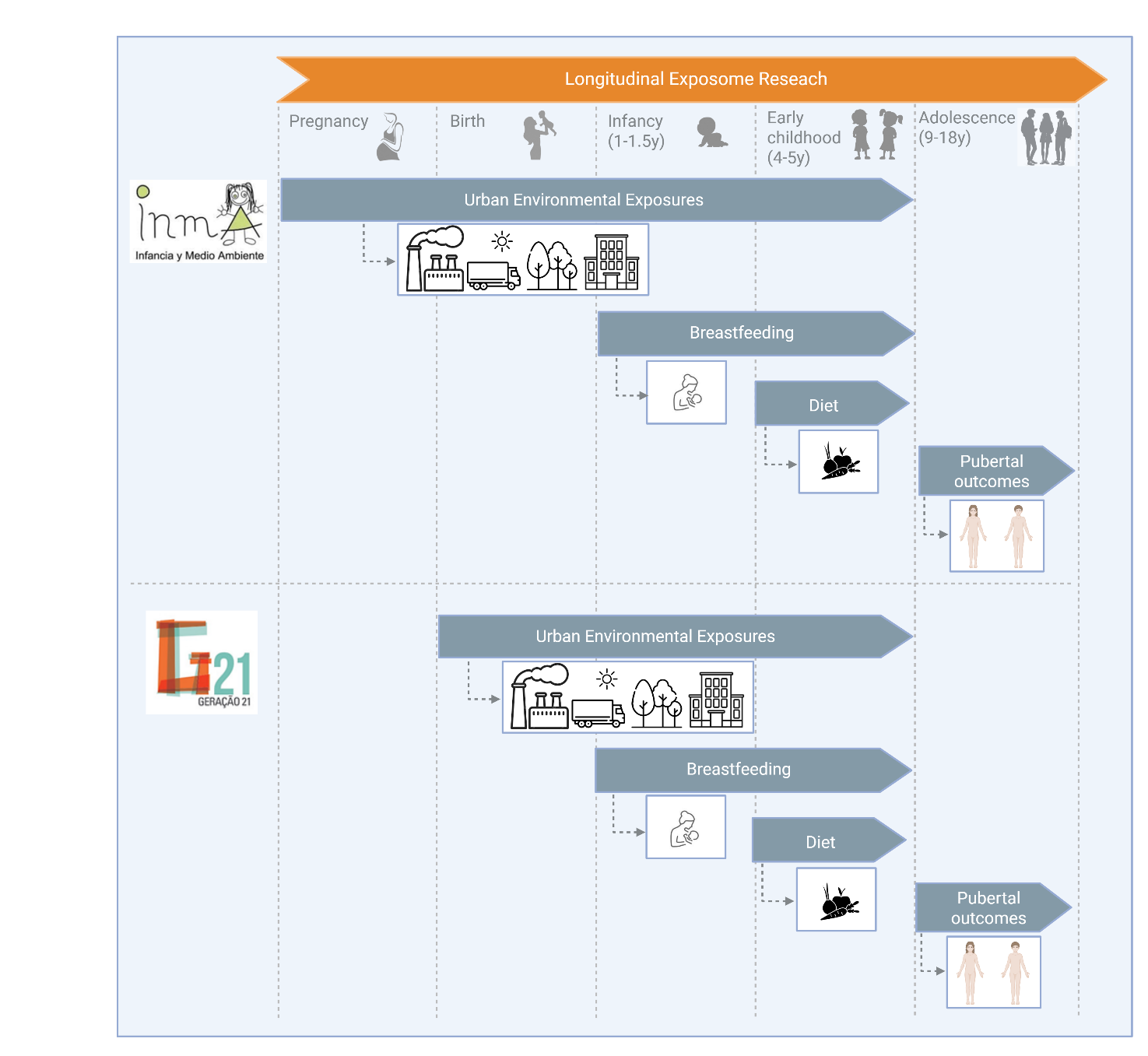

**Supplementary Figure 1**. **Timeline of exposures and pubertal outcomes assessed in the INMA and G21 cohorts.** The figure illustrates the timing of assessment of urban environmental exposures (built environment, natural spaces, and air pollution & traffic), exclusive breastfeeding duration, diet quality, and pubertal outcomes across different time points in the INMA and G21 birth cohorts.

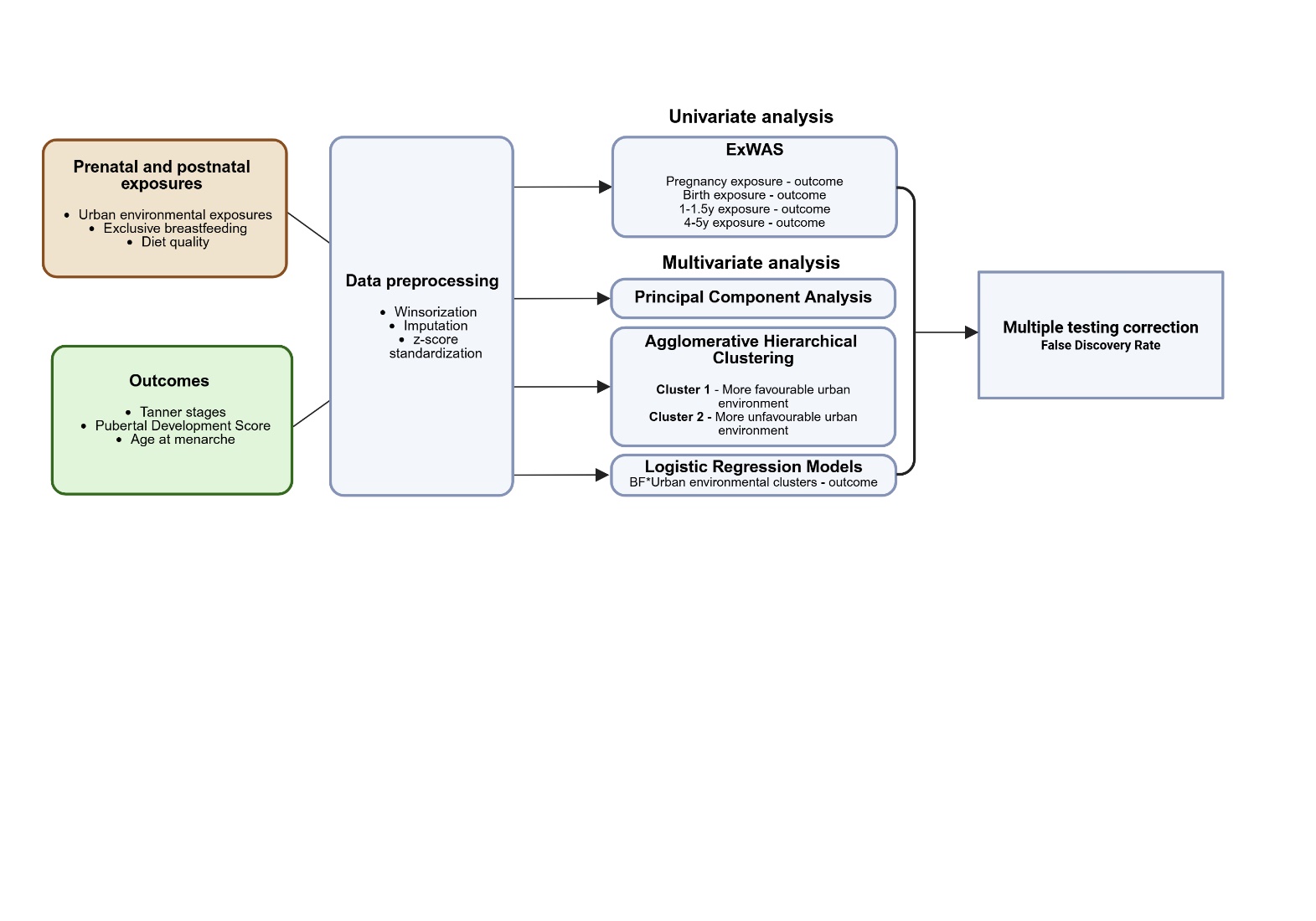

**Supplementary Figure 2.** **Statistical analysis pipeline.** Abbreviations: IQR, Interquartile range; ExWAS, Exposome-wide association study; BF, breastfeeding.

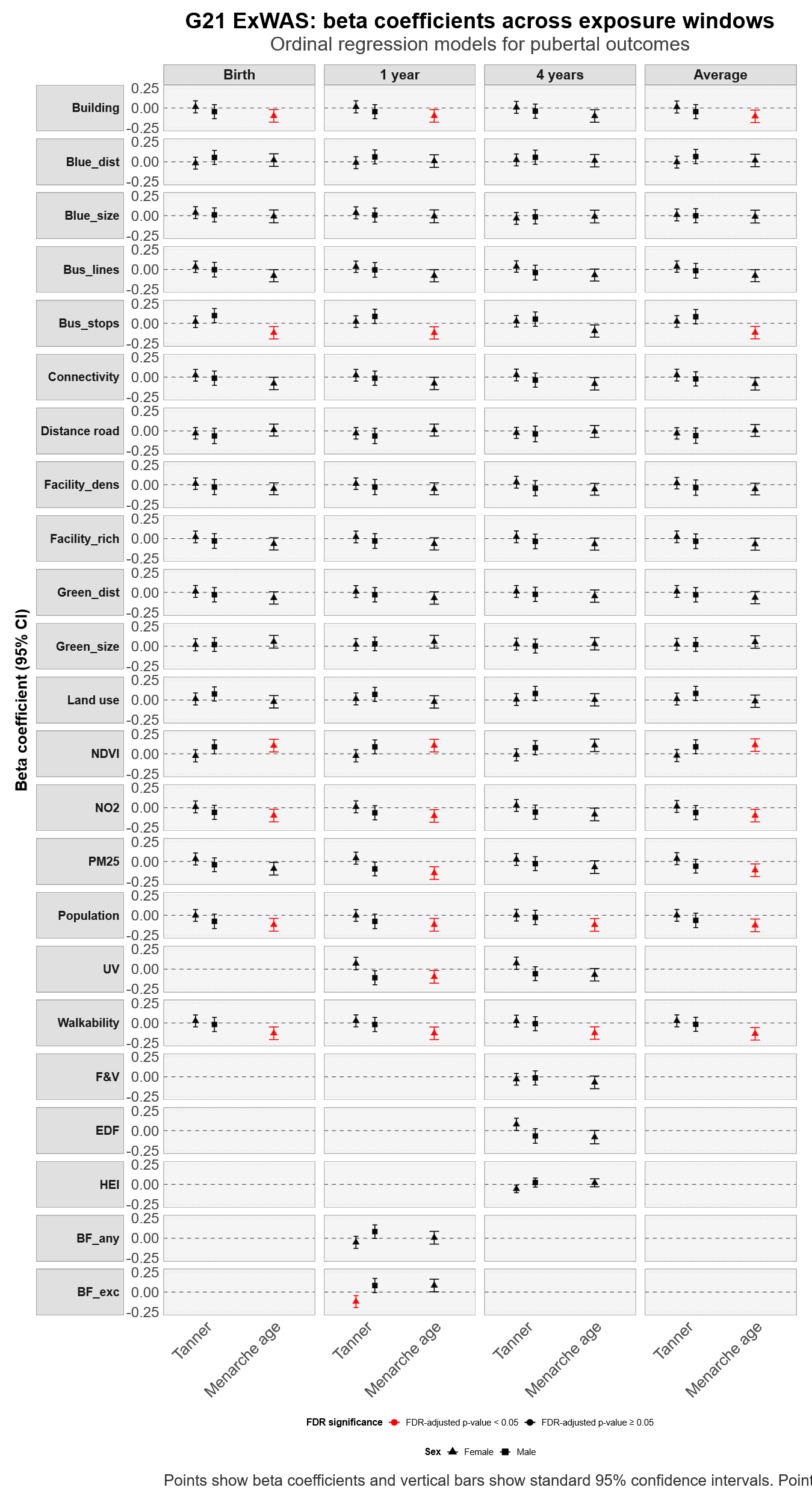
a)

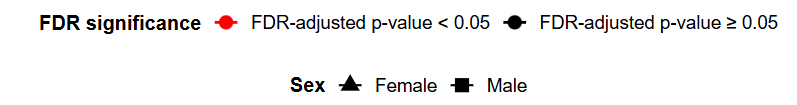

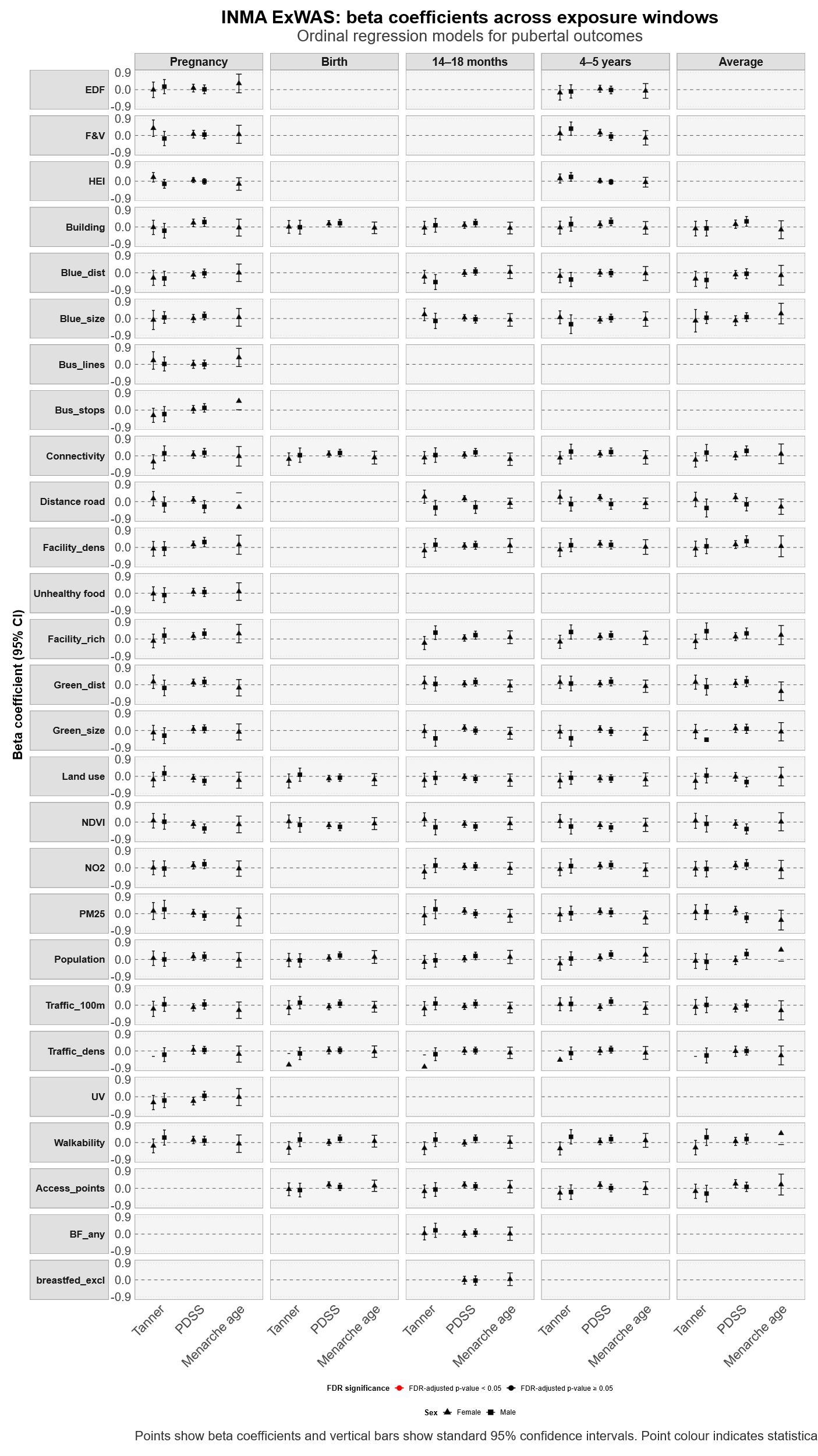
b)

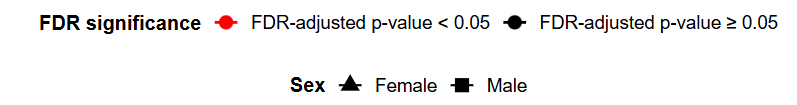

**Supplementary Figure 3** - **Standardized beta estimates from ExWAS analyses of urban environment, breastfeeding duration, and diet in relation to pubertal development at 10 years and age at menarche in the G21 and INMA cohorts.** a) G21 cohort; b) INMA cohort. Each ExWAS was conducted separately for each outcome and exposure time point, with false discovery rate (FDR) correction applied based on the number of exposures tested in each ExWAS. Results with FDR-adjusted p-values <0.05 are highlighted in red. Points show beta coefficients and vertical bars show standard 95% confidence intervals. Models are stratified by sex. Abbreviations: Access_points, number of bus public transport mode stops; Building, building density; Blue_dist, distance to nearest major blue space; Blue_size, size of the nearest major blue space; Green_dist, distance to nearest major green space; Green_size, size of the nearest major green space; Bus_lines, length of public transport lines; Bus_stops, density of public bus stops; Connectivity, connectivity density; Distance road, inverse distance to nearest road; Facility_dens, facility density; Facility_rich, facility richness; Unhealthy food, unhealthy food facility density; Land use, land use Shannon’s Evenness Index; NDVI, Normalized Difference Vegetation Index ; NO2, nitrogen dioxide; PM25, particulate matter with an aerodynamic diameter of less than 2.5 μm; Population, population density; Traffic_100, traffic load on all roads in 100m buffer; Traffic_dens, traffic density in nearest road; UV, average of vitamin D UV dose; Walkability, walkability index; F&V, fruit and vegetables; EDF, energy-dense foods; HEI, healthy eating index; BF_any, non-exclusive breastfeeding duration; BF_exc, exclusive breastfeeding duration.

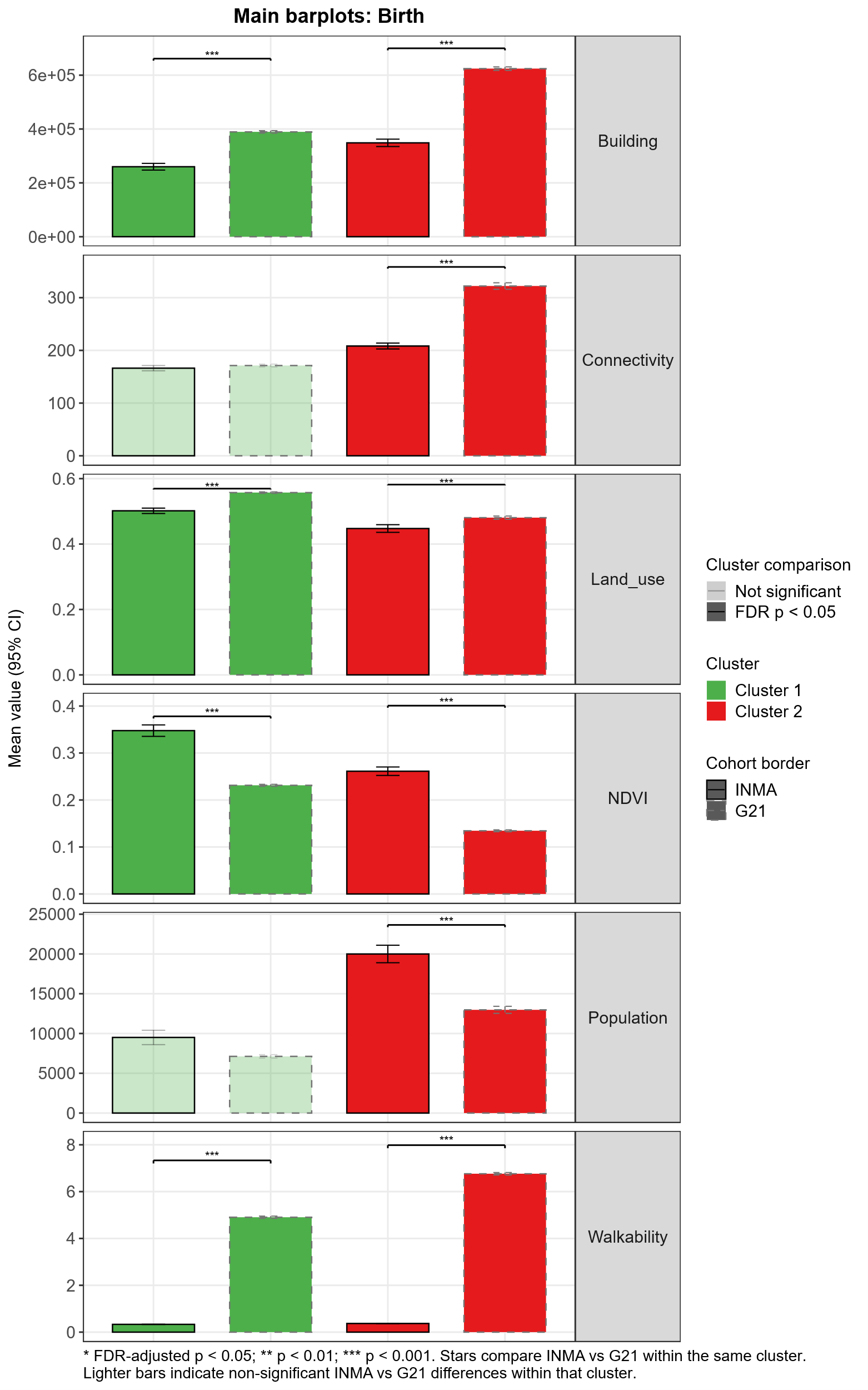

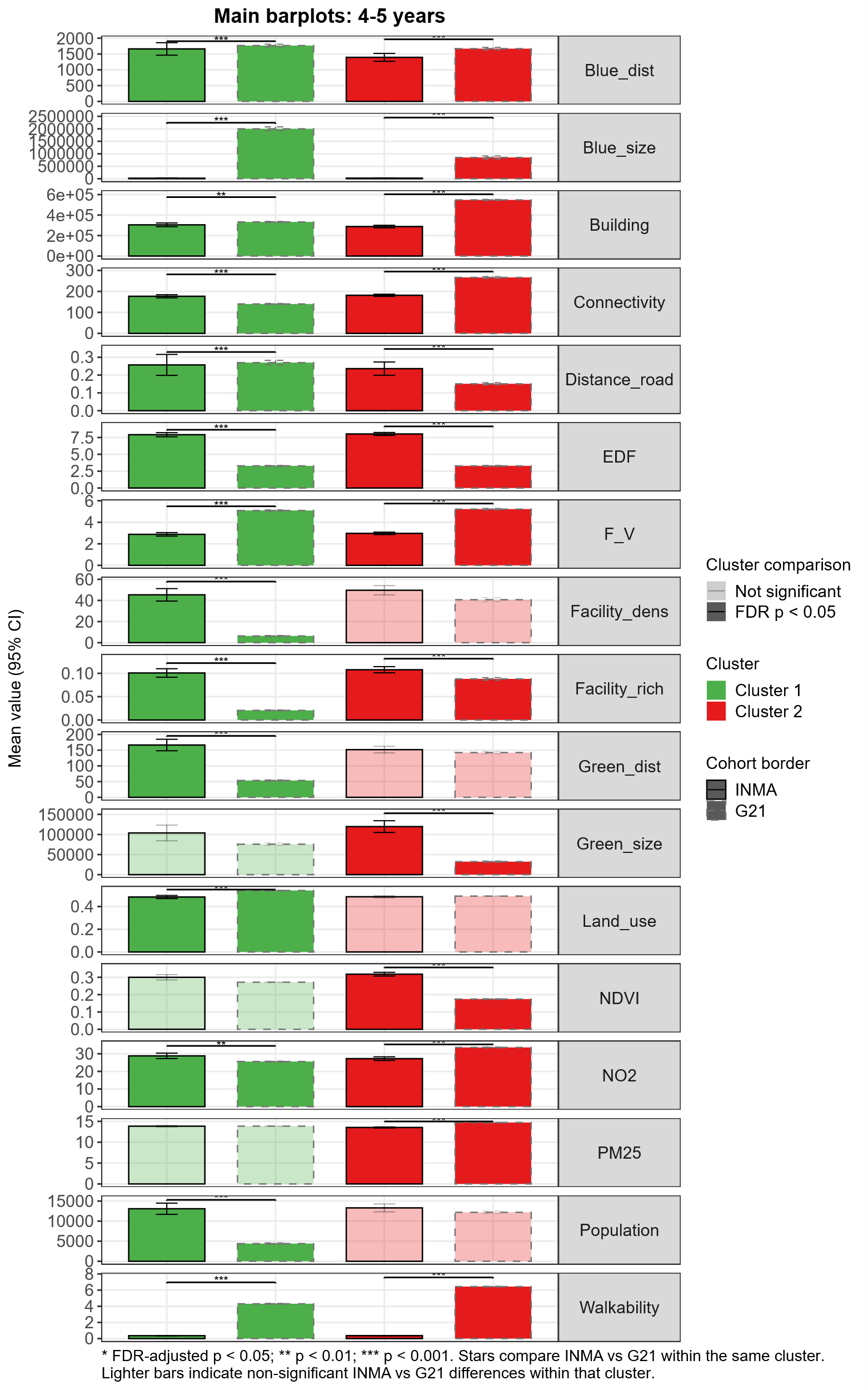

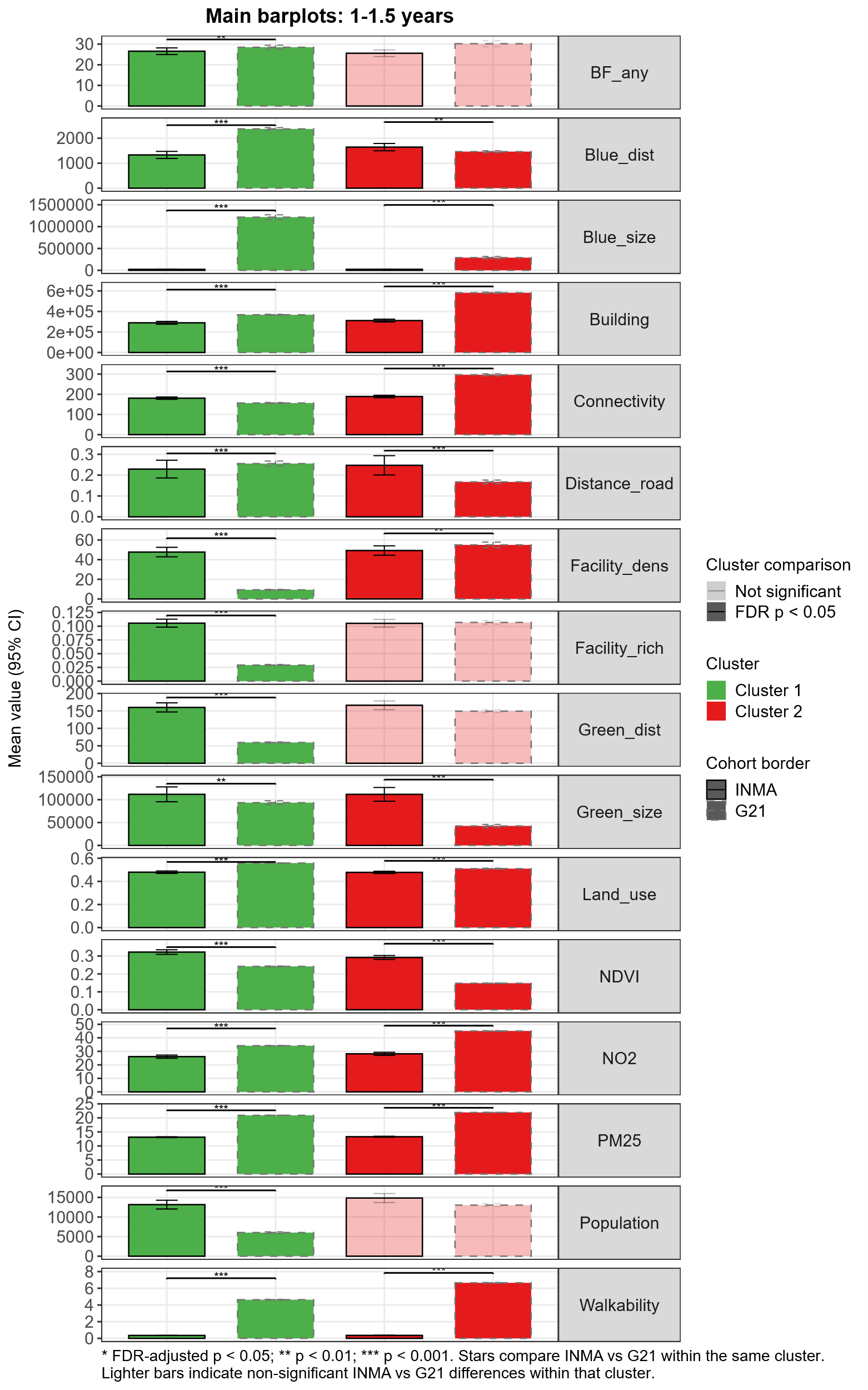
**.**

**Supplementary Figure 4. Urban environmental cluster characteristics across exposure time points in the G21 and INMA cohorts.** Cluster 1 represents a more favourable or healthier environmental profile, characterised by greater availability of green spaces, vegetation and land-use diversity, and lower building density, traffic intensity, and air pollution levels. Cluster 2 represents a less favourable or unhealthier environmental profile, characterised by higher building density, traffic intensity, and pollutant concentrations, and lower access to natural spaces. Stars compare INMA vs G21 cohorts within the same cluster based on FDR-adjusted p-values (* FDR-adjusted p < 0.05; ** FDR-adjusted p < 0.01; *** FDR-adjusted p < 0.001). Lighter bars indicate non-significant INMA vs G21 differences within that cluster. Abbreviations: Building, building density; Blue_dist, distance to nearest major blue space; Blue_size, size of the nearest major blue space; Green_dist, distance to nearest major green space; Green_size, size of the nearest major green space; Connectivity, connectivity density; Distance road, inverse distance to nearest road; Facility_dens, facility density; Facility_rich, facility richness; Land use, land use Shannon’s Evenness Index; NDVI, Normalized Difference Vegetation Index ; NO2, nitrogen dioxide; PM25, particulate matter with an aerodynamic diameter of less than 2.5 μm; Population, population density; Walkability, walkability index; F_V, fruit and vegetables; EDF, energy-dense foods; HEI, healthy eating index; BF_any, non-exclusive breastfeeding duration; BF_exc, exclusive breastfeeding duration.

Across all exposure windows in both cohorts, Cluster 1 was characterised by greater availability of green and blue spaces, higher vegetation levels, and lower building density, connectivity, population density, and air pollution levels. In contrast, Cluster 2 was characterised by higher building density, connectivity, population density, walkability, and air pollution levels, together with lower availability of natural spaces. Similar clustering patterns were observed across the G21 and INMA cohorts, although some differences were identified between cohorts. Compared with INMA, the G21 cohort presented higher values for building density, connectivity, walkability, and air pollution indicators, but also greater availability of blue and green spaces.

**Section IV. Results with no corrected p-values**

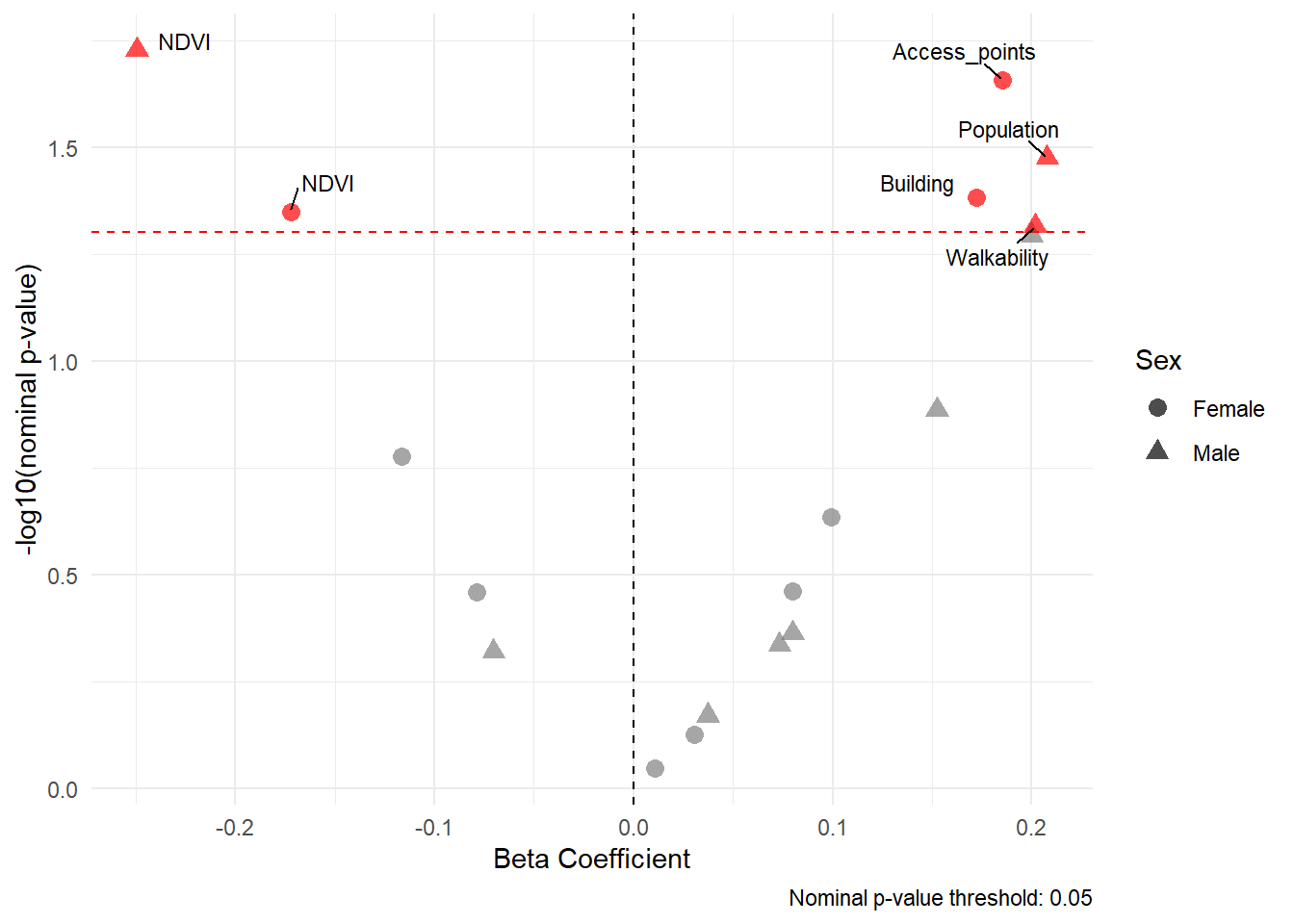

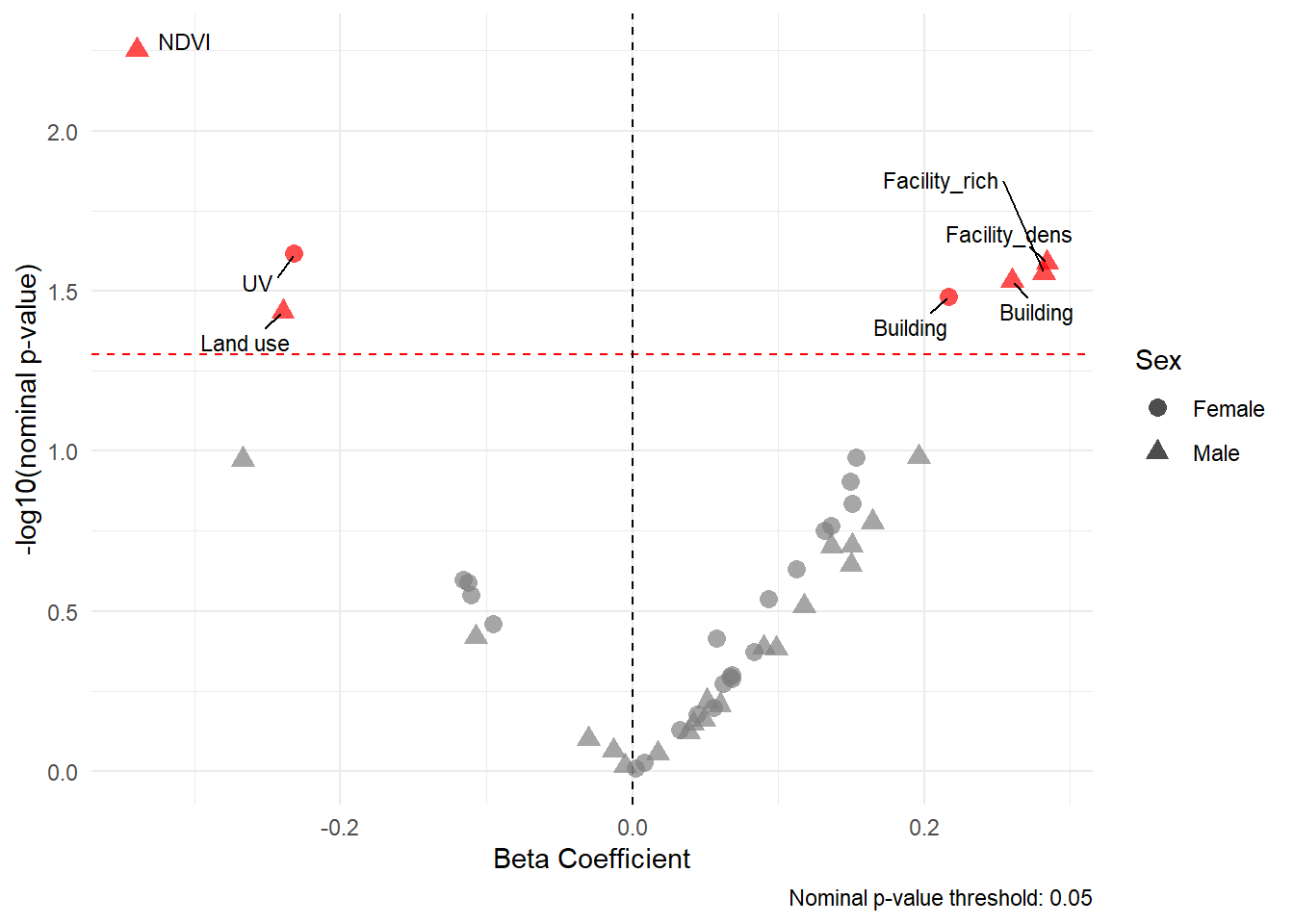
INMA – PDS score

a)	b)

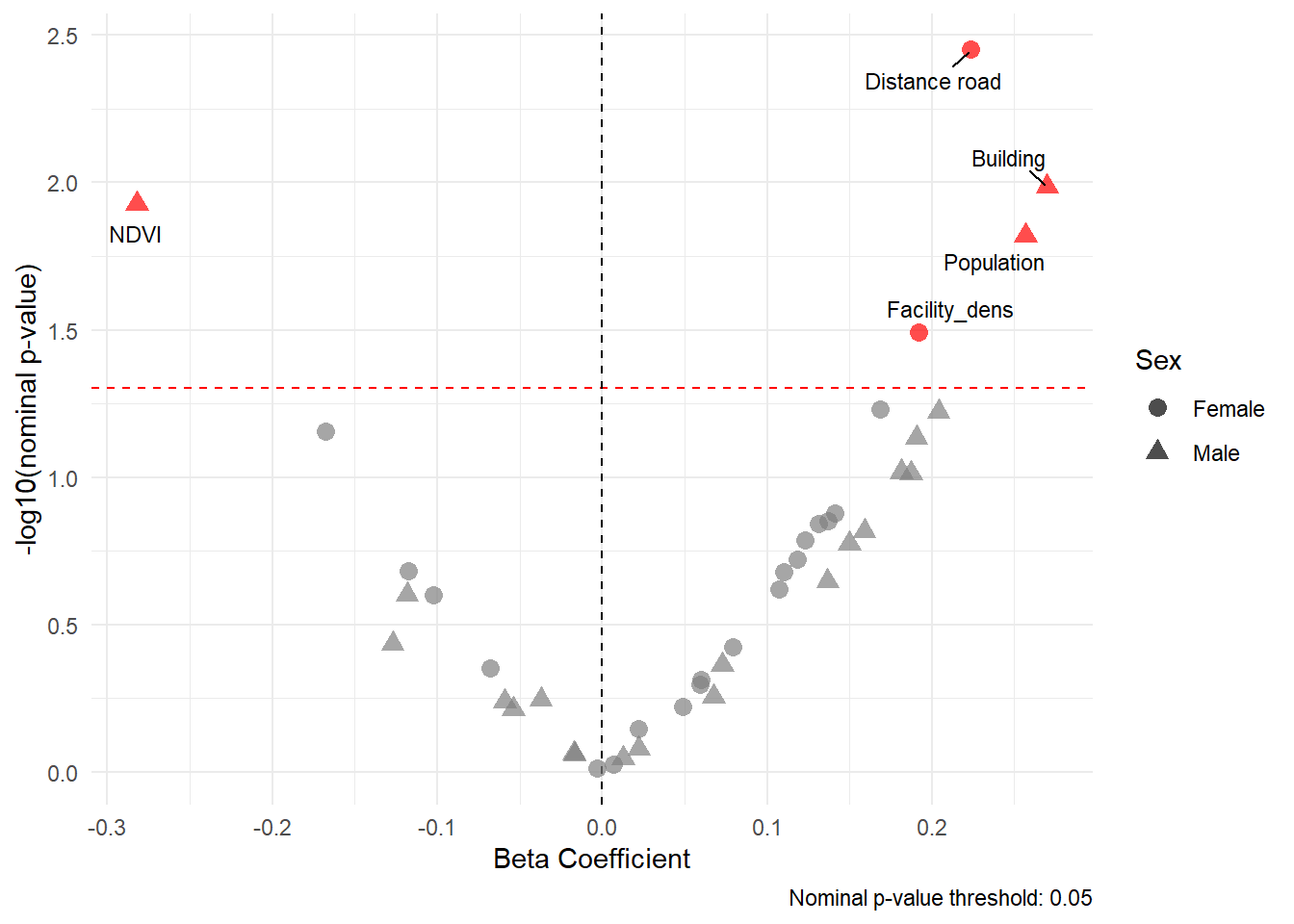

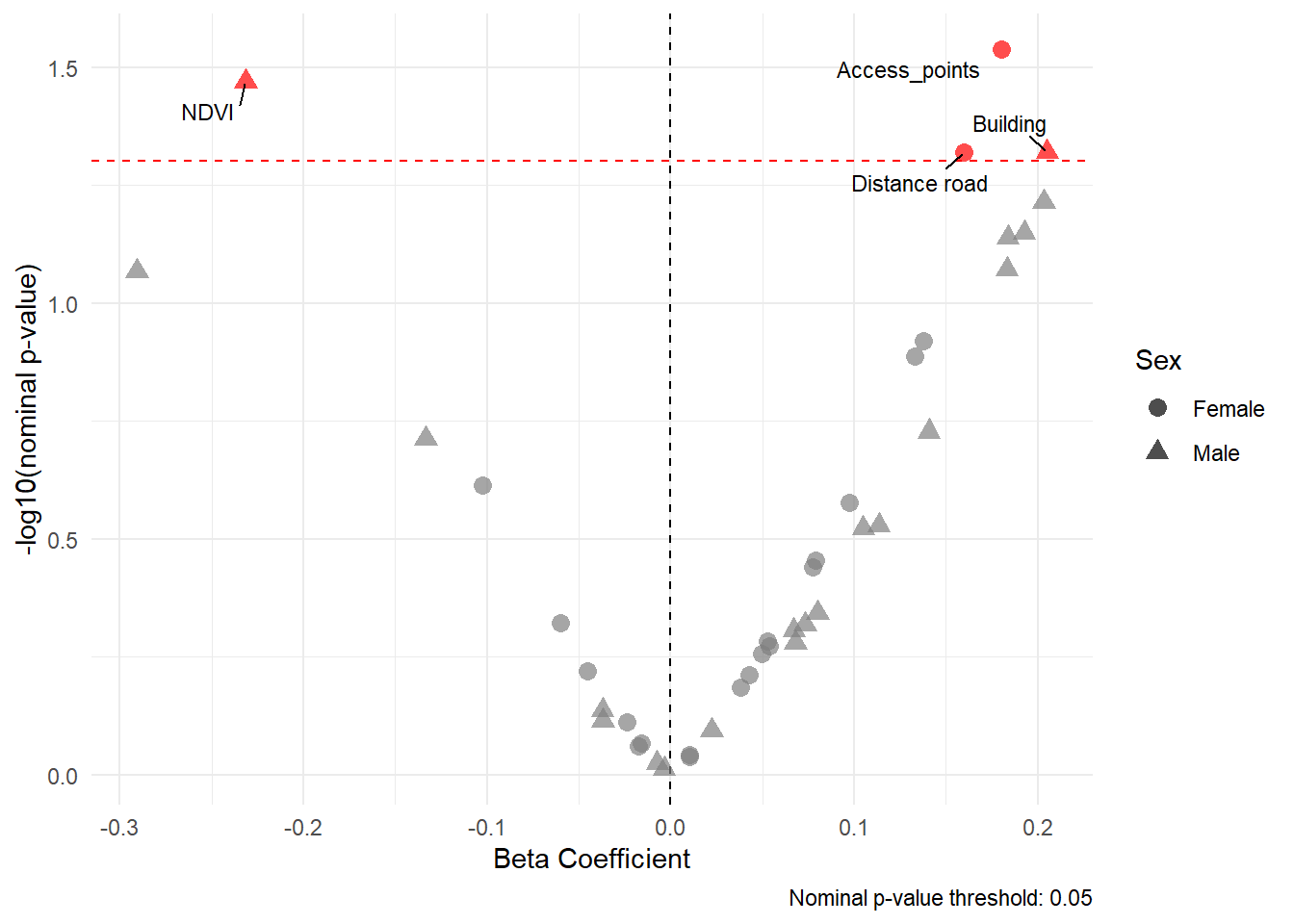

c)	d)

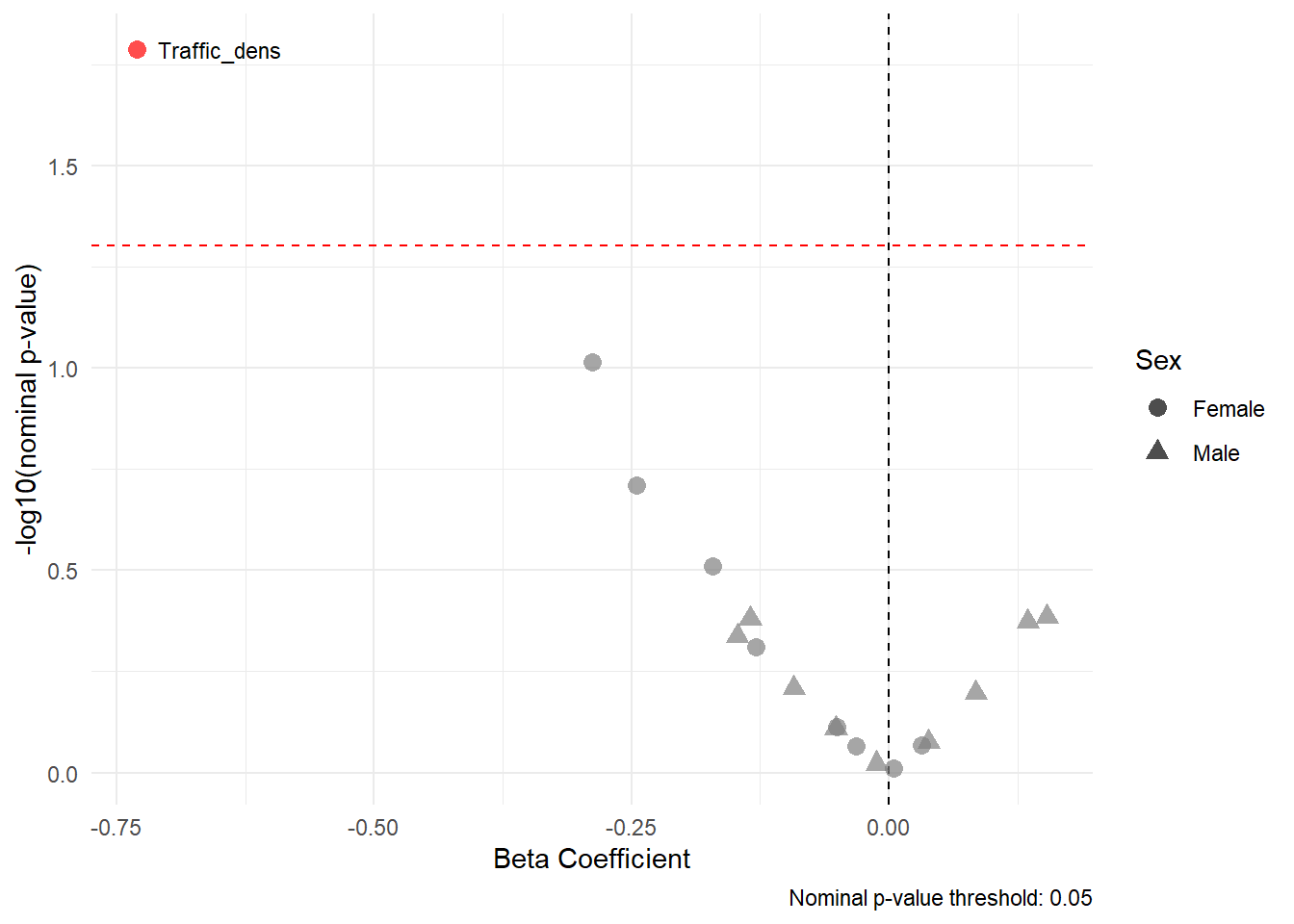

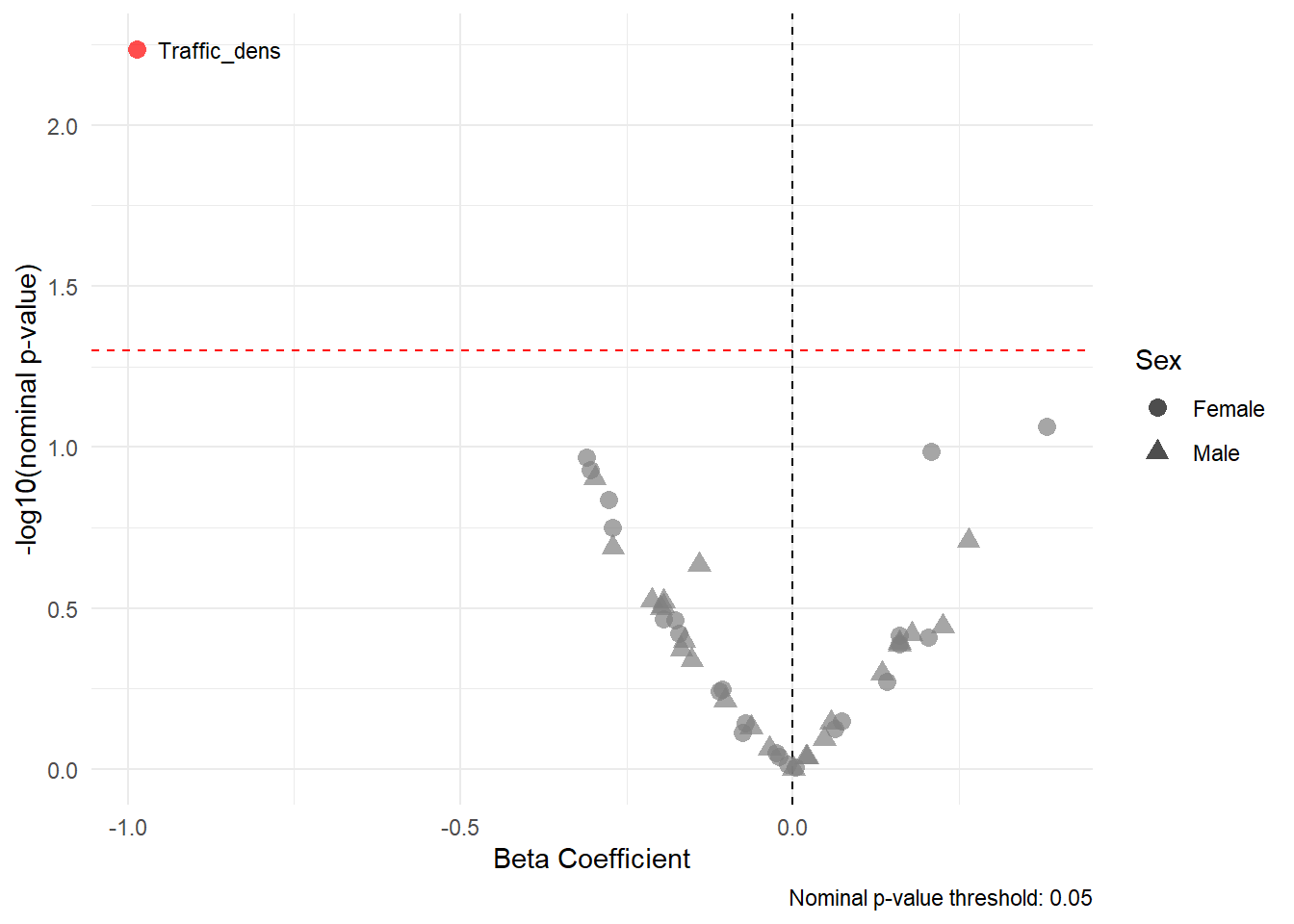
INMA - Tanner stages

e)	f)

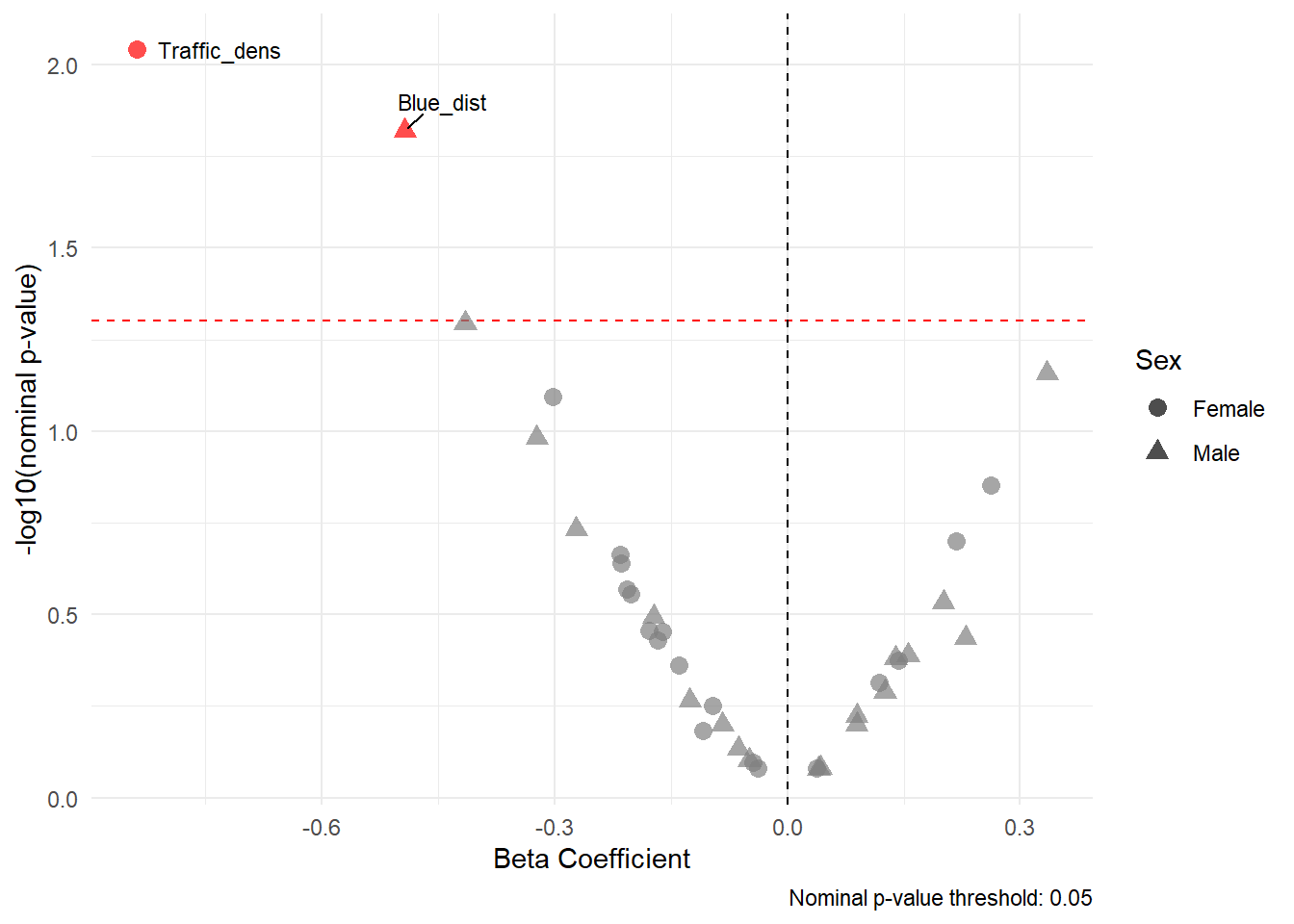

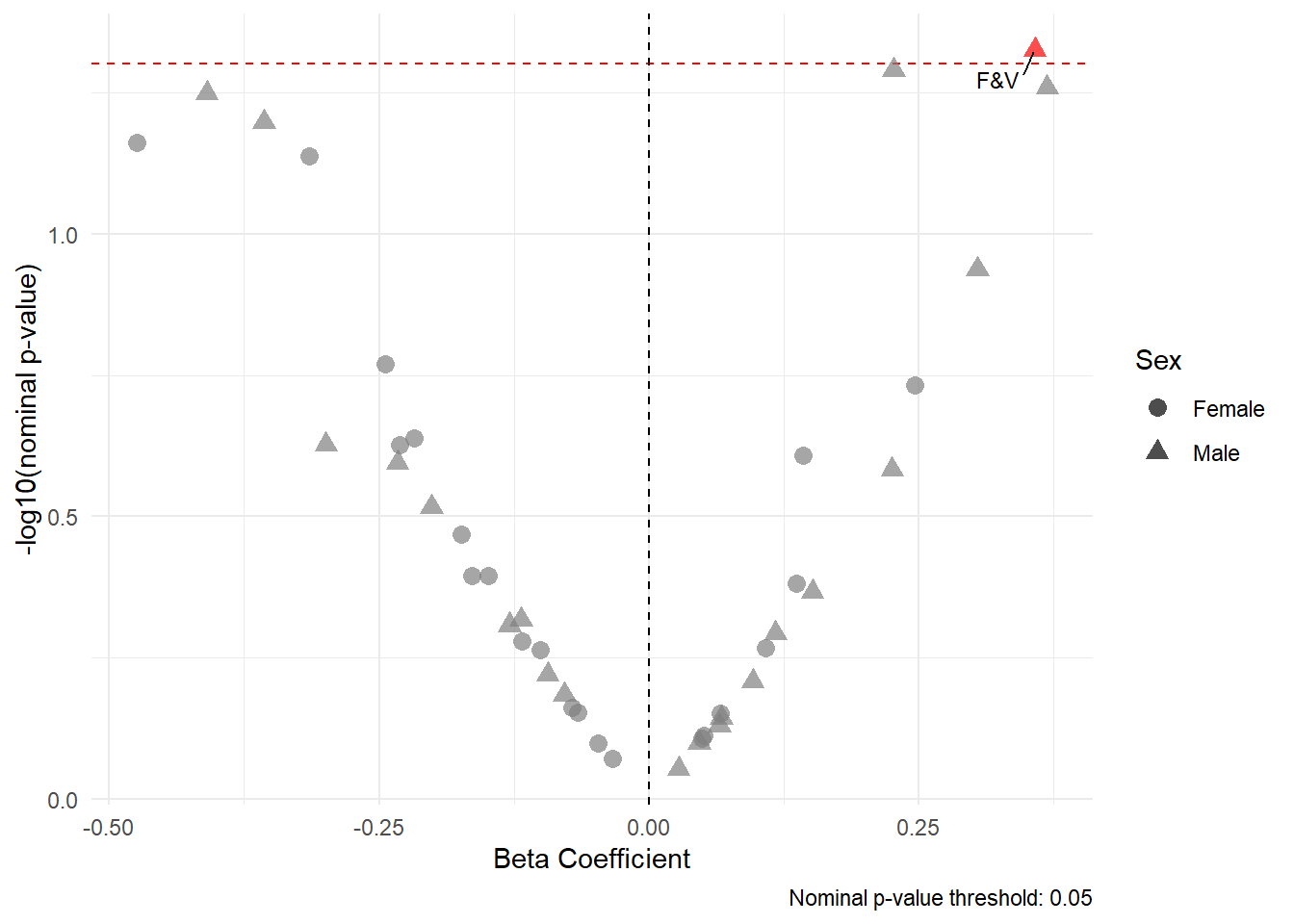
g)	h)

INMA – Age at menarche

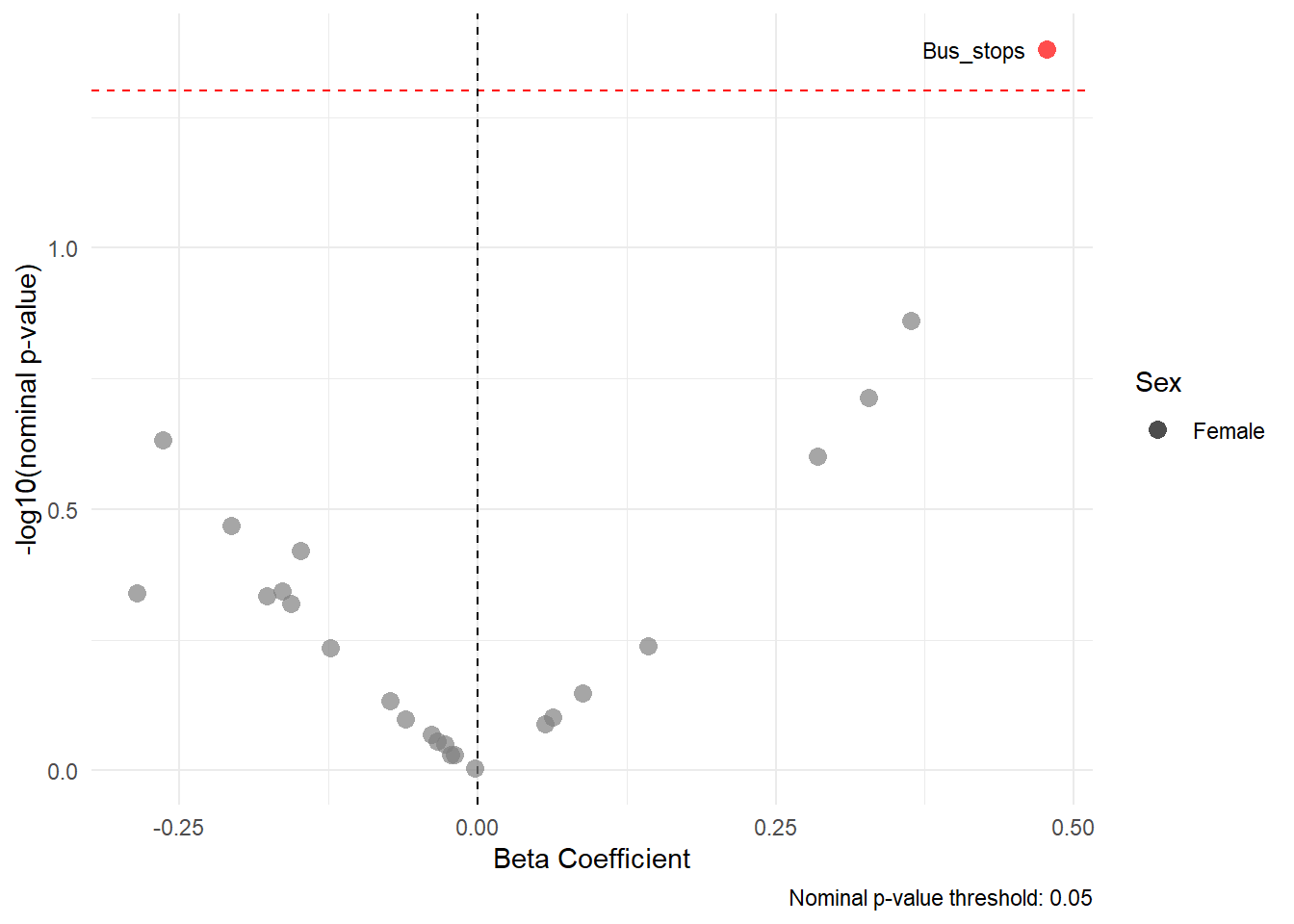

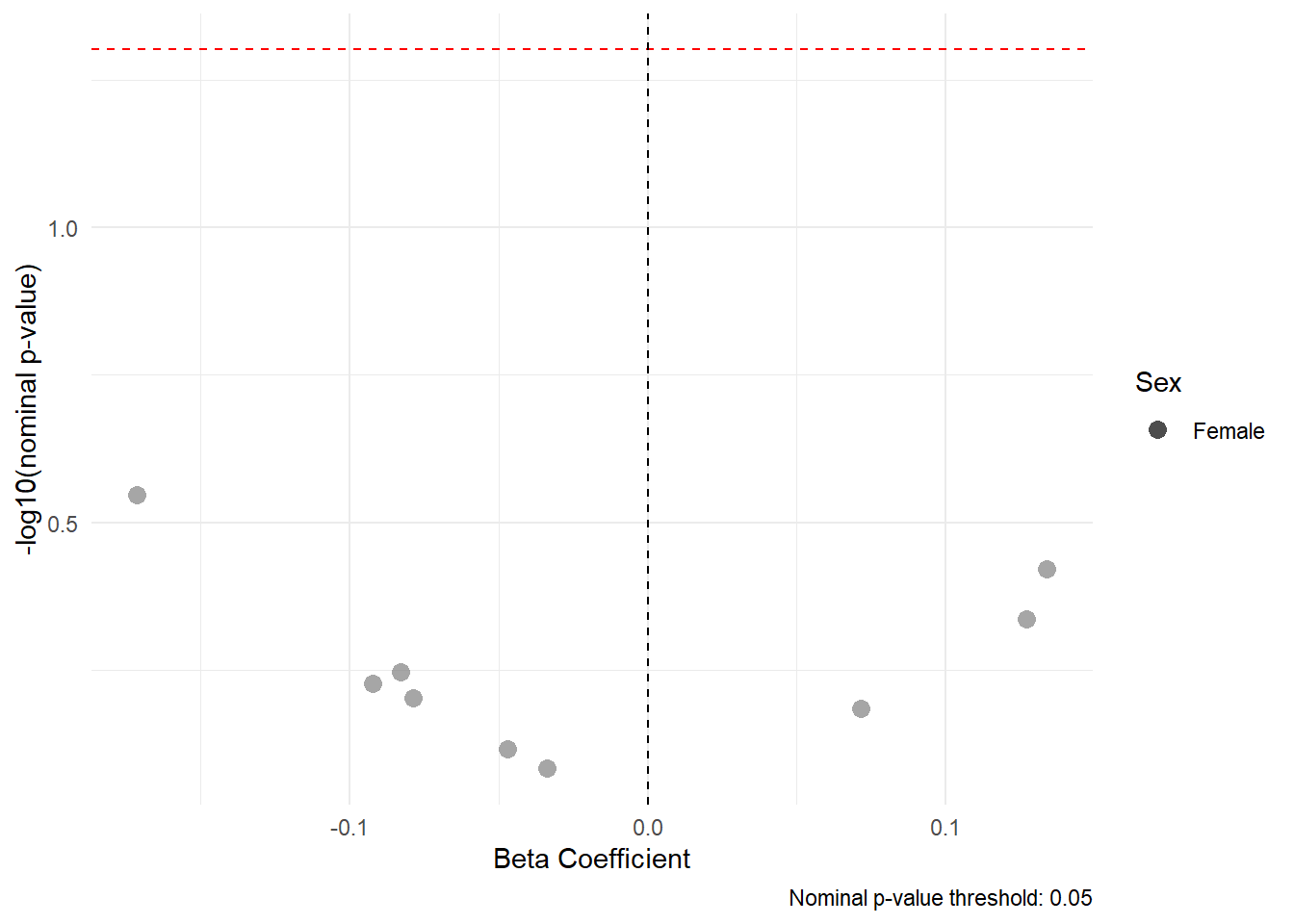
i)	j)

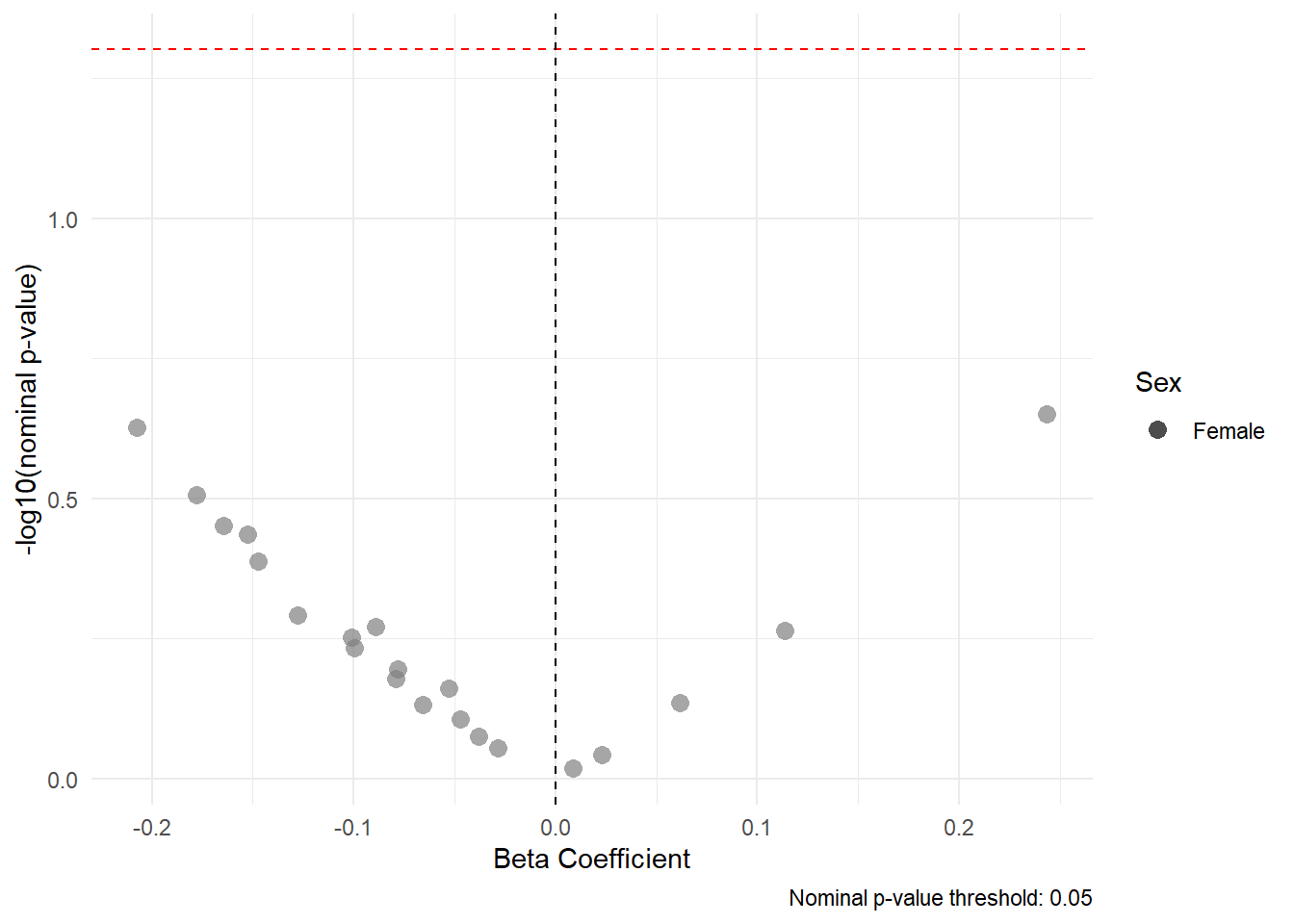

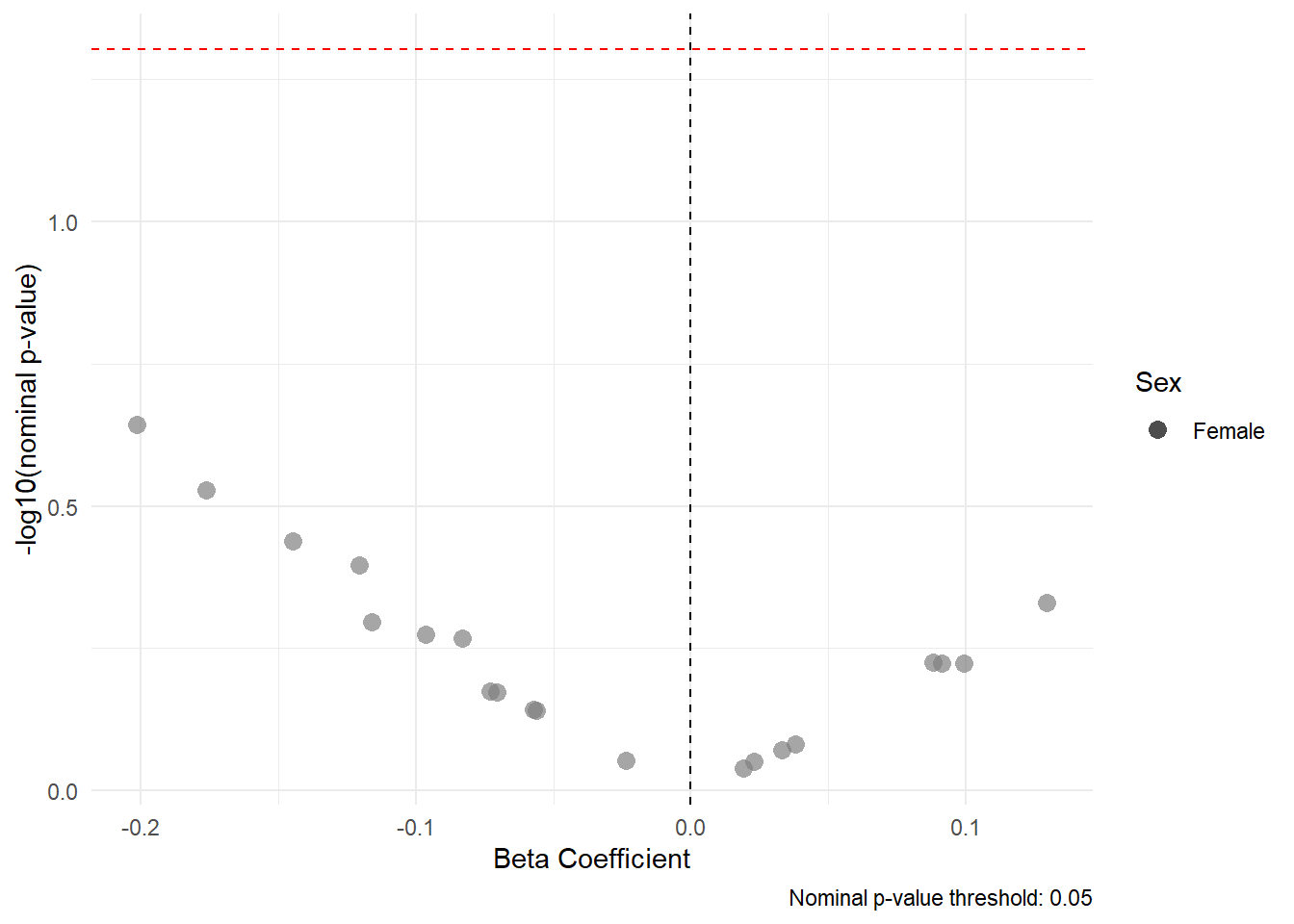

k)	l)

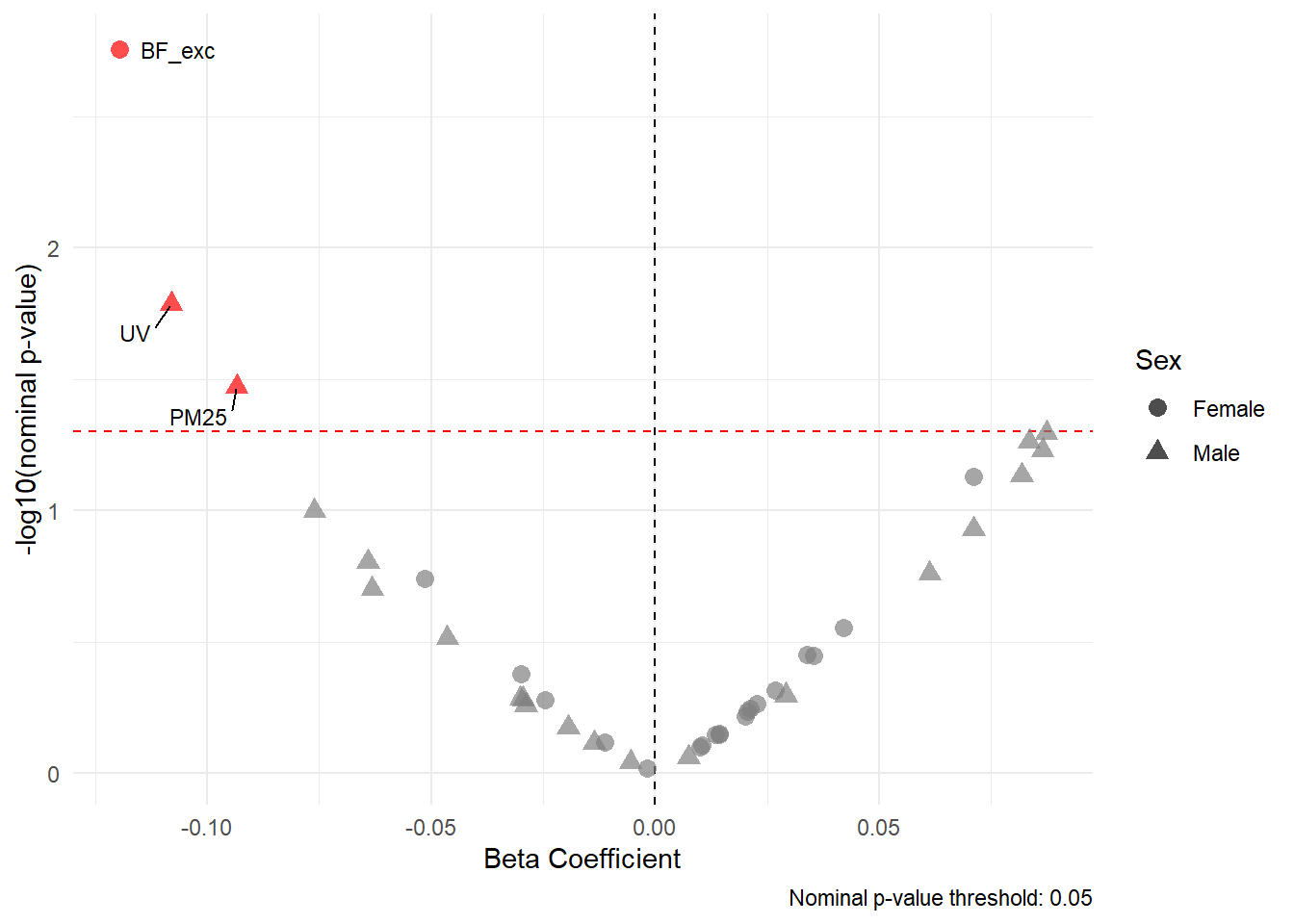
G21 – Tanner stages

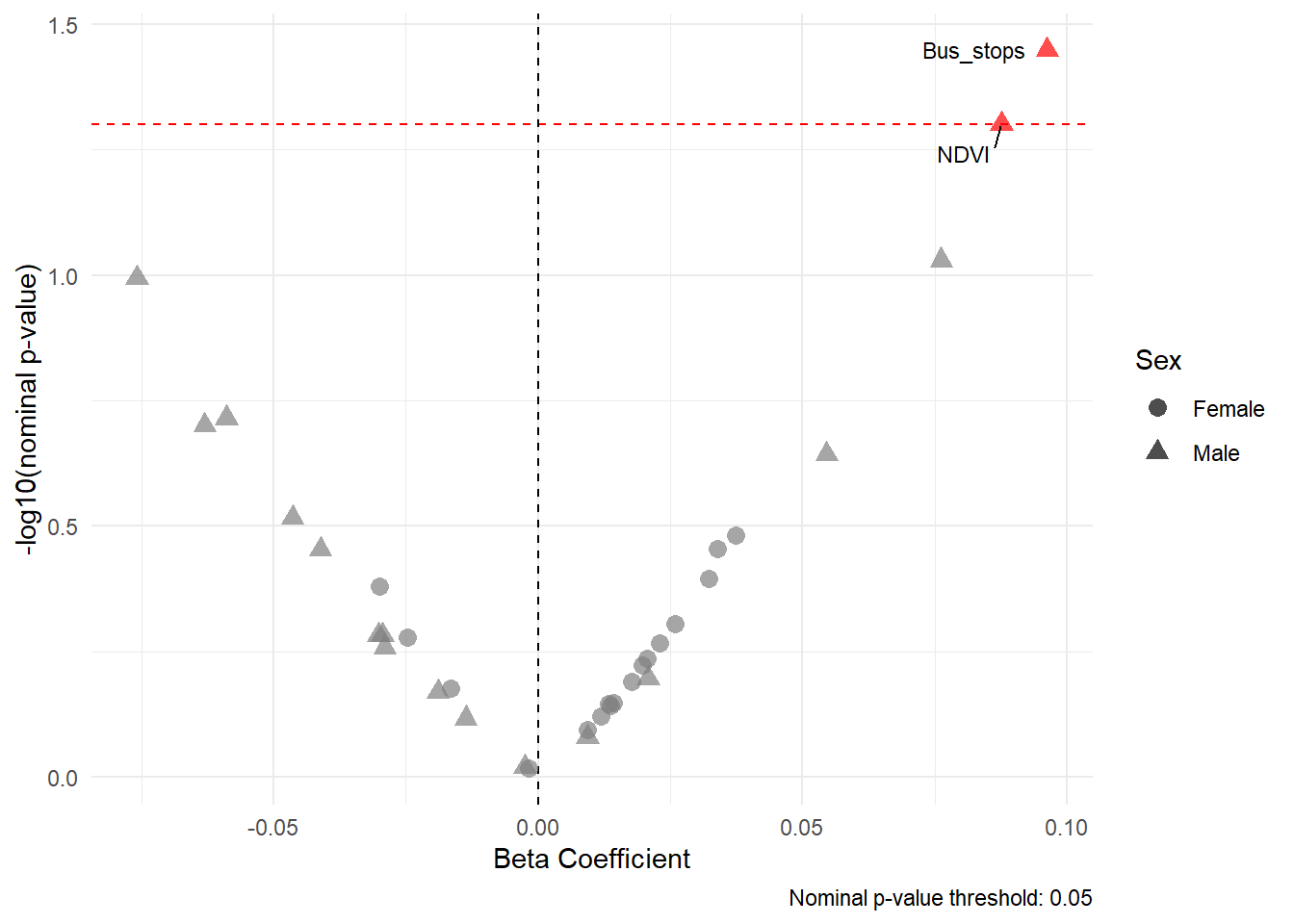
m)	n)

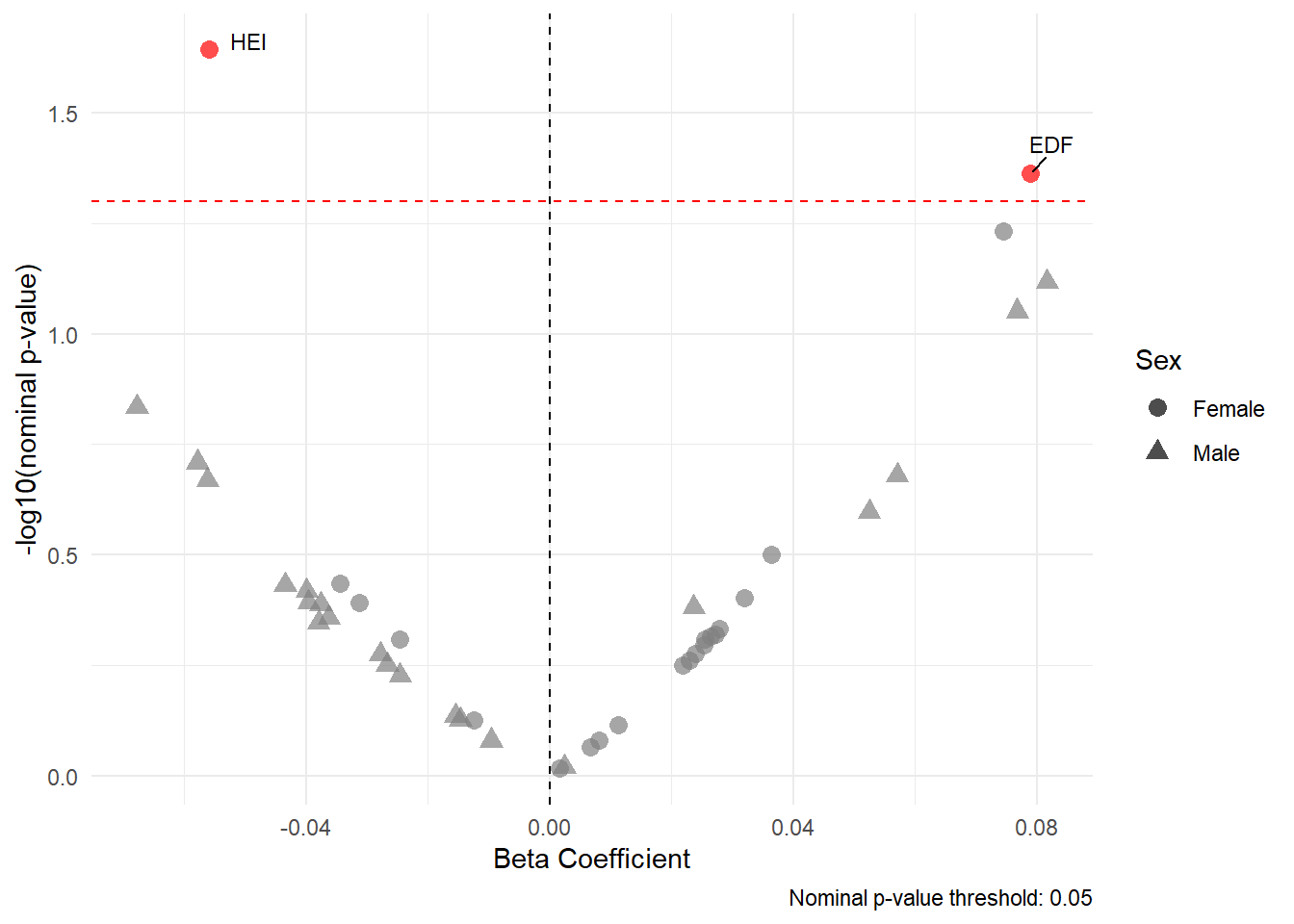

o)

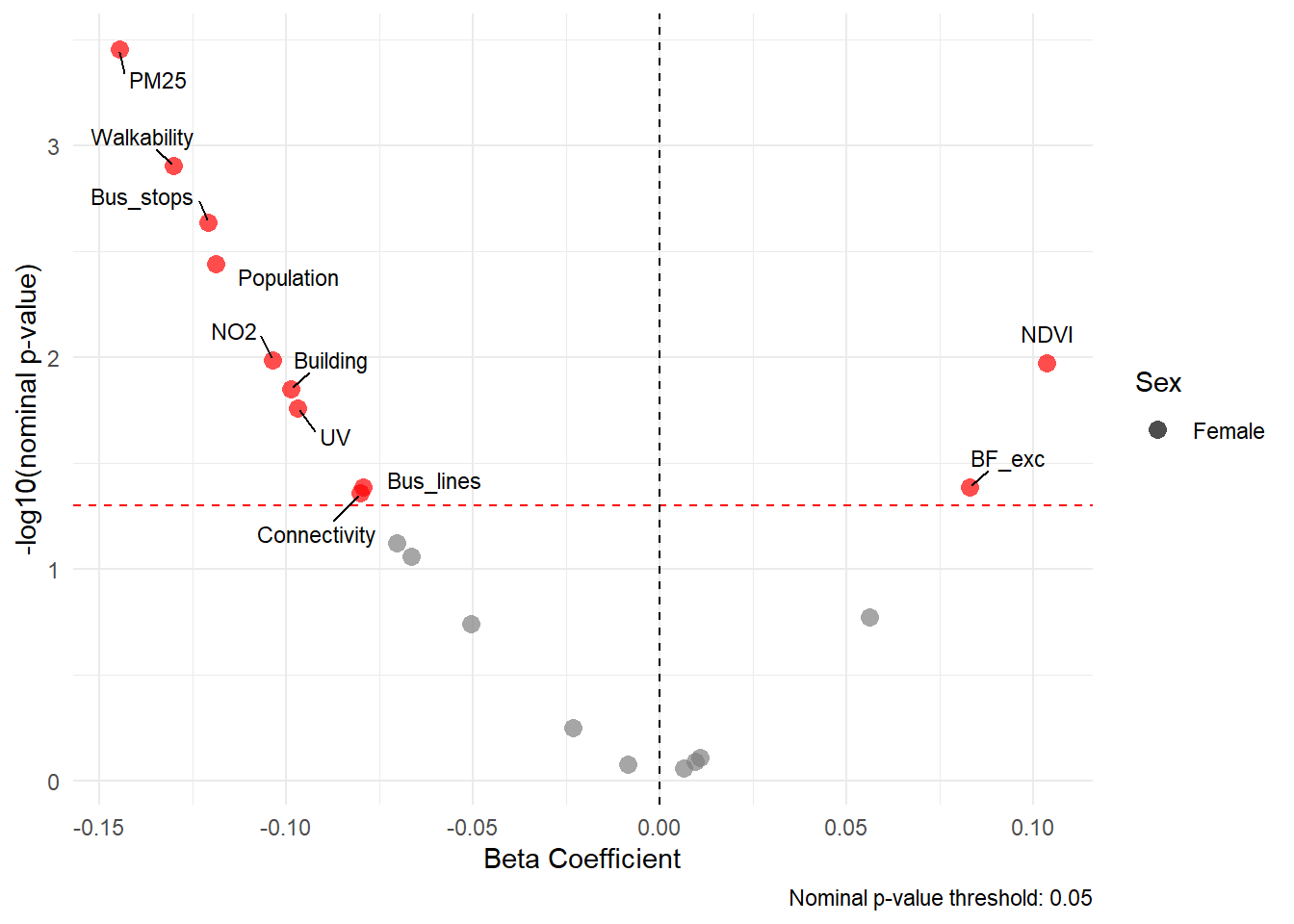
G21 – Age at menarche

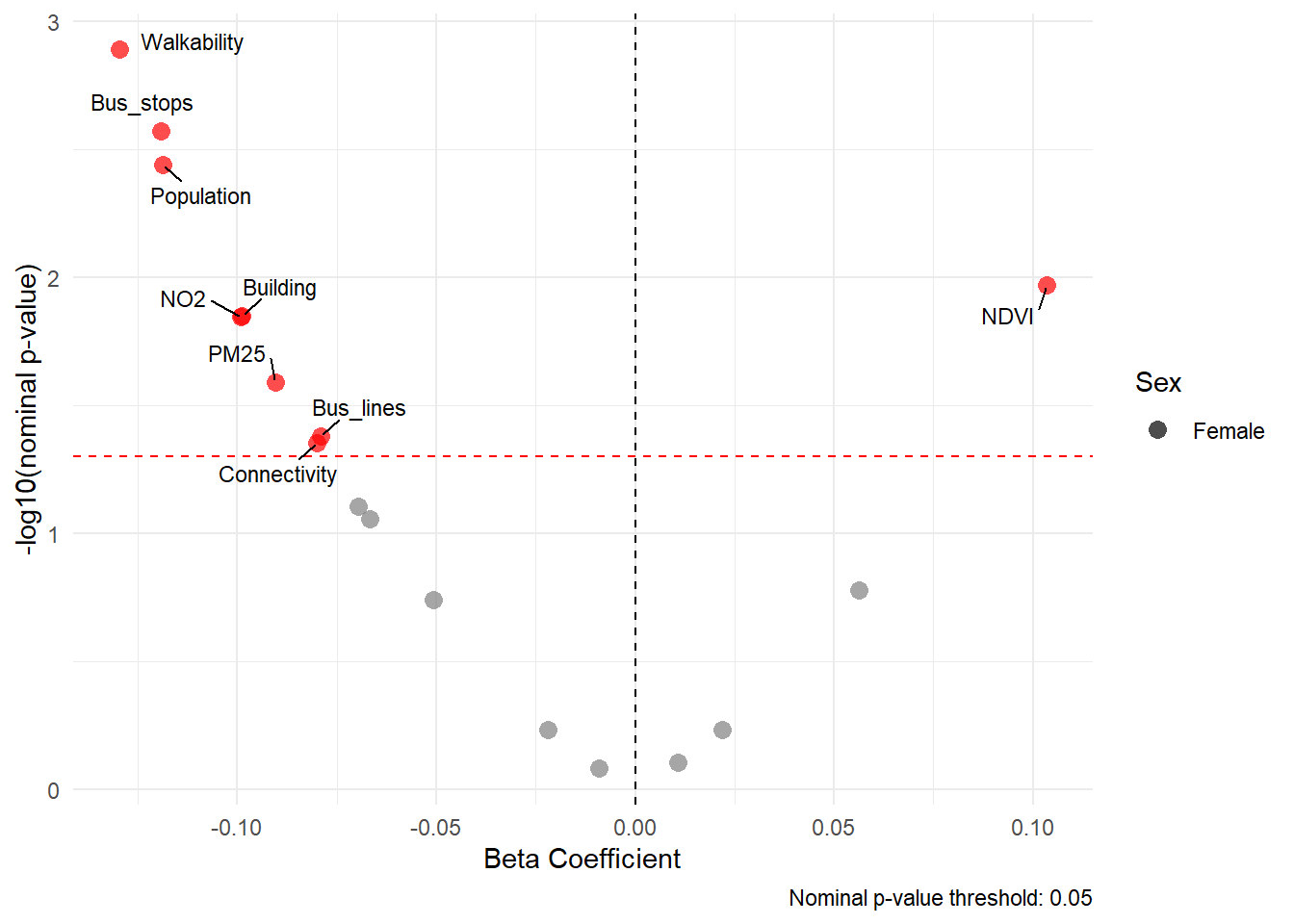
p)	q)

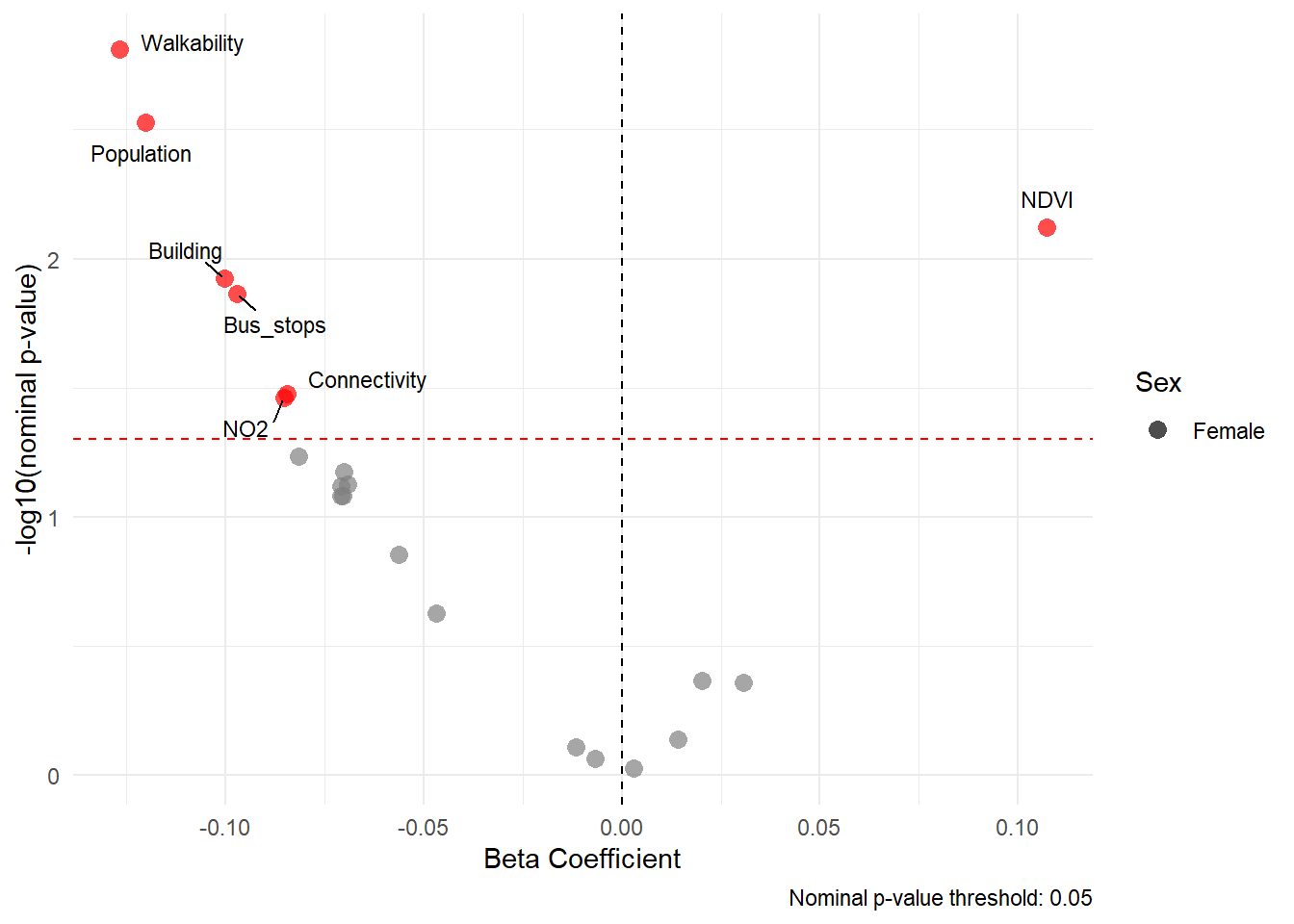

r)

**Supplementary Figure 3. Volcano plots of the Exposure Wild Association Study (ExWAS, single exposure models) analyses between the urban environment, breastfeeding duration and diet, and pubertal development at 9-10 years old, using non-FDR-corrected p-values**. INMA: a) Pregnancy exposure and pubertal timing defined by PDS score; b) Birth exposure and pubertal timing defined by PDS score; c) 14-18 months exposure and pubertal timing defined by PDS score; d) 4-5 years exposure and pubertal timing defined by PDS score; e) Pregnancy exposure and pubertal timing defined by Tanner stages; f) Birth exposure and pubertal timing defined by Tanner stages; g) 14-18 months exposure and pubertal timing defined by Tanner stages; h) 4-5 years exposure and pubertal timing defined by Tanner stages; i) Pregnancy exposure and age at menarche; j) Birth exposure and age at menarche; k) 14-18 months exposure and age at menarche; l) 4-5 years exposure and age at menarche; G21: m) Birth exposure and pubertal timing defined by Tanner stages; n) 1 year exposure and pubertal timing defined by Tanner stages; o) 4 years exposure and pubertal timing defined by Tanner stages; p) Birth exposure and age at menarche; q) 1 year exposure and age at menarche; r) 4 years exposure and age at menarche. Abbreviations: Access_points, number of bus public transport mode stops; Building, building density; Blue_dist, distance to nearest major blue space; Blue_size, size of the nearest major blue space; Green_dist, distance to nearest major green space; Green_size, size of the nearest major green space; Bus_lines, length of public transport lines; Bus_stops, density of public bus stops; Connectivity, connectivity density; Distance road, inverse distance to nearest road; Facility_dens, facility density; Facility_rich, facility richness; Unhealthy food, unhealthy food facility density; Land use, land use Shannon’s Evenness Index; NDVI, Normalized Difference Vegetation Index ; NO2, nitrogen dioxide; PM25, particulate matter with an aerodynamic diameter of less than 2.5 μm; Population, population density; Traffic_100, traffic load on all roads in 100m buffer; Traffic_dens, traffic density in nearest road; UV, average of vitamin D UV dose; Walkability, walkability index; F&V, fruit and vegetables; EDF, energy-dense foods; HEI, healthy eating index; BF_any, non-exclusive breastfeeding duration; BF_exc, exclusive breastfeeding duration.
